# *CADM2* is implicated in impulsive personality and numerous other traits by genome- and phenome-wide association studies in humans and mice

**DOI:** 10.1101/2022.01.29.22270095

**Authors:** Sandra Sanchez-Roige, Mariela V Jennings, Hayley H A Thorpe, Jazlene E Mallari, Lieke C van der Werf, Sevim B Bianchi, Calvin Lee, Travis T Mallard, Samuel A Barnes, Jin Yi Wu, Amanda M Barkley-Levenson, Ely C Boussaty, Cedric E Snethlage, Danielle Schafer, Zeljana Babic, Boyer D Winters, Katherine E Watters, Thomas Biederer, the 23andMe Research Team, James Mackillop, David N Stephens, Sarah L Elson, Pierre Fontanillas, Jibran Y Khokhar, Jared W Young, Abraham A Palmer

## Abstract

Impulsivity is a multidimensional heritable phenotype that broadly refers to the tendency to act prematurely and is associated with multiple forms of psychopathology, including substance use disorders. We performed genome-wide association studies (**GWAS**) of eight impulsive personality traits from the Barratt Impulsiveness Scale and the short UPPS-P Impulsive Personality Scale (N=123,509-133,517 23andMe research participants of European ancestry), and a measure of Drug Experimentation (N=130,684). Because these GWAS implicated the gene *CADM2*, we next performed single-SNP phenome-wide studies (**PheWAS**) of several of the implicated variants in *CADM2* in a multi-ancestral 23andMe cohort (N=3,229,317, European; N=579,623, Latin American; N=199,663, African American). Finally, we produced *Cadm2* mutant mice and used them to perform a Mouse-PheWAS (“MouseWAS”) by testing them with a battery of relevant behavioral tasks. In humans, impulsive personality traits showed modest chip-heritability (∼6-11%), and moderate genetic correlations (*r*_*g*_=.20-.50) with other personality traits, and various psychiatric and medical traits. We identified significant associations proximal to genes such as *TCF4* and *PTPRF*, and also identified nominal associations proximal to *DRD2* and *CRHR1*. PheWAS for *CADM2* variants identified associations with 378 traits in European participants, and 47 traits in Latin American participants, replicating associations with risky behaviors, cognition and BMI, and revealing novel associations including allergies, anxiety, irritable bowel syndrome, and migraine. Our MouseWAS recapitulated some of the associations found in humans, including impulsivity, cognition, and BMI. Our results further delineate the role of *CADM2* in impulsivity and numerous other psychiatric and somatic traits across ancestries and species.

## INTRODUCTION

Impulsivity is a multifaceted psychological construct that has been broadly defined as thoughts or actions that are “poorly conceived, prematurely expressed, unduly risky or inappropriate to the situation, and that often result in undesirable consequences” [1]. Impulsivity has been repeatedly associated with numerous psychiatric diseases, including ADHD and substance use disorders [2, 3]. We previously performed genome-wide association studies (**GWAS**) of impulsive personality traits (n=21,806-22,861) using two of the most widely used impulsivity questionnaires, the Barratt Impulsiveness Scale (**BIS-11**; 3 traits) and the Impulsive Personality Scale (**UPPS-P**; 5 traits), as well as a measure of Drug Experimentation [4]. These traits were partially genetically correlated, suggesting that each impulsivity domain is governed by overlapping but distinct biological mechanisms [4, 5]. Our work also identified significant genetic correlations between impulsivity and numerous psychiatric and substance use traits, in line with the NIMH Research Domain Criteria (**RDoC**), proposing impulsivity as a transdiagnostic endophenotype for psychopathology [6].

The cell adhesion molecule 2 (*CADM2*) gene, which was the most robustly implicated gene in our prior GWAS of impulsivity [4], has also been extensively implicated in other risky and substance use behaviors [7]. *CADM2* mediates synaptic plasticity and is enriched in the frontal cortex and striatum, which are regions that regulate reward and inhibitory processes. We and others have implicated this gene in traits that may underlie disinhibition in humans, supporting the observed genetic correlations between impulsivity and personality [8], educational attainment [9], cognition [10], risk-taking [11], substance use [4, 10, 12–15], externalizing psychopathology [16], neurodevelopmental disorders [17, 18], physical activity [19], reproductive health [20, 21], metabolic traits [22], and BMI [23], among others (see GWAS Catalog www.ebi.ac.uk/gwas/). *Cadm2* knockout mice have previously been assessed for body weight and energy homeostasis [24] but have never been behaviorally characterized for measures of impulsivity or related behaviors.

Here, we took three approaches to elucidate genetic factors related to impulsivity. First, we collaborated with 23andMe, Inc., to extend upon our earlier GWAS of impulsivity [4] by increasing our sample size approximately 6-fold (n=123,509-133,517). Second, we performed single-SNP phenome-wide studies (**PheWAS**) of the 5 single nucleotide polymorphisms (**SNPs**) in and around *CADM2* that have been most strongly implicated by the current and prior GWAS. PheWAS were conducted in three ancestral groups (N=3,229,317, European; N=579,623, Latin American; N=199,663, African American) from the 23andMe research cohort, examining close to 1,300 traits, most with no published GWAS. Finally, we performed a mouse-PheWAS (“MouseWAS”) by creating and phenotyping mice harboring a *Cadm2* mutant allele in a broad battery of behavioral tasks that included analogous human measures of risk-taking and impulsivity, substance use, cognition and BMI.

## MATERIALS AND METHODS

### Human Studies

#### GWAS cohort and phenotypes

We analyzed data from a cohort of up to 133,517 male and female research participants of European ancestry, a subset of which were analyzed in our prior publications [4, 13, 13, 25, 26]. All participants were drawn from the research participant base of 23andMe, Inc., a direct-to-consumer genetics company, and were not compensated for their participation. Participants provided informed consent and participated in the research online, under a protocol approved by the external AAHRPP-accredited IRB, Ethical & Independent Review Services (www.eandireview.com). During 4 months in 2015 and 14 months from 2018-2020, participants responded to a survey that included up to 139 questions pertaining to aspects of impulsivity and substance use and misuse. To measure impulsive personality, we used five subscales from the UPPS-P ([27, 28]; a 20-item that measures (lack of) Premeditation, (lack of) Perseverance, Positive Urgency, Negative Urgency, and Sensation Seeking; **Table S1**). We also administered the BIS-11 ([29]; a 30-item questionnaire that measures Attentional, Motor, and Nonplanning impulsiveness; **Table S1**). Lastly, we measured Drug Experimentation, defined as the number of substances an individual has used (adapted from the PhenX toolkit [30]; **Table S1**). We scored UPPS-P, BIS-11 and Drug Experimentation as previously described [4]. We used quantile normalization, since some scores were not normally distributed (**Figures S1-3**). Only individuals identified as European ancestry based on empirical genotype data [31] were included in this study. Basic demographic information about this sample is presented in **Table S2**. We used Pearson correlation coefficients (*r*) to measure the phenotypic relationships between impulsivity subscales and demographics.

#### Genome-wide association and secondary analyses

DNA extraction and genotyping were performed on saliva samples by CLIA-certified and CAP-accredited clinical laboratories of Laboratory Corporation of America. Quality control, imputation, and genome-wide analyses were performed by 23andMe (**Table S3**; [32, 33]). 23andMe’s analysis pipeline performs linear regression assuming an additive model for allelic effects (**Supplementary Material**). Covariates included age (inverse-normal transformed), sex, the top five principal genotype components, and indicator variables for genotyping platforms. *p*-values were corrected for genomic control. We examined genotype*sex interactions for suggestive loci. In addition, for the top loci, we examined African American and Latin American 23andMe research participants who had responded to the same survey.

We used the FUMA web-based platform (version 1.3.6a) and MAGMA v1.08 [34, 35] to explore the functional consequences of the GWAS loci and to conduct gene-based analyses.

We used LDSC [36] to calculate genetic correlations (r_g_) between UPPS-P, BIS and Drug Experimentation, and 96 selected traits informed by prior literature.

#### Phenome-wide association scan (PheWAS) in 23andMe

We performed single-SNP PheWAS for 5 *CADM2* SNPs (rs993137, rs62263923, rs11708632, rs818219, rs6803322) using up to 1,291 well-curated self-reported phenotypes from a separate cohort of 23andMe research participants of European (N≤3,229,317), Latin American (N≤579,623) and African American (N≤199,663) ancestries. We excluded traits with <1,000 responses, based on a prior simulation study for PheWAS power analysis [37]. Ancestry was determined by analyzing local ancestry ([31] **Supplementary Material**). The variants were selected based on our GWAS results and previous literature (**Table S4**, and **Supplementary Material**). Genotyped and imputed variant statistics in PheWAS are shown in **Table S5**.

An overview of the data collection process has been previously described [38]. All regression analyses were performed using R version 3.2.2. We assumed additive allelic effects and included covariates for age (as determined by participant date of birth), sex, and the top five ancestry-specific principal components. We used a 5% FDR correction for multiple testing.

### MouseWAS

#### Subjects, behavioral characterization, and statistical analyses

Our *Cadm2* mutant mice were produced at the University of California San Diego, Moores Cancer Center, Transgenic Mouse Core. We used the JM8.N4 cryosperm line (CSD70565 KOMP), which carries a floxed null allele in the *Cadm2* gene (**Figure S30**), on a C57BL/6N background. We crossed the floxed null allele line with a constitutive CRE driver line (Stock# 014094; The Jackson Laboratory), yielding a global constitutive null allele. We used a heterozygous x heterozygous (**HET**) breeding scheme, which produced homozygous (**HOM**) mutant *Cadm2* mice and their HET and wildtype (**WT**) littermates. Mice were genotyped using allele-specific polymerase chain reaction on ear notch tissue followed by gel electrophoresis [39]. CADM2 protein expression levels were quantified by western blotting (**Figure S31**).

Five separate cohorts of male and female mice were used for these studies. See **Supplementary Material** for a more detailed description of the tasks and analyses of main variables. Procedures were approved by the University of California San Diego Institutional Animal Care and Use Committee. The UCSD animal facility meets all federal and state requirements for animal care and was approved by the American Association for Accreditation of Laboratory Animal Care. Procedures from cohort 2 were conducted in accordance with the Canadian Council on Animal Care and were approved by the University of Guelph Institutional Animal Care and Use Committee.

## RESULTS

### Genome-wide association analyses and secondary analyses

Self-reported impulsivity and Drug Experimentation scores are shown in **Table S6**. We found that ∼6-11% of the phenotypic variation of these traits can be explained by common variants (**Table S7**). We identified 21 genome-wide significant associations (*p*<5.0E-08) for UPPS-P (5 traits), BIS (3 traits), and Drug Experimentation (**Figure 1**; **Figures S4-21**; **Table S8**). Although we tested 9 traits, in keeping with the standards of the field, we did not adjust the significance threshold. We also detected several nominal associations (*p*<1.0E-06, **Table S8**); we discuss some of them in the **Supplementary Material**.

**Figure 1.**
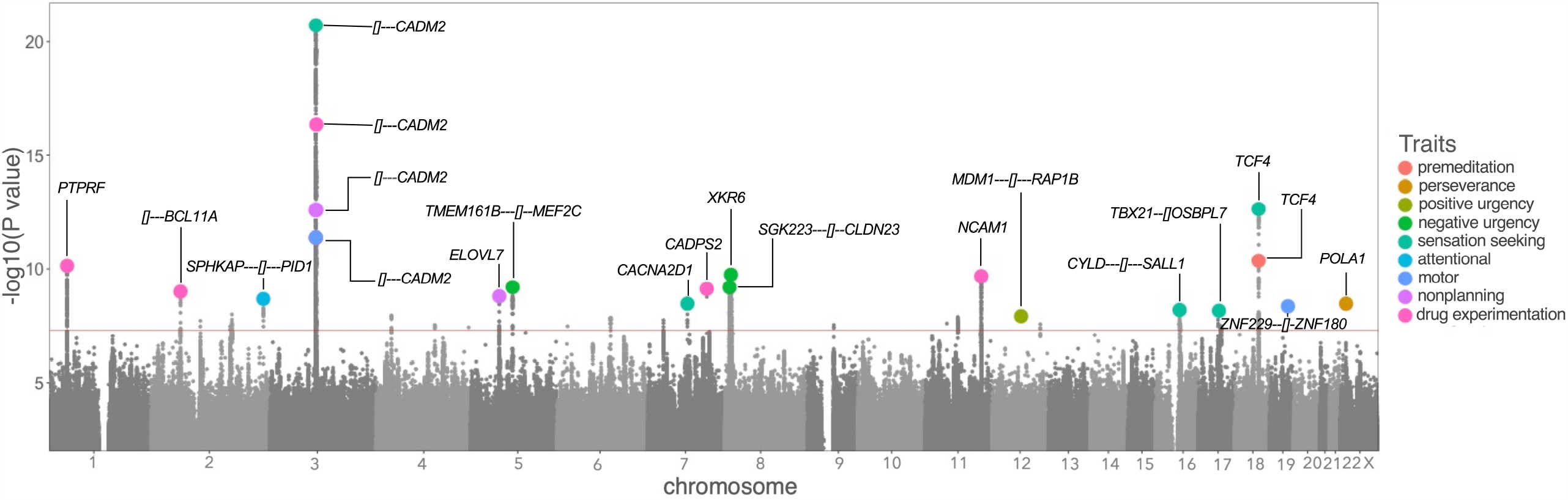
Porcupine plot displaying 21 genome-wide significant hits for all impulsivity facets and Drug Experimentation. *CADM2* was consistent across 3/8 impulsivity facets [Sensation Seeking (UPPS-P), Motor and Nonplanning impulsivity (BIS-11)] at a genome-wide association level, and with 3 more impulsivity facets [Attentional (BIS-11), Negative Urgency and Premeditation (UPPS-P]), at a gene-based level (**Table S8**). *CADM2* was also associated with risky behavior, such as Drug Experimentation.

#### GWAS of UPPS-P

##### Premeditation

We detected one significant hit (rs2958162, *p*=2.50E-10), located on chromosome 18 in the *TCF4* gene, which encodes a helix-loop-helix transcription factor and is widely expressed throughout the body and during development. Polymorphisms in *TCF4* have been associated with risk-taking and adventurousness [15], alcohol consumption [40], schizophrenia [41], depression [42, 43], and neuroticism [44, 45] (**Table S9**); *TCF4* is also a non-GWAS candidate gene for other psychiatric and neurological conditions [46].

##### Perseverance

We detected one significant association (rs5943997, *p*=1.50E-8) in the *POLA1* gene on the X chromosome. *POLA1* has been related to blood traits [46] and neurodevelopmental disorders [47], but its association with impulsivity is novel.

##### Positive Urgency

We identified one significant hit (rs143987963, *p*=4.30E-08) on chromosome 12, near the genes *MDM1* and *RAP1B*; however, inspection of the locus zoom plot (**Figure S9**) does not support a robust association.

##### Negative Urgency

We detected three significant hits: rs4840542 (*p*=1.60E-09), on chromosome 8, in the *XKR6* gene; rs5008475 (*p*=4.90E-09), on chromosome 5, near *TMEM161B* and *MEF2C*; and rs7829975, on chromosome 8, near *SGK223* and *CLDN23* (*p*=5.00E-09). Variants in strong LD with rs4840542 and rs7829975 are highly pleiotropic, and have been previously associated with several traits (**Table S9**), including body mass index (**BMI**) [48, 49], neuroticism [50, 51], depression [52], blood pressure, and alcohol consumption [53]. *XKR6* was also implicated in a recent GWAS of externalizing [16], and a GWAS of anxiety and depression [52].

##### Sensation Seeking

We detected 5 significant associations. First, we again observed a previously reported [4] association with a SNP near *CADM2* (rs11288859, *p*=2.10E-09) on chromosome 3. We also detected an association with a SNP in *TCF4* (rs2958178, *p*=3.80E-12). We identified a significant hit in *CACNA2D1* (rs38547, *p*=2.10E-08) on chromosome 18. *CACNA2D1* has been previously associated with feeling nervous [50], and levels of sex hormone-binding globulin [54]. Furthermore, we found a significant association (rs1605379, *p*=3.80E-08) on chromosome 16, near *CYLD* and *SALL1*. SNPs in strong LD with rs1605379 have been previously identified for risk-taking, adventurousness, and smoking initiation (**Table S9**). Lastly, we found a significant association (rs12600879, *p*=4.10E-08) on chromosome 17, near *TBX21* and *OSBPL7*. Variants in strong LD with rs12600879 have been associated with BMI [55], but the finding in relation to impulsivity is novel.

#### GWAS of BIS-11

##### Attentional

We identified one significant association (rs10196237, *p*=1.10E-08) on chromosome 2, near the genes *SPHKAP and PID1. SPHKAP* has been previously associated with educational attainment [9], but the association with impulsivity is novel.

##### Motor

We detected one significant association near *CADM2* (rs35614735, *p*=3.20E-11). We also identified an association (rs111502401, *p*=2.00E-08), on chromosome 19, near the genes *ZNF229* and *ZNF180*; however, inspection of the regional association is not supportive of a strong association (**Figure S17**).

##### Nonplanning

We detected 2 variants: rs35614735 (*p*=4.70E-12) near *CADM2*, which was the same SNP identified for Motor impulsivity; and rs6872863 (*p*=1.20E-08) in the gene *ELOVL7* on chromosome 5. Variants in strong LD with rs6872863 have been reported for a variety of traits including educational attainment, mathematical ability [9], household income [56], and brain morphology, such as cortical surface area [57] (**Table S9**). However, there is extensive LD in this region, making the association difficult to interpret.

#### GWAS of Drug Experimentation

We previously reported [4] a suggestive association (rs2163971, *p*=3.00E-07) near the *CADM2* gene. In the present study, we identified a nearby SNP that was genome-wide significant (rs35614735, *p*=2.80E-15). We also report 4 novel hits (rs951740, *p*=9.70E-10, *PTPRF* on chromosome 1; rs12713405, *p*=9.70E-09, *BLC11A* on chromosome 2; rs67660520, *p*=7.60E-09, *CADPS2* on chromosome 7; rs7128648, *p*=2.50E-09, *NCAM1* on chromosome 11). Intriguingly, *PTPRF* has been recently associated with problematic prescription opioid use [25] and opioid use disorder [58], as well as smoking initiation/cessation [59], cognition [60], and educational attainment [9] (**Table S9**). Variants in strong LD with rs67660520 have been associated with ADHD [61], smoking initiation [59], number of sexual partners [15] and BMI [49] (**Table S9**). *NCAM1* variants have been previously associated with alcohol, cannabis and smoking behaviors [59, 63], mathematical ability [9], and anxiety and depression [52], among other traits.

#### Gene-based analyses

Similar to the GWAS results, gene-based analyses using MAGMA identified an association (Bonferroni *p*<2.53E-06; **Table S10**) between *CADM2* and 6 of the 9 traits examined in this paper: Premeditation, Sensation Seeking (UPPS-P); Attentional, Motor and Nonplanning (BIS-11); and Drug Experimentation. *TCF4*, which was significantly associated with Premeditation and Sensation Seeking in the GWAS, was significantly associated with these traits in the gene-based analysis. *MAPT*, which has been previously associated with many traits including multiple alcohol-related behaviors [13], was implicated in Negative Urgency. Lastly, *KDM4A*, which was recently related to problematic opioid use and interacts with selective serotonin reuptake inhibitors and dopaminergic agents [25], was significantly associated with Drug Experimentation.

#### Phenotypic and genetic correlations

A phenotypic and genetic correlation matrix of all 9 traits is shown in **Figure S22** and **Tables S11-12**. Consistent with the literature and our prior work [4, 5, 64, 65], both phenotypic and genetic inter-correlations among the UPPS-P and BIS subscales were high and positive, with the exception of Sensation Seeking and Perseverance, suggesting that these traits may represent relatively different constructs [5, 13, 64]. Drug experimentation was positively and significantly associated with all impulsive personality traits.

All impulsivity traits were phenotypically associated (*r*=-0.34-0.11) with demographic variables (**Table S12**), impulsivity scores being greater in male and younger research participants, compared to female and older participants; and in participants with higher BMI, lower household income, and fewer years of education, as we previously reported [13].

Figure 2. shows a genetic correlation matrix of BIS, UPPS-P, Drug Experimentation and several other phenotypes (full results in **Table S13**). As anticipated, we found positive moderate to high genetic correlations (*r*_*g*_=0.25-0.79) between virtually all UPPS-P (except Perseverance and Sensation Seeking) and BIS subscales, and Drug Experimentation, and substance use disorders **Table S13**).

**Figure 2.**
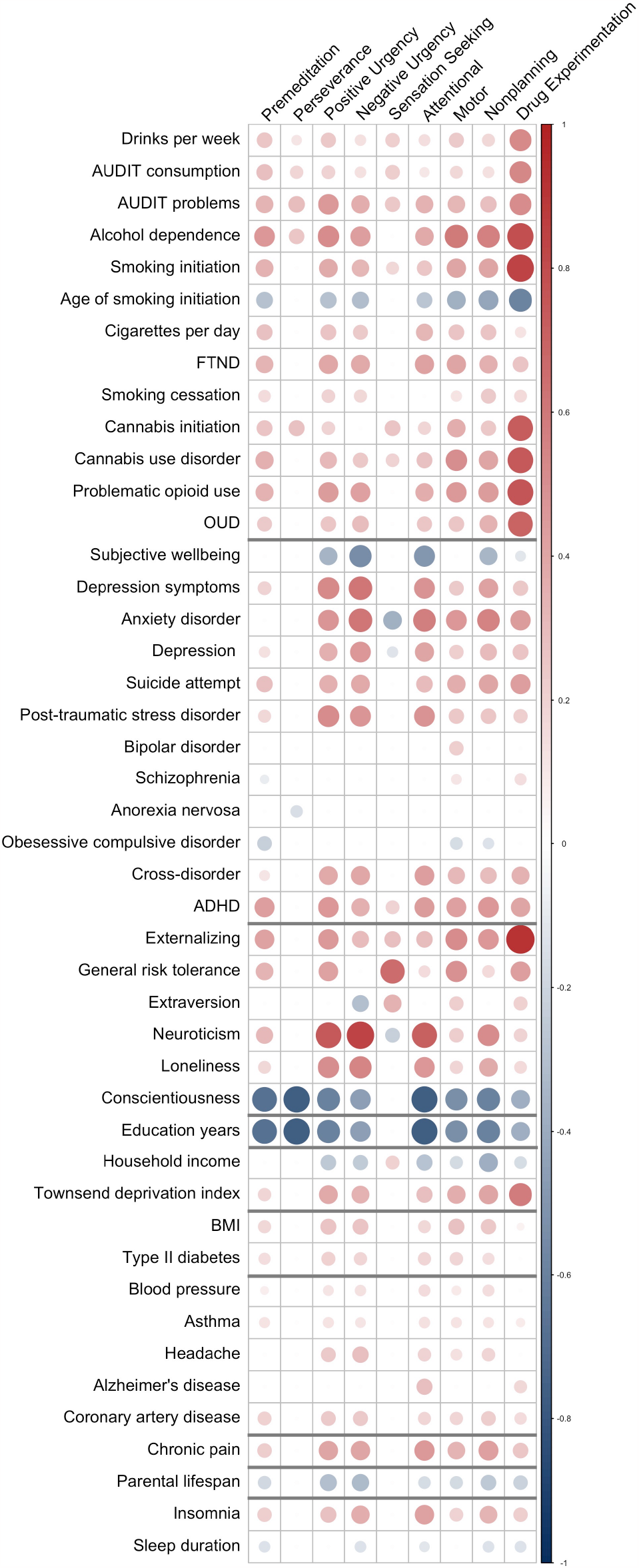
Genetic correlations (*r*_*g*_) between UPPS-P, BIS, and Drug Experimentation, and other substance use, psychiatric, personality, cognitive, metabolic, health, pain, longevity and sleep traits (see **Table S9** for full results). All values survive 5% FDR correction.

We also observed moderate to strong associations between all impulsive subscales (except UPPS-P Perseverance) and other personality traits, such as risk-taking (*r*_*g*_=0.15-0.65), neuroticism (*r*_*g*_=-0.23-0.84), and loneliness (*r*_*g*_=0.17-0.54), particularly for Positive and Negative Urgency. Extraversion was positively associated with Sensation Seeking (*r*_*g*_=0.34). Externalizing psychopathology, which represents disorders and behaviors characterized by deficits in inhibition, was strongly associated with all impulsivity facets (*r*_*g*_=0.28-0.92), except Perseverance.

We also identified positive associations with an array of psychiatric phenotypes, including ADHD (*r*_*g*_=0.20-0.47), depression (*r*_*g*_=-0.13-0.47) and anxiety (*r*_*g*_=-0.38-0.61) disorders, and cross-disorder (*r*_*g*_=0.12-0.44). The associations were again primarily significant for all except Perseverance and Sensation Seeking. Other disorders showed weaker associations (e.g., schizophrenia, *r*_*g*_=-0.09-0.15) or were only significantly associated with one impulsivity facet [e.g., anorexia nervosa (Perseverance, *r*_*g*_=-0.16); bipolar disorder (Motor, *r*_*g*_=0.22)].

Most impulsivity subscales were genetically correlated with lower socioeconomic variables [e.g., educational attainment (*r*_*g*_=-0.49 to -0.16), income (*r*_*g*_=-0.38 to -0.16), Townsend index (*r*_*g*_=0.18-0.58)].

Metabolic and medical phenotypes, such as BMI (*r*_*g*_=0.18-0.28), chronic pain (*r*_*g*_=0.22-0.46), insomnia (*r*_*g*_=0.20-0.42), and coronary artery disease (*r*_*g*_=0.18-0.30), were genetically correlated with all impulsive subscales (except Perseverance and Sensation Seeking). We also noted negative genetic associations with parental longevity (*r*_*g*_=-0.17 to -0.32).

### PheWAS

To explore the impact of specific variants in and around *CADM2*, we performed single-SNP PheWAS using 5 of the most implicated SNPs, independently, against 1,291 traits (**Figure 3**). The list of PheWAS association results using the 23andMe cohort after 5% FDR correction is available in **Tables S14** (summary), **S15** (Europeans), **S16** (Latin American) and **S17** (African Americans).

**Figure 3.**
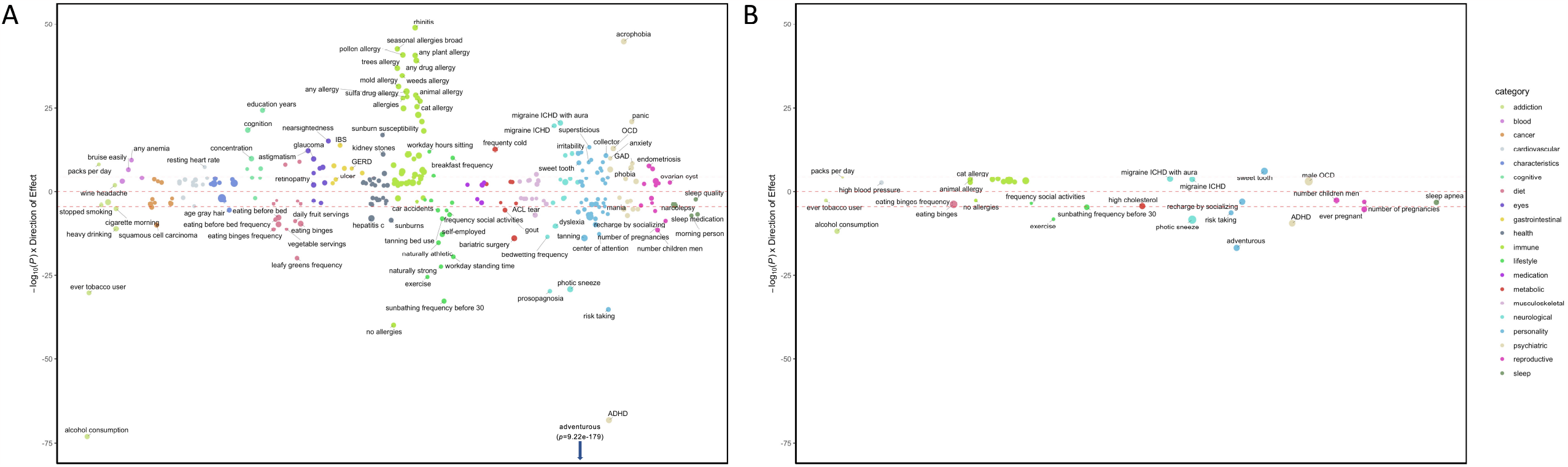
FDR-significant associations from *CADM2* single-SNP PheWAS in individuals of European ancestry (**A**) and Latin American ancestry (**B**). Results for the SNP with the highest number of significant PheWAS associations (rs993137) are presented; results for additional SNPs are included in Supplementary Tables 15-17. No FDR-significant findings were detected in individuals of African American ancestry. The size of the dots represents the magnitude of the effect size for each trait. The effect sizes ranged from -0.14 to 0.13 in the European cohort and from -0.08 to 0.16 in the Latin American cohort. Dotted line denotes Bonferroni significance (*p*<3.79E-05).

In European cohorts, *CADM2* variants had been previously identified to be significantly associated with numerous traits (**Table S18**). Most SNPs were highly correlated (R^2^>0.1) and tagged similar traits (**Figure S23**), but the overlap was incomplete (**Figure S24** and **Table S19**). rs993137, located at 85,449,885 bp on chromosome 3, showed the highest number of associations (378), which we describe below.

We replicated all previously known associations in 23andMe participants of European ancestry, identifying signals across all categories tested (**Table S15**). These included negative associations with risky behavior (e.g., lower risk for adventurousness [β=-0.05, *p*=1.33E-08], risk-taking tendencies [β=-0.02, *p*=1.13E-07]) and substance use behaviors (e.g., lower risk for alcohol consumption [β=-0.03, *p*=2.05E-09] and tobacco initiation (β=-0.02, *p*=3.66E-12; but see packs per day, β=0.01, *p*=1.05E-03), as well as negative associations with psychiatric disorders characterized by deficits in impulsivity, such as lower risk for ADHD (β=-0.05, *p*=2.17E-41). Furthermore, we found positive associations with educational outcomes (e.g., higher educational attainment (β=0.03, *p*=1.67E-12). Novel findings included positive associations with allergies (β=0.04, *p*=4.51E-03), anxiety (e.g., panic [β=0.02, *p*=6.82E-08]), and medical conditions (e.g., IBS [β=0.02, *p*=8.89E-07]), anemia (β=0.01, *p*=8.30E-74), hepatitis C (β=-0.06, p=8.36E-10). Intriguingly, we also detected positive associations with pain phenotypes (β=0.02, p=8.37E-12) and a need for a higher dose of pain medication (β=0.01, *p*=1.02E-06).

For the overlapping phenotypes, UK Biobank PheWAS results [66] largely supported the 23andMe PheWAS findings (except for smoking behaviors). For example, we identified associations with dietary traits (e.g., daily fruit and vegetable intake (β=-0.01, *p*=4.23E-11), pastry frequency (β=0.01, *p*=7.36E-06), sleep quality (β=-0.01, *p*=2.53E-03), and number of pregnancies (β=-0.01, *p*=7.69E-04), among others (**Table S15**, [12]).

For the PheWAS of the Latin American cohort, 47 traits were significantly associated with *CADM2* variants (**Table S16**). The highest number of associations were again observed for rs993137 [67], which are described below. Similarly, although some of the SNPs were correlated (R^2^>0.1; **Figure S24**), the overlap was incomplete (**Figure S26, Table S20**). The pattern of associations was consistent with those described in the European cohort. The strongest associations were with risky behaviors, such as adventurousness (β=-0.04, *p*=1.76E-17), risk-taking (β=-0.02, *p*=5.90E-07), alcohol consumption (β=-0.03, *p*=1.41E-12), and disorders characterized by high levels of impulsivity, such as ADHD (β=-0.04, *p*=4.74E-10). The novel findings were, again, with multiple forms of allergies (e.g., seasonal allergies, β=0.03, *p*=3.0E-04), migraine (β=0.04, *p*=1.56E-04), sleep behaviors (e.g., sleep apnea, β=-0.03, *p*=6.76E-04), among others.

All findings that were in common between the European and Latin American cohorts showed the same direction of effect and similar effect sizes. We did not identify FDR-significant associations in the African American cohort (**Table S17**). The effect sizes were generally extremely small (**Figures S27-28**), as is expected for a single gene and complex traits.

### MouseWAS

Figure 4. summarizes the MouseWAS results across the five cohorts tested. Full statistics and additional secondary measures are described in the **Supplementary Material** and **Table S20**.

**Figure 4.**
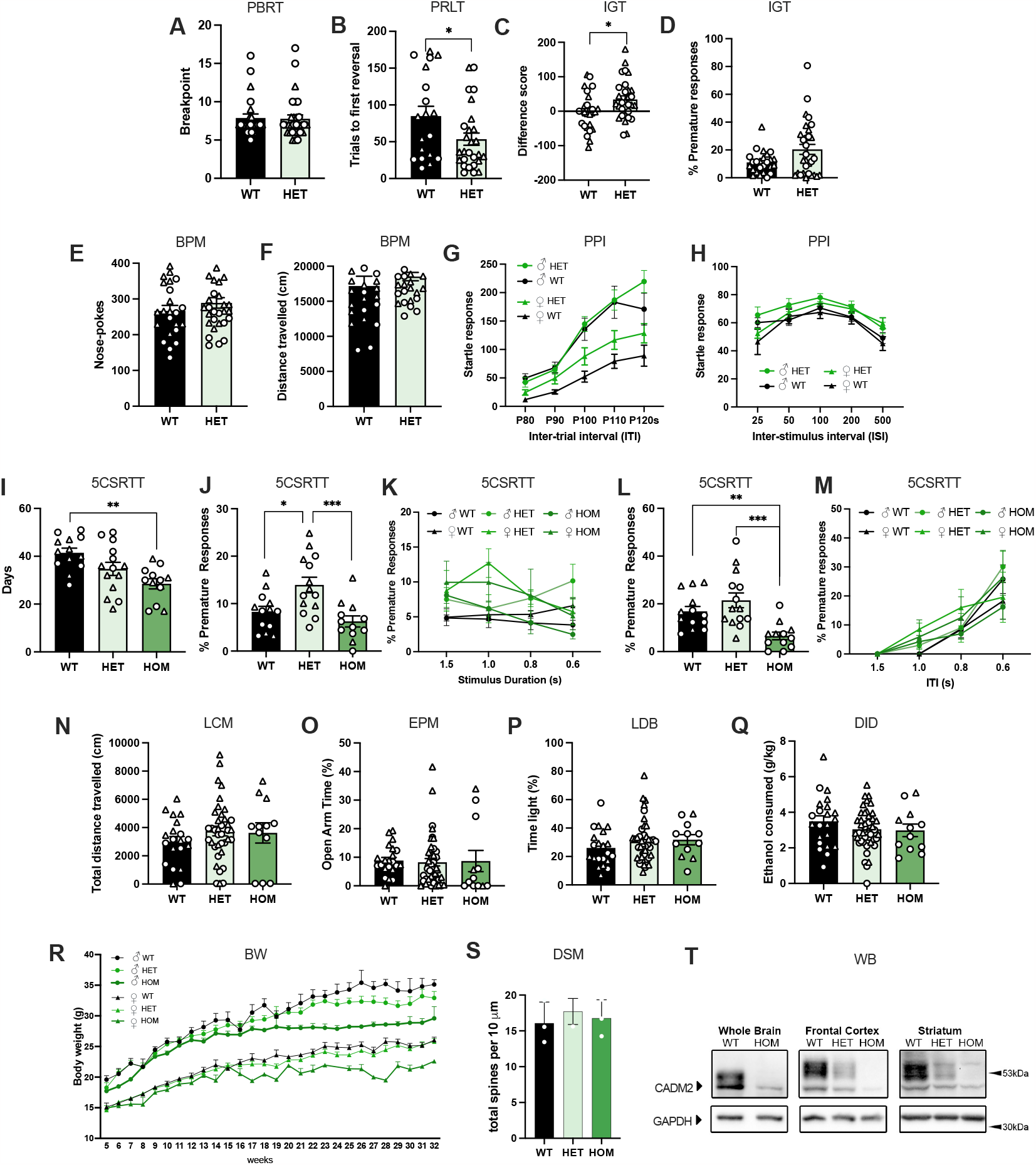
MouseWAS examined the complete and partial loss of *Cadm2* function and behavioral consequences in the Probability Breakpoint Ratio Task (PBRT, **A**), Probabilistic Reversal Learning Task (PRLT, **B**), Iowa gambling task (IGT, **C-D**), Behavioral Pattern Monitor task (BPM, **E-F**), Prepulse Inhibition (PPI, **G-H**), 5-Choice Serial Reaction Time Task (5CSRTT) performance (**I-M**), Locomotor (LCM) activity (**N**), Elevated Plus Maze (EPM, **O**), Light-Dark Box (LDB, **P**), and Drinking in the Dark (DID, **Q**); longitudinal body weight changes (**R**), and dendrite morphology (DSM) in the nucleus accumbens (**S**). Western blot (**WB**) analysis (**T**) of Cadm2 protein in whole brain, frontal cortex and striatum. WT, wildtype, HET, heterozygote, HOM, homozygote. Sample size by cohort: cohort 1 (WT=25, HET=30, HOM=3), cohort 2 (WT=13; HET=14, HOM=12), cohort 3 (WT=22; HET=44, HOM=12), cohort 4 (WT=29, HT=54, HOM=17), cohort 5 (WT=3, HET=3, HOM=3). In cohort 1, the sample size of the HOM mice deviated from the expected Mendelian frequency for unknown reasons; these animals were excluded from the analyses. Males are represented in circles, females in triangles. * *p*<0.05, ** *p*<0.01, *** *p*<0.001.

#### Cohort 1 -Motivation, inhibition, and risk-taking behavior

No differences in motivation were found between WT and HET mice during the Progressive Breakpoint task [F_(1,51)_=0.003, *p*=9.57E-01; **Figure 4A**]. However, we noted significant genotype differences in behavioral flexibility in the Probabilistic Reversal Learning (**PRL**) task, as indexed by the number of trials to first reversal [F_(1,42)_=4.27, *p*=4.50E-02; **Figure 4B**], and risky behavior in the mouse Iowa Gambling Task (**IGT**; F_(1,51)_=4.70, *p*=3.50E-02; **Figure 4C**], HET mice requiring fewer trials to reach criterion and choosing risky options less frequently than WT mice (*p*<0.05), respectively. The number of premature responses, on the contrary, were higher in HET mice [F_(1,51)_=5.78, *p*=2.00E-02] compared to WT mice (*p*<0.05; **Figure 4D**). In the Behavioral pattern monitor (**BPM**), HET mice exhibited greater exploratory behavior, as shown by an increase in hole-pokes [F_(1,53)_= 4.88, *p*=3.20E-02; **Figure 4E**], compared to WT mice (*p*<0.05), but general levels of activity, such as distance traveled (F_(1,53)_= 0.42, *p*=5.21E-01; **Figure 4F**), were similar across the genotypes. Lastly, although the startle response was equal across the groups (**Figure 4G**), prepulse inhibition (**PPI**) was larger in HET mice compared to WT mice (*p*<0.05; **Figure 4H**), particularly at ISI 25 and 100 in HET mice [F_(1,53)_=8.23, *p*=6.00E-03, F_(1,53)_=4.50, *p*=3.90E-02, respectively].

#### Cohort 2 -Motoric impulsivity

The main outcome tested in cohort 2 were premature responses via the 5-choice serial reaction time task (**5CSRTT**; **Figure 4J-M**). Premature responses were lower in HOM (*p*<0.001) and WT (*p*<0.02) mice compared to HET mice under standard conditions (F_(2,36)_=8.74, *p*=8.06E-04; **Figure 4J**), and compared to both HET (*p*<0.001) and WT (*p*<0.01) mice during a long ITI session (H_(2)_=16.10, *p*=3.19E-04; **Figure 4L**). HOM mice were faster at learning the 5CSRTT, requiring fewer days for adequate baseline performance (F_(2,36)_=7.42, *p*=2.00E-03; **Figure 4I**), compared to WT mice (*p*<0.01).

#### Cohort 3 - General locomotion, anxiety-like behavior, and ethanol consumption

We found a significant effect of genotype on the distance traveled in the Open Field [**OF**; F_(2,70)_=7.525, *p*=1.00E-03; **Figure 4N**], HOM mice showing higher levels of locomotor activity than WT mice (*p*=1.40E-02). No differences in anxiety-like behavior were detected across WT, HET or HOM mice in the Elevated Plus Maze (**EPM**) or Light-Dark Box (**LDB**) tests (**Figure 4O-P, Table S20**). The total amount of ethanol consumed during the drinking-in-the-dark (**DID**) paradigm did not differ between the groups ([F_(2,78)_=1.084, *p*=3.44E-01]; **Figure 4Q**).

#### Cohort 4 - Body weight

Relative to WT mice, there was a significant reduction in body weight in HOM mice from week 21 onwards (β=-3.74±1.27, *p*=4.00E-03; **Figure 4R**).

#### Cohort 5 - Dendrite morphology

Quantitative analyses of MSN in the NAc revealed no difference in dendritic spine density across the groups (**Figure 5S**).

## DISCUSSION

In this study, we performed the largest GWAS of impulsive personality traits to date, we conducted the first multi-ancestral PheWAS exploring the role of *CADM2* on a diverse array of traits, and we performed a corresponding MouseWAS using *Cadm2* mutant mice to assess its role in impulsivity and other relevant behaviors. We extend on prior findings [4, 5] showing that the genetic architecture of impulsivity facets only partially overlap, providing further support to the idea of impulsivity being a multifaceted construct even at the genetic level. We identified positive genetic correlations across multiple domains, particularly substance use disorders, confirming that NIMH RDoC transdiagnostic domains [6], or endophenotypes, such as impulsive personality traits, can be used to dissect the genetic basis of psychiatric illness and normal functioning. RDoC or transdiagnostic traits are beneficial because they enable translational research and provide a more granular biological understanding of psychiatric disorders. Using mouse and human correlates, we provide further evidence that *CADM2* is a robust candidate gene for impulsivity and an important modulator of numerous other psychiatric and somatic traits.

We increased the sample size of our prior GWAS of impulsivity by almost 6-fold and identified 21 genome-wide significant loci implicated in impulsive personality and Drug Experimentation. For instance, SNPs located in the gene *TCF4* were implicated in 3 subscales; this gene is also highly pleiotropic for other psychiatric conditions. Furthermore, we identified associations with *NCAM1*, which, intriguingly, is a critical member of the NTAD (*NCAM1-TTC12-ANKK1-DRD2*) gene cluster [68] and variants correlated with *NCAM1* in that cluster have been associated with differences in D2 receptor density [69]. We also detected associations near *XKR6* and *AFF3*, which have been recently implicated in externalizing psychopathology [16], and *PTPRF* and *KDM4A*, recently implicated in problematic opioid use [25] and opioid use disorder [58]. Although in this report we focused on *CADM2*, functional studies of those genes are also warranted. Furthermore, we found nominal evidence for candidate gene studies implicating monoamine neurotransmitters in impulsivity and Drug Experimentation (*DRD2, HTR3B*). High impulsivity depends on a neural network that includes the ventral striatum (subsuming the NAc) with top-down control from prefrontal cortical regions, and is modulated by monoamine neurotransmitters including dopamine and serotonin [70]; this is the first GWAS to implicate genes modulating these systems as robust candidate genes for impulsivity.

Recent studies have implicated the *CADM2* gene in impulsivity and traits associated with reward sensitivity and multiple domains of human health. We confirmed numerous previously reported associations and extended our findings of variants related to *CADM2. CADM2* was significantly associated with 4 out of the 9 traits that we measured in GWAS and 6 out of the 9 traits that we measured in gene-based analyses. In the PheWAS, *CADM2* variants were associated with decreased risk for externalizing psychopathology, but also increased risk for internalizing psychopathology (anxiety, depression, OCD). We also observed novel associations with migraines and various allergies. Others using a similar approach with UK Biobank data have found that this enrichment of associations is higher than expected [12] compared to other genes. These results provide evidence that *CADM2* variants are associated with broad health outcomes, but whether this gene affects human health via disruptions in inhibitory control or reward systems, or whether it acts via multiple pathways [71], is still not fully understood.

A relatively unique feature of our study is that, to follow up on the *CADM2* loci implicated in human studies, we generated a *Cadm2* mutant mouse line and used it to perform a PheWAS-like study in mice, which we have termed a MouseWAS to emphasize its conceptual similarity to human PheWAS studies. These functional experiments provided information about the causality and directionality of effects in its reported associations. We found evidence that loss of *Cadm2* resulted in *less* risky behavior and *improved* information processing, extending on prior work in humans [4, 10, 16, 62, 69].

*Cadm2* expression may uniquely contribute to the different domains of impulsivity. The IGT assays preference for high risk, high reward (disadvantageous) choices vs low risk, low reward (advantageous) choices [72]. HET mice exhibited a greater preference for selecting the safe option vs their WT littermates. This finding can be contrasted with the *elevated* premature responses in the 5CSRTT seen in HET vs WT mice, reflective of motoric impulsivity. However, premature responses have also been linked to temporal discrimination, wherein mice and humans overestimating the passage of time exhibit higher premature responses [73, 74]. The preference for less risky options of HET mice in the IGT could reflect their misjudgment of time – resulting in higher premature responses – and thus avoidance of higher temporal punishment in the IGT.

We also observed genotype differences in performance that could be indicative of *Cadm2* function in information processing. HET mice exhibiting *better* PPI at the shortest temporal window (25 ms) supports the premise that these mice have faster processing speeds. HET mice also showed small increases in hole-poking in the BPM test, which is thought to reflect exploration of an environment and information gathering. Finally, we observed that HOM mice acquired 5CSRTT faster than WT littermates. Taken together, these results suggest that *Cadm2* reduction may improve some facets of information processing.

Findings from the 5CSRTT provide evidence that *Cadm2* deletion improves some information processing and impulsivity outcomes, while being detrimental to others. HET mice were the most likely to commit 5CSRTT premature responses, although HOM mice were surprisingly the least likely to make premature responses. Interestingly, although not significant, there was a consistent elevation in the number of premature responses committed by the HOM mice as the stimulus duration was reduced. This could suggest that HOM mice, like HET mice, may show motoric impulsivity deficits when performing tasks that require greater attentional demand. Compared with WT, HOM mice also showed impaired accuracy performance under RSD conditions, in line with our human findings of *CADM2* association with BIS Attentional, and cognitive function by others [10]. The heterogeneity of performance outcomes in the HOM mice further supports a unique but overlapping contribution of genetics across impulsivity domains.

In this paper, we have translated measures from human to mice. These studies begin the process of understanding the biological basis of associations identified by GWAS. The methods for measuring impulsivity in humans and mice are fundamentally different. Despite these differences, our MouseWAS identified several measures of impulsivity that were influenced by *Cadm2*, consistent with our observations in humans. Furthermore, *CADM2* has been shown to be implicated in BMI in humans [24, 75] and energy homeostasis in mice [24]; extending on this, we found novel evidence of body weight reductions in adult mutant mice. Interestingly, *Cadm2* did not have more general effects on mouse behavior; for instance, we did not observe deficits in anxiety-like behavior or general motivation, as some of the human PheWAS findings revealed. A few other measures were also inconsistent across species, particularly measures of alcohol consumption, where *CADM2* showed a role in humans [7, 13, 76, 77] but not mice. Lastly, some measures identified by our human PheWAS (e.g., allergies and other medical conditions) were not examined in our MouseWAS. This approach highlights the challenges of using mouse models to further investigate the role of specific genes in behavioral traits.

*CADM2* encodes the immunoglobulin adhesion protein SynCAM 2, which is part of the family of synaptic adhesion molecules known as SynCAMs. Studies have shown the influence of SynCAMs on synaptogenesis [78–82], axon guidance [83], and neuron myelination [84–86], processes that have direct effects on the pathology of neurodevelopmental diseases [56]. *CADM2* is strongly expressed in the striatum and frontal cortex, which are core regions that regulate impulsivity [70]. We did not observe changes in spine density in the Nac, which suggests that *Cadm2* may not have a role as a postsynaptic organizer of spines in this region, or may have redundant functions that are compensated in the mutant mice by other molecules. Based on *in-silico* analyses in humans, *CADM2* expression seems to be greater at earlier stages of development (**Figure S29**); whether *Cadm2* may affect earlier stages of development (prenatal and early postnatal) that are compensated in adulthood has not been investigated in this study.

Several limitations of this study are worth noting. The discovery GWAS only includes male and female participants of European ancestry. While we provided exploratory analyses of top variants in other ancestries and by broken down by sex (**Supplementary Table 22**), larger sample sizes would be needed to perform GWAS separately in males and females. Our results are also biased by potential ascertainment and characteristics of the sample; the 23andMe participant population is more educated and has higher socioeconomic status and lower levels of drug use and impulsivity than the general US population [86]. Replication in additional cohorts with different characteristics is warranted. Moreover, although the traits we studied are extracted via well-established questionnaires, they are self-reported measures, which are different from behavioral phenotypes [87, 88]. Another issue is that, although we tested multiple variants in the *CADM2* loci, further conditional analyses are required to determine if this signal and previously reported associations implicating *CADM2* loci, including a large non-coding rare deletion in the first intron of *CADM2* [71], may tag the same underlying genetic effect. We are also unaware of the sequence of events, and whether there is true pleiotropy or mediation effects has not been examined. The analyses were well-powered for moderate and large effect sizes. Still, for unclear reasons, despite similar minor allele frequencies and imputation quality of the SNPs we tested across all ancestries, we identified no significant associations in the African American cohort. Finally, although our mouse studies detected some discordant cross-species effects of *Cadm2* on behavior, background strain effects [89] or subtle allelic variations (vs whole KO) may explain those differences. While some results are suggestive of additive effects, we were unable to evaluate different genetic models due to lack of sufficient sample sizes for HOM mice. Future multivariate analyses examining paths of commonality and specificity across impulsivity facets may provide further insights not herein examined.

In conclusion, we show that impulsivity facets are extremely polygenic, but of very high transdiagnostic significance. Genetic studies using research participants not ascertained for neuropsychiatric disorders may represent an efficient and cost-effective strategy for elucidating the genetic basis and etiology of genetically complex psychiatric diseases. Using homologous measures of impulsivity in mice and humans across three ancestral backgrounds, we provide evidence of the overarching role of *CADM2* on impulsivity, and a much broader impact on human health.

## Data Availability

We will provide summary statistics for the top 10,000 SNPs upon publication (Tables S22-30). Full GWAS summary statistics will be made available through 23andMe to qualified researchers under an agreement with 23andMe that protects the privacy of the 23andMe participants. Please visit https://research.23andme.com/collaborate/#dataset-access/ for more information and to apply to access the data.

## Data availability

We will provide summary statistics for the top 10,000 SNPs upon publication (**Tables S22-30**). Full GWAS summary statistics will be made available through 23andMe to qualified researchers under an agreement with 23andMe that protects the privacy of the 23andMe participants. Please visit https://research.23andme.com/collaborate/#dataset-access/ for more information and to apply to access the data.

## Acknowledgements

We would like to thank the research participants and employees of 23andMe for making this work possible. The following members of the 23andMe Research Team contributed to this study: Stella Aslibekyan, Adam Auton, Elizabeth Babalola, Robert K. Bell, Jessica Bielenberg, Katarzyna Bryc, Emily Bullis, Daniella Coker, Gabriel Cuellar Partida, Devika Dhamija, Sayantan Das, Teresa Filshtein, Kipper Fletez-Brant, Will Freyman, Karl Heilbron, Pooja M. Gandhi, Karl Heilbron, Barry Hicks, David A. Hinds, Ethan M. Jewett, Yunxuan Jiang, Katelyn Kukar, Keng-Han Lin, Maya Lowe, Jey C. McCreight, Matthew H. McIntyre, Steven J. Micheletti, Meghan E. Moreno, Joanna L. Mountain, Priyanka Nandakumar, Elizabeth S. Noblin, Jared O’Connell, Aaron A. Petrakovitz, G. David Poznik, Morgan Schumacher, Anjali J. Shastri, Janie F. Shelton, Jingchunzi Shi, Suyash Shringarpure, Vinh Tran, Joyce Y. Tung, Xin Wang, Wei Wang, Catherine H. Weldon, Peter Wilton, Alejandro Hernandez, Corinna Wong, Christophe Toukam Tchakouté.

We would also like to thank The Externalizing Consortium for sharing the GWAS summary statistics of externalizing. The Externalizing Consortium: Principal Investigators: Danielle M. Dick, Philipp Koellinger, K. Paige Harden, Abraham A. Palmer. Lead Analysts: Richard Karlsson Linnér, Travis T. Mallard, Peter B. Barr, Sandra Sanchez-Roige. Significant Contributors: Irwin D. Waldman. The Externalizing Consortium has been supported by the National Institute on Alcohol Abuse and Alcoholism (R01AA015416 -administrative supplement), and the National Institute on Drug Abuse (R01DA050721). Additional funding for investigator effort has been provided by K02AA018755, U10AA008401, P50AA022537, as well as a European Research Council Consolidator Grant (647648 EdGe to Koellinger). The content is solely the responsibility of the authors and does not necessarily represent the official views of the above funding bodies. The Externalizing Consortium would like to thank the following groups for making the research possible: 23andMe, Add Health, Vanderbilt University Medical Center’s BioVU, Collaborative Study on the Genetics of Alcoholism (COGA), the Psychiatric Genomics Consortium’s Substance Use Disorders working group, UK10K Consortium, UK Biobank, and Philadelphia Neurodevelopmental Cohort.

