## Supplementary Material for "*CADM2* is implicated in impulsive personality and numerous other traits by genome- and phenome-wide association studies in humans and mice"

**Contents:**

1. Human studies

1.1 Analysis of local ancestry.…...………………………………………………………….3

1.2 Genome-wide association and secondary analyses.………………………………...4

1.3 Genome-wide association analyses – Nominal Results……………………….…….5	1.4 Distribution of impulsivity scores………………………………………………………..7	1.5 Manhattan and QQ plots and regional association plots…………………………... 10 1.6 Genetic and phenotypic inter-correlations…………………………......................... 35 1.7 FDR-significant results for European and Latin American ancestries……………. 36 1.8 *Cadm2* expression………………………………………………………………………42

2. Mouse studies

2.1 Establishment of the Cadm2 mouse line…………………………………………….. 43

2.2 Western blotting……………………………………………………………………….... 44

2.3 Cadm2 protein expression results …………………………………………………... 45

2.4 Cohort 1…………………………………………………………………………………..46

2.4.1 Behavioral testing………………………………………………………….....46

2.4.10 Statistical analyses for cohort 1…………………………………………....51

2.4.11 Results for cohort 1……………………………………………………….....52

2.4.11.1 PRBT…………………………………………………………….....52

2.4.11.2 PRLT………………………………………………………………..52

2.4.11.3 IGT………………………………………………………………....52

2.4.11.4 BPM………………………………………………………………..53

2.4.11.5 PPI………………………………………………………………....54

2.5 Cohort 2…………………………………………………………………………………...56

2.5.1 Behavioral testing……………………………………………………………. 56

2.5.2 Behavioral sequence………………………………………………………... 56

2.5.3 Statistical analyses for Cohort 2………………………………………….... 59

2.5.4 Results for Cohort 2…………………………………………………………. 59

2.5.4.1 5CSRTT……………………………………………………………. 59

2.6 Cohort 3…………………………………………………………………………………...63

2.6.1 Behavioral testing……………………………………………………………. 63

2.6.2 Open field testing and apparatus…………………………………………... 63

2.6.3 Elevated plus maze, Light-dark box and apparatus……………………... 63

2.6.4 Drinking-in-the-dark procedure…………………………………………….. 64

2.6.5 Statistical analyses for cohort 3……………………………………………. 65

2.6.6 Results for Cohort 3…………………………………………………………. 65

2.6.6.1 OFT, EPM, LDB…………………………………………………....65

2.6.6.2 DID procedure………………………………………………………66

2.7 Cohort 4…………………………………………………………………………………...68

2.7.1 Body weight measurements………………………………………………....68

2.8 Cohort 5…………………………………………………………………………………..69

2.8.1 Dendritic spine analysis………………………………………………………69

2.8.2 Statistics for cohort 5………………………………………………………....69

2.8.3 Results for cohort 5……………………………………………………………70

2.8.3.1 Dendritic spine density……………………………………………………..70

*Human studies*

**Analysis of local ancestry**

Ancestry was determined by analyzing local ancestry [1]. Briefly, the 23andMe algorithm first partitions phased genomic data into short windows of about 300 SNPs. Within each window, we use a support vector machine to classify individual haplotypes into one of 31 reference populations ([https://www.23andme.com/ancestry-composition-guide/](https://urldefense.com/v3/__https://www.23andme.com/ancestry-composition-guide/__;!!LLK065n_VXAQ!z5rFfTqrdN6q6vGu1ot2ECJk_285VG_A7OkNusXVPeuiLsCpyVVLc647SpftxX9s-vMbXQ$)). The support vector machine classifications are fed into a hidden Markov model that accounts for switch errors and incorrect assignments, and gives probabilities for each reference population in each window. Finally, we used simulated admixed individuals to recalibrate the hidden Markov model probabilities so that the reported assignments are consistent with the simulated admixture proportions. The reference population data is derived from public datasets (the Human Genome Diversity Project, HapMap, and 1000 Genomes), as well as 23andMe customers who have reported having four grandparents from the same country.

| **Ancestry** | **Classification criteria** |
| --- | --- |
| European | European + Middle Eastern > 0.97, European > 0.90 |
| East Asian | East Asian + Southeast Asian > 0.97 |
| South Asian | South Asian > 0.97 |
| Middle Eastern (& North African) | Middle Eastern + European > 0.97, Middle Eastern > 0.90 |
| African American + Latin American | European + African + East Asian + Native American + Middle Eastern > 0.90, African + Native American > 0.01 |

Ancestries are defined as follows:

African Americans and Latin American are admixed with broadly varying contributions from Europe, Africa and the Americas. The distributions of the length of segments of European, African and American ancestry are very different between African Americans and Latin American because of distinct admixture timing between the three ancestral populations in the two ethnic groups. Therefore, we trained a logistic classifier that takes one individual's length histogram of segments of African, European and American ancestry and predicts whether the customer is likely African American or Latin American.

**Genome-wide association and secondary analyses**

Participants were genotyped on one of five Illumina genotyping platforms, containing between 550,000 to 950,000 variants, for a total of 1.6 million genotyped variants. Samples that failed to reach 98.5% call rate were re-analyzed. Genotyping quality controls included discarding variants with a Hardy-Weinberg *p*<1.00E−20, batch effects (ANOVA *p*<1.00E-20), or a call rate of <90% (33,35,36). About 64.4M variants were then imputed against the Haplotype Reference Consortium (**HRC**) panel, augmented by a single unified imputation reference panel combining the May 2015 release of the 1000 Genomes Phase 3 haplotypes with the UK10K imputation reference panel, for variants not present in the HRC. Imputed variants with low imputation quality (r^2^<0.5 averaged across batches or a minimum r^2^<0.3), or with evidence of batch effects (*p*<1.00E-50) were removed (33,35). A total of 1.3M genotyped and 30.5M imputed variants passed the pre- and post GWAS quality controls. We furthermore filtered out variants with minor allele frequency (**MAF**) <0.1%, which are extremely sensitive to quantitative trait over-dispersion, reducing to 14.1M variants available for follow-up analyses. Principal components were computed using ~65,000 high-quality genotyped variants present in all five genotyping platforms.

A maximal set of unrelated individuals was chosen for the analysis using a segmental identity-by-descent (**IBD**) estimation algorithm [2] to ensure that only unrelated individuals were included in the sample. Individuals were defined as related if they shared more than 700 cM IBD, including regions where the two individuals shared either one or both genomic segments IBD. This level of relatedness (~20% of the genome) corresponds to approximately the minimal expected sharing between first cousins in an outbred population.

We imputed participant genotype data against the September 2013 release of 1000 Genomes phase 1 version 3 reference haplotypes. We phased and imputed data for each genotyping platform separately. We phased using an internally developed phasing tool, Finch, which implements the Beagle haplotype graph-based phasing algorithm, modified to separate the haplotype graph construction and phasing steps. In preparation for imputation, we split phased chromosomes into segments of no more than 10,000 genotyped SNPs, with overlaps of 200 SNPs. We excluded SNPs with Hardy-Weinberg equilibrium *P* < 10^−20^, call rate < 95%, or with large allele frequency discrepancies compared to European 1000 Genomes reference data. Frequency discrepancies were identified by computing a 2 x 2 table of allele counts for European 1000 Genomes samples and 2000 randomly sampled 23andMe customers with European ancestry, and identifying SNPs with a 𝜒^2^ *P* < 10^−15^. We imputed each phased segment against all-ethnicity 1000 Genomes haplotypes (excluding monomorphic and singleton sites) using Minimac2[3], using 5 rounds and 200 states for parameter estimation.

For the X chromosome, we built separate haplotype graphs for the non-pseudoautosomal region and each pseudoautosomal region, and these regions were phased separately. We then imputed males and females together using Minimac2, as with the autosomes, treating males as homozygous pseudo-diploids for the non-pseudoautosomal region.

For tests using imputed data, we use the imputed dosages rather than best-guess genotypes. We imputed HLA allele dosages from SNP genotype data using HIBAG. We imputed alleles for HLA-A, B, C, DPB1, DQA1, DQB1, and DRB1 loci at four-digit resolution. To test associations between HLA allele dosages and phenotypes, we performed linear regression using the same set of covariates used in the SNP based GWAS. We performed separate association tests for each imputed allele. HLA analysis did not identify any significant signal and hence are not described in the main text.

MAGMA was used for gene-based analyses, which uses Ensembl (build 85) to map SNPs to 19,773 protein-coding genes. We used a Bonferroni correction based on the number of genes tested (*p*<2.53E-06).

We followed-up on one of the most robust signals (i.e., *CADM2* locus) using single-SNP PheWAS. Because the *CADM2* locus is located in a dense LD region, and considering that previous studies have identified various SNPs (rs993137, rs6803322, rs818219, rs62263923, rs11708632) to be implicated in multiple unrelated traits (e.g., risk, educational attainment, cognition, schizophrenia, BMI, systolic blood pressure, neuroticism, anxiety), this made us hypothesize that there may be independent loci underlying these traits. To gain more information about the pleiotropic effect of these various lead SNPs (and not the broad polygenic architecture) we performed single-SNP PheWASs of some of the most implicated variants in or near *CADM2* based on prior literature (as summarized in **Supplementary Table 4**). The SNPs selected showed a moderate to low degree of correlation (European: **Supplementary Figure 23**; Latin American: **Supplementary Figure 35**; African American: only rs993137 and rs818219 showed some degree of correlation, R^2^=0.29). We excluded traits with <1,000 responses (11.99%, 3.42%, and 6.61% from the European, Latin American, and African American cohorts). For case-control comparisons, we computed association test results by logistic regression. For quantitative and categorical traits, association tests were performed by linear regression. There was some degree of overlap between the discovery GWAS with that of the PheWAS; however, we do not think this is unusual or problematic. For example, our observation of pleiotropy in the primary GWAS reflects almost complete sample overlap, which is common in the scientific literature. In contrast, the single-SNP PheWAS that we present is only partially overlapping. While it is true that other techniques (e.g., polygenic risk scores) can suffer when there is sample overlap, we do not think single-SNP PheWAS suffers from these limitations.

**Genome-wide association analyses – Nominal Results**

*GWAS of UPPS-P*

*Premeditation.* We noted two interesting nominal associations: rs774880622 (*p*=2.10E-07) on chromosome 11, near the *DRD2* gene, which has been extensively studied in relation to reward and is a candidate gene for many psychiatric disorders, particularly substance use disorders [e.g., [4]]; and rs72819189 (*p*=9.70E-07), on chromosome 2, near the *AFF3* gene, which was a robust candidate gene associated with externalizing psychopathology [5].

*Perseverance.* We noted a nominal association with rs10401120 (*p*=6.80E-07), in the *TCF4* gene.

*Negative Urgency.* We note nominal associations near the corticotropin receptor gene (*CRHR1*; rs2532373, *p*=9.10E-08), which is a robust candidate gene in relation to stress, depression and anxiety disorders; with *CDH13* (rs426583, *p*=8.10E-07), which has been previously associated with methamphetamine response in both humans [6] and rats [7]; and with *NRXN1* (rs10651842, *p*=7.10E-07), which has previously been associated with educational attainment [8] and BMI [9].

*GWAS of BIS-11*

*Attentional.* We also detected a nominal association (rs145225651, *p*=5.00E-07), which is near the gene *CADM2*. Also of note is a nominal association within *PABPC4* (rs1601647, *p*=6.30E-07), on chromosome 4, which was also nominally associated with Nonplanning, but novel in relation to impulsivity.

*Nonplanning*. Again, we detected a nominal association within *PABPC4* (rs1601647, *p*=5.30E-07); with *AFF3* (rs72819158, *p*=1.10E-07), a robust candidate gene associated with externalizing [5]; and within *FOXP2* (rs936146, *p*=5.50E-07), a robust candidate for cannabis use disorders [10], smoking initiation [4], general risk tolerance [11], cognitive ability, years of educational attainment and schizophrenia [12].

*GWAS of Drug Experimentation*

We noted a nominal association near the gene encoding the Glutamate Metabotropic Receptor 3 (*GRM3*; rs12673181, *p*=4.50E-07). Phenotypes associated with *GRM3* include educational attainment [8], schizophrenia [86], and neuroticism [87]. We also identified a nominal association near *HTR3B* (rs6589400, p=2.80E-07), which encodes the serotonin receptor 3B and is implicated in various forms of impulsivity and the reward system [88].

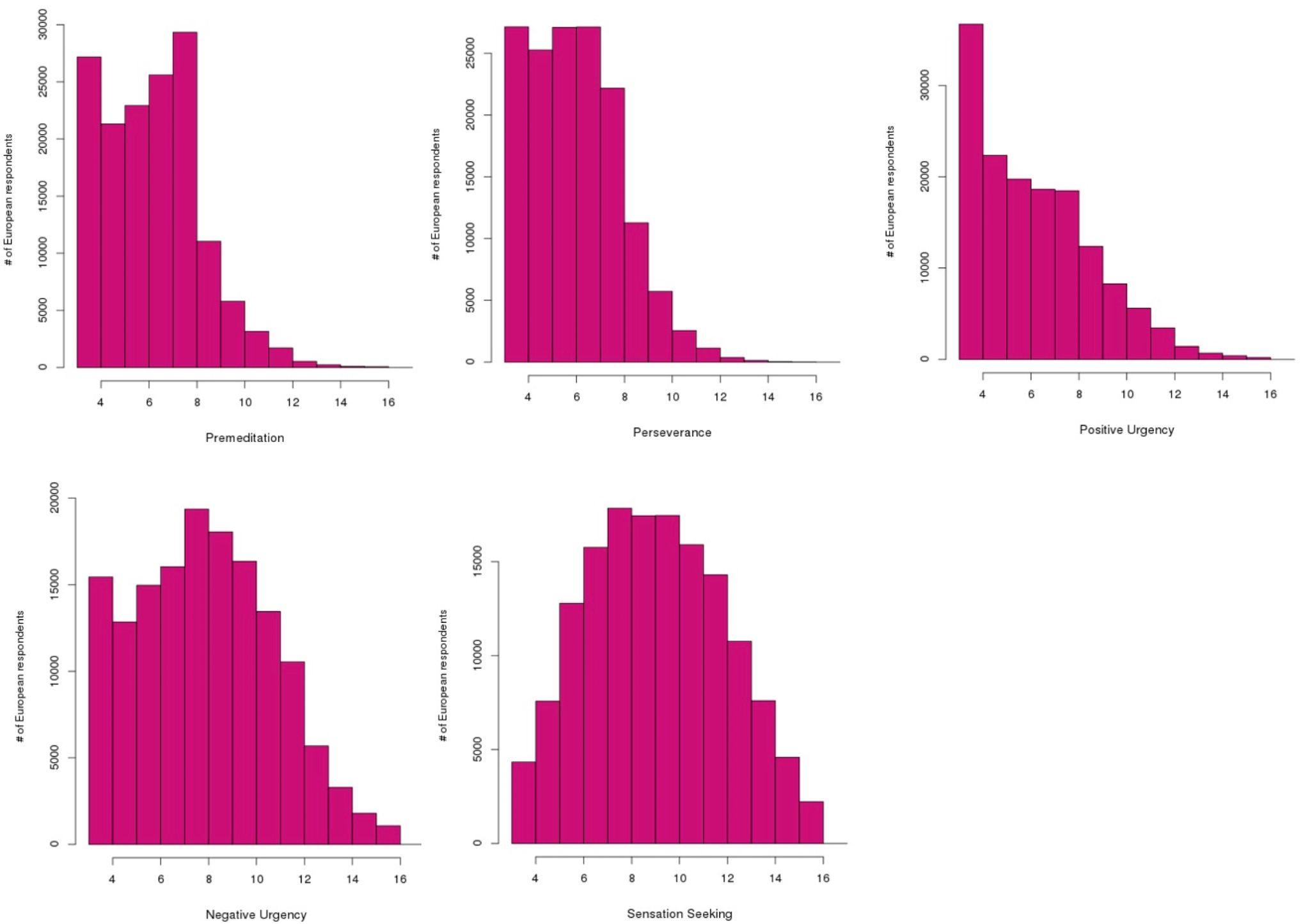

**Figure S1. Distributions for UPPSP.** Distribution (%) of UPPS-P scores (European subjects). Low scores indicate lower rates of Premeditation, Perseverance, Positive Urgency, Negative Urgency, and Sensation Seeking. Each row corresponds to a particular range. Each subscale includes 4-items and yields integer scores from 4 to 16.

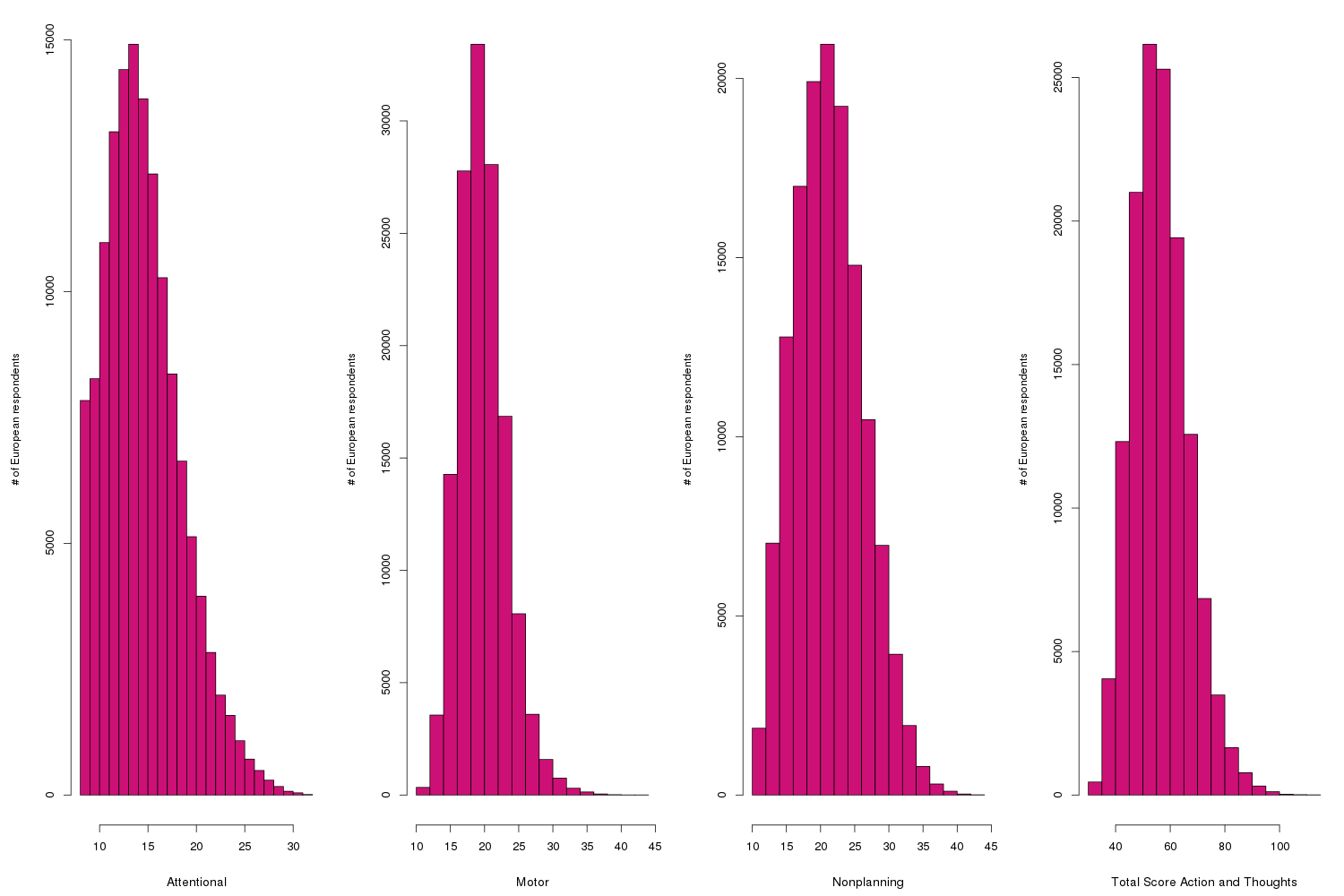

**Figure S1. Distributions for BIS.** Distribution (%) of BIS-11 scores (European subjects). Low scores indicate lower rates of self-reported Attentional, Motor and Nonplanning impulsivity as measured by BIS-11. Each row corresponds to a particular range.

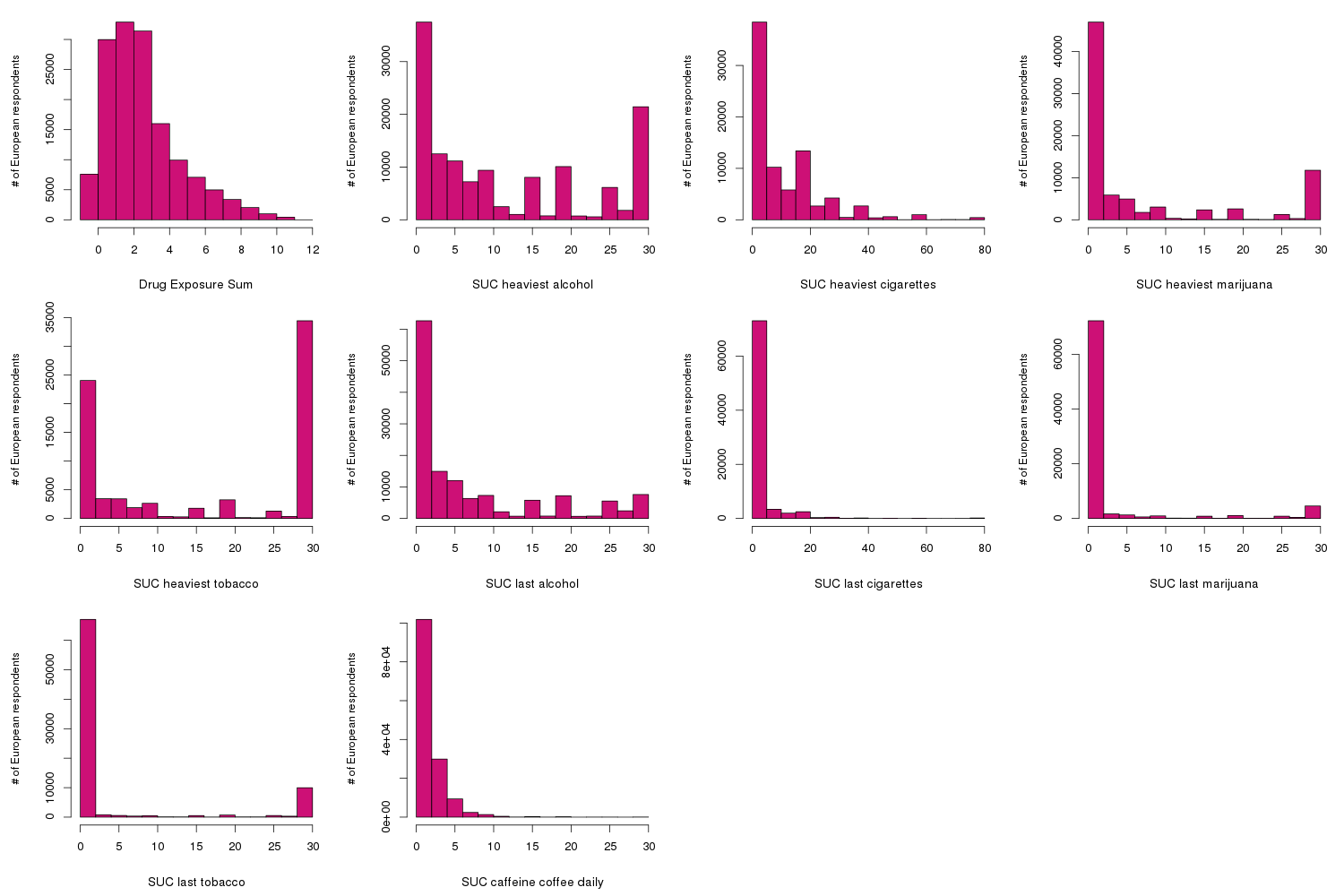

**Figure S3. Distribution for Drug Experimentation.** Distribution (%) of Drug Experimentation scores (European subjects). Each row corresponds to a particular range. To calculate Drug Experimentation one point was given for each drug the subject endorsed having tried: alcohol, marijuana, cocaine, methamphetamines, LSD/magic mushrooms, ecstasy, prescription stimulants (taken not as prescribed), prescription painkillers (taken not as prescribed), heroin, opium.

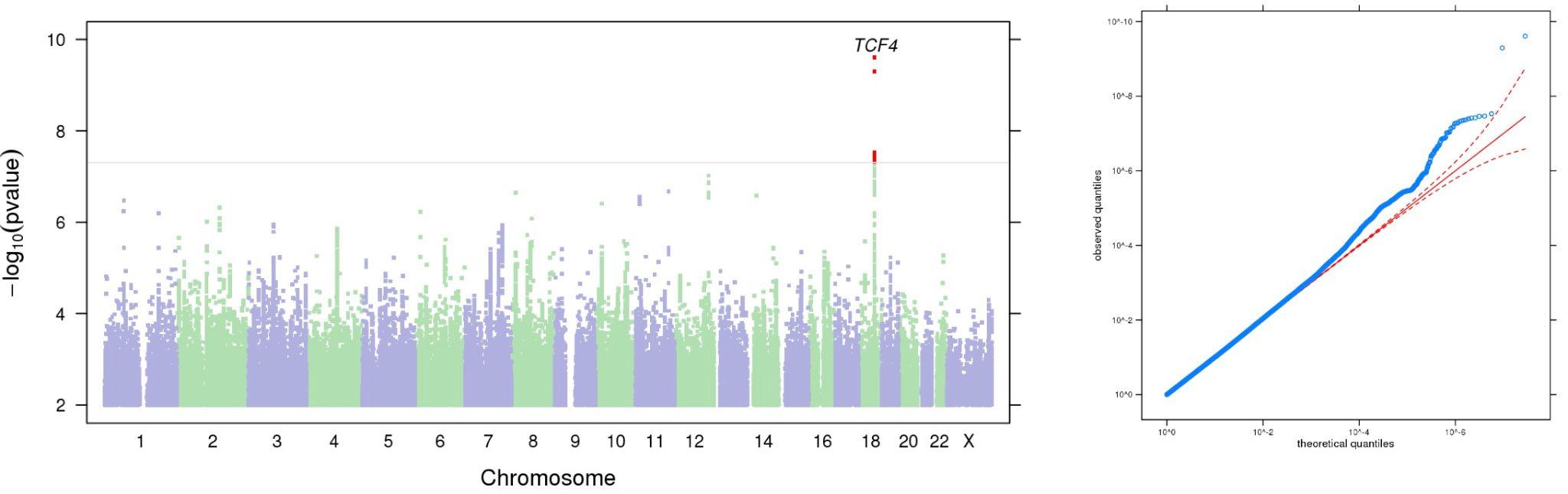

**Figure S4.** Manhattan and QQ plots of GWAS results indicating the strongest associations between the 22 autosomes, X chromosome, and **UPPS-P Premeditation**. The results have been adjusted for a genomic control inflation factor λ=1.085 (sample size = 132,667).

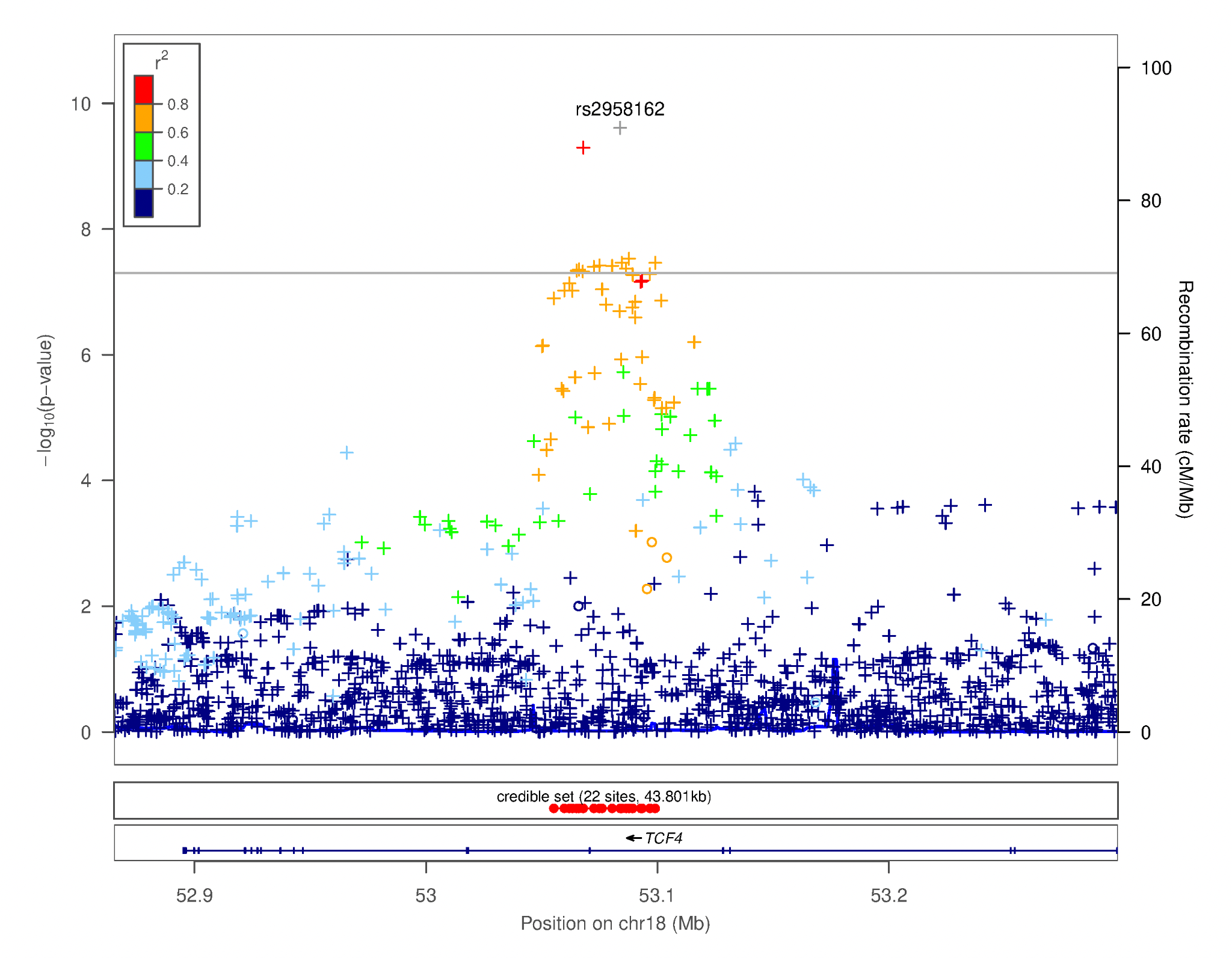

**Figure S5.** Regional association plot focusing on genetic variants associated with **UPPS-P Premeditation**. This plot was generated using LocusZoom [16]. The -log_10_(*p-*value) is shown on the left *y****-***axis; position in Mb is on the *x***-**axis. Recombination rates (expressed in centiMorgans cM per Mb; NCBI Build GRCh37; highlighted in blue) are shown on the right *y****-***axis. Pairwise linkage disequilibrium (r^2^) of each SNP with the top SNP in the region is indicated by its color. Crossed points represent imputed SNPs, circles represent directly genotyped SNPs.

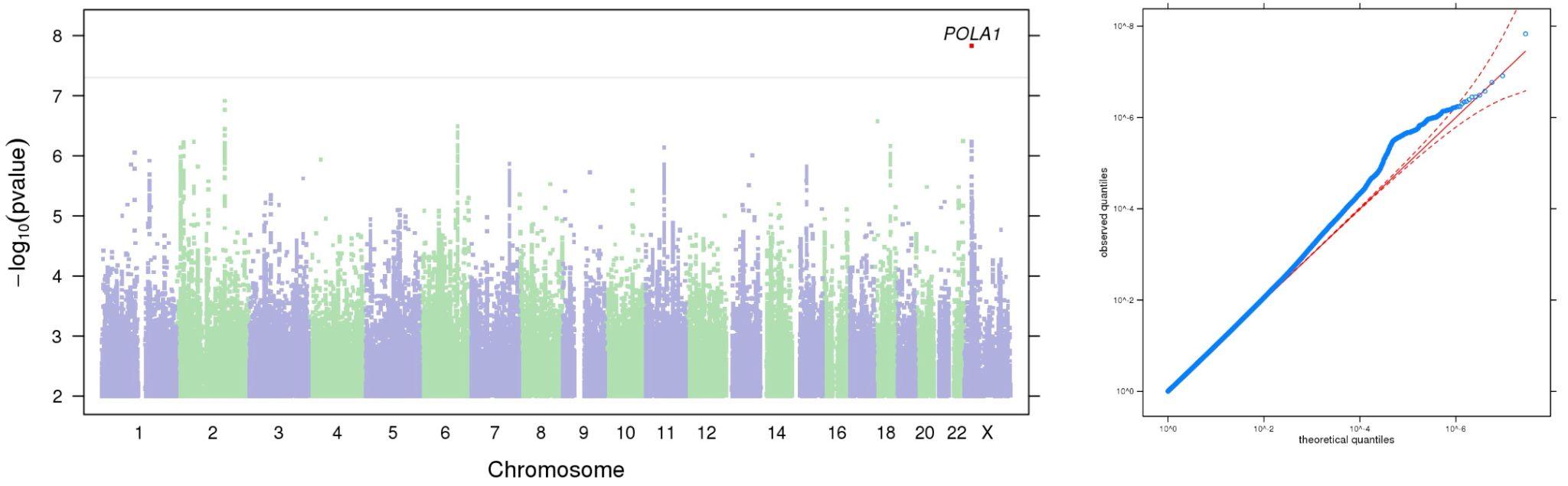

**Figure S6.** Manhattan and QQ plots of GWAS results indicating the strongest associations between the 22 autosomes, X chromosome, and **UPPS-P Perseverance**. The results have been adjusted for a genomic control inflation factor λ=1.089 (sample size = 133,517).

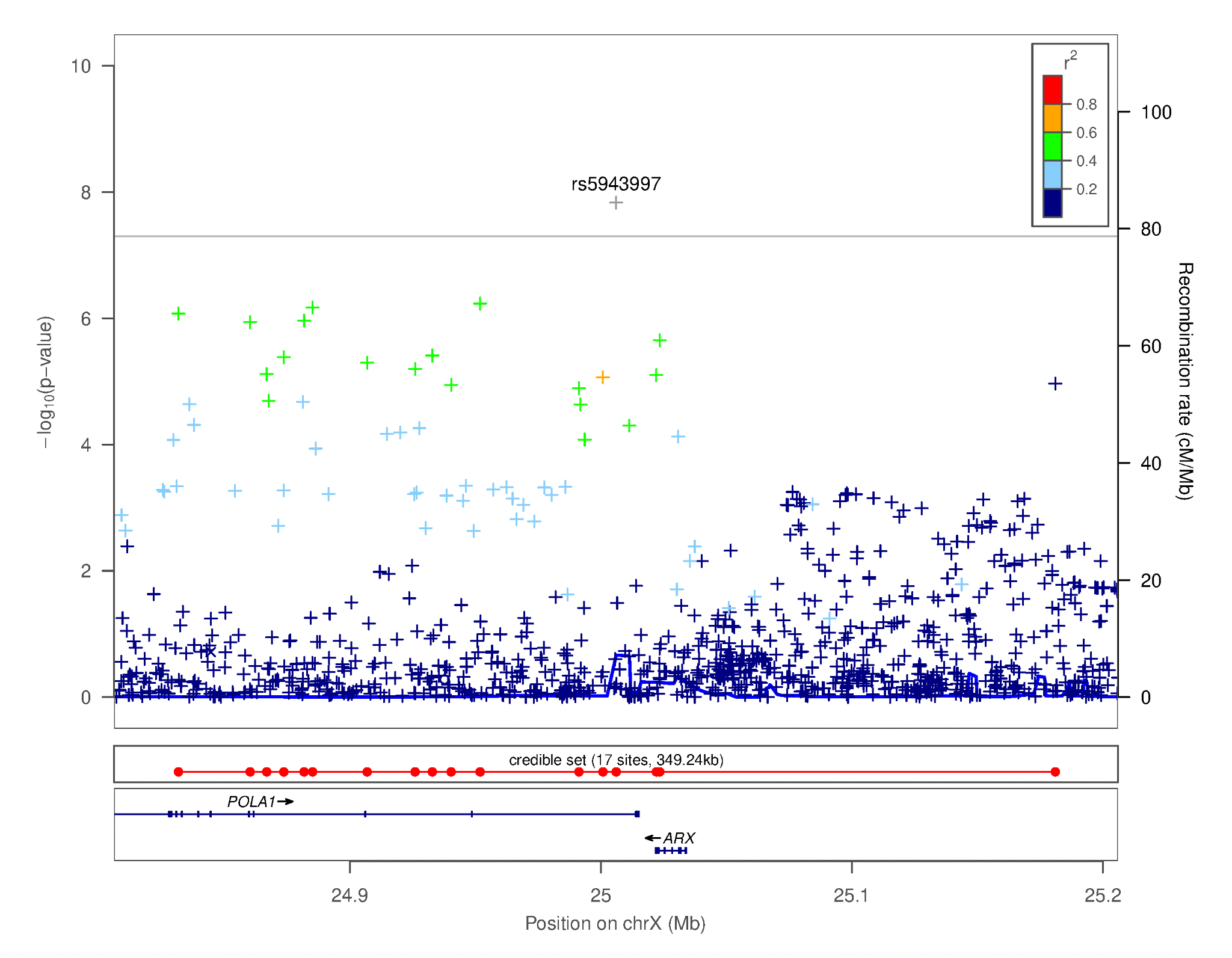

**Figure S7.** Regional association plot focusing on genetic variants associated with **UPPS-P Perseverance**. This plot was generated using LocusZoom [16]. The -log_10_(*p-*value) is shown on the left *y****-***axis; position in Mb is on the *x***-**axis. Recombination rates (expressed in centiMorgans cM per Mb; NCBI Build GRCh37; highlighted in blue) are shown on the right *y****-***axis. Pairwise linkage disequilibrium (r^2^) of each SNP with the top SNP in the region is indicated by its color. Crossed points represent imputed SNPs, circles represent directly genotyped SNPs.

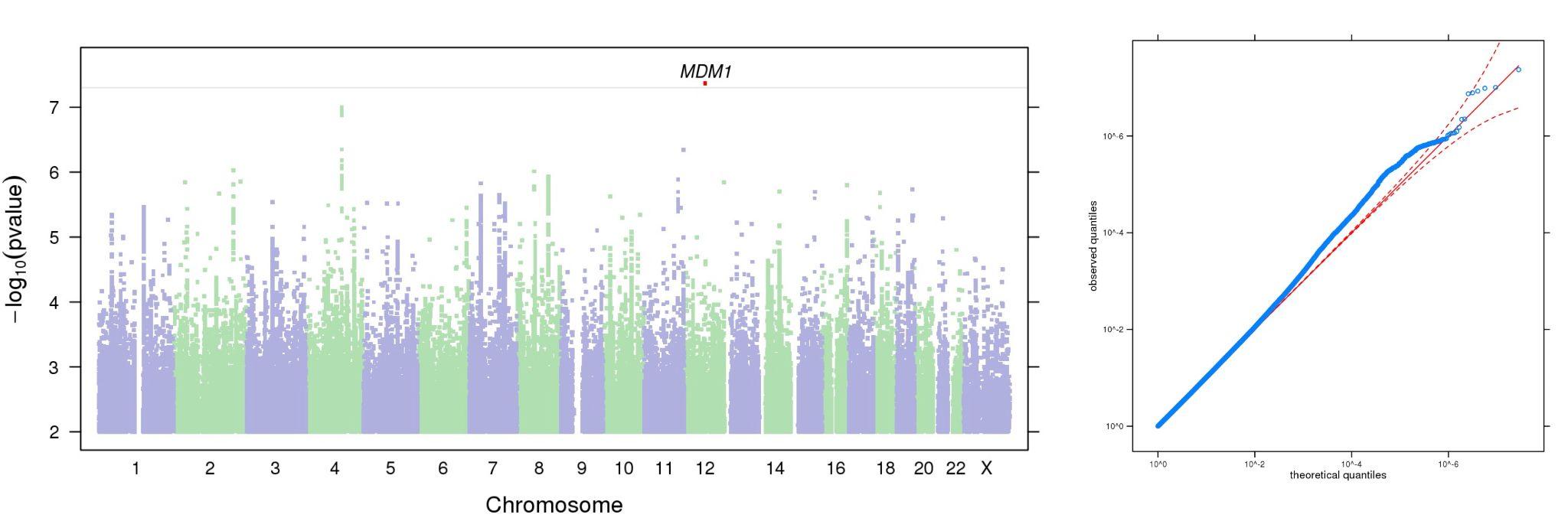

**Figure S8.** Manhattan and QQ plots of GWAS results indicating the strongest associations between the 22 autosomes, X chromosome, and **UPPS-P Positive Urgency**. The results have been adjusted for a genomic control inflation factor λ=1.001 (sample size = 132,132).

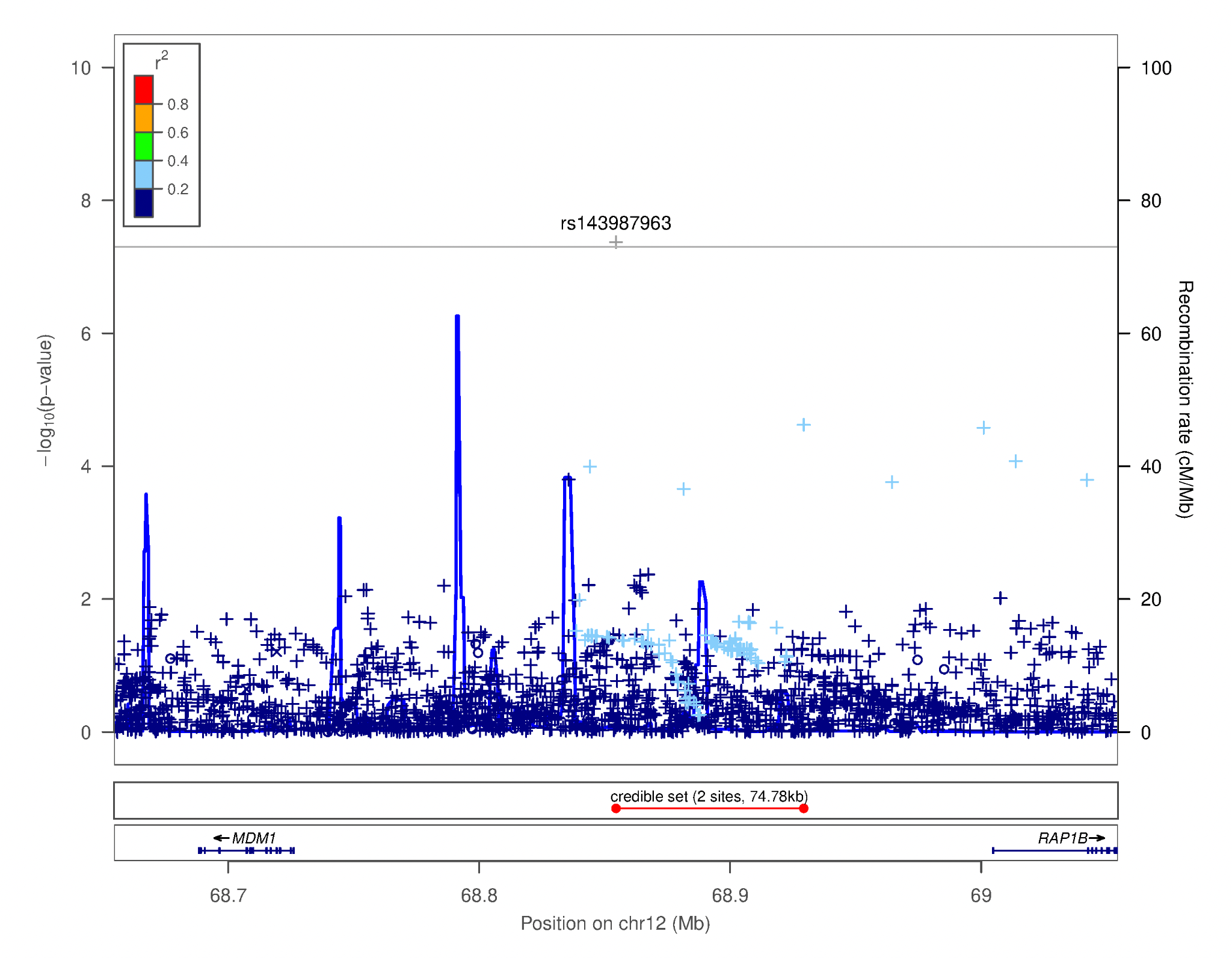

**Figure S9.** Regional association plot focusing on genetic variants associated with **UPPS-P Positive Urgency**. This plot was generated using LocusZoom [16]. The -log_10_(*p-*value) is shown on the left *y****-***axis; position in Mb is on the *x***-**axis. Recombination rates (expressed in centiMorgans cM per Mb; NCBI Build GRCh37; highlighted in blue) are shown on the right *y****-***axis. Pairwise linkage disequilibrium (r^2^) of each SNP with the top SNP in the region is indicated by its color. Crossed points represent imputed SNPs, circles represent directly genotyped SNPs. Inspection of this LocusZoom plot is not supportive of a robust signal.

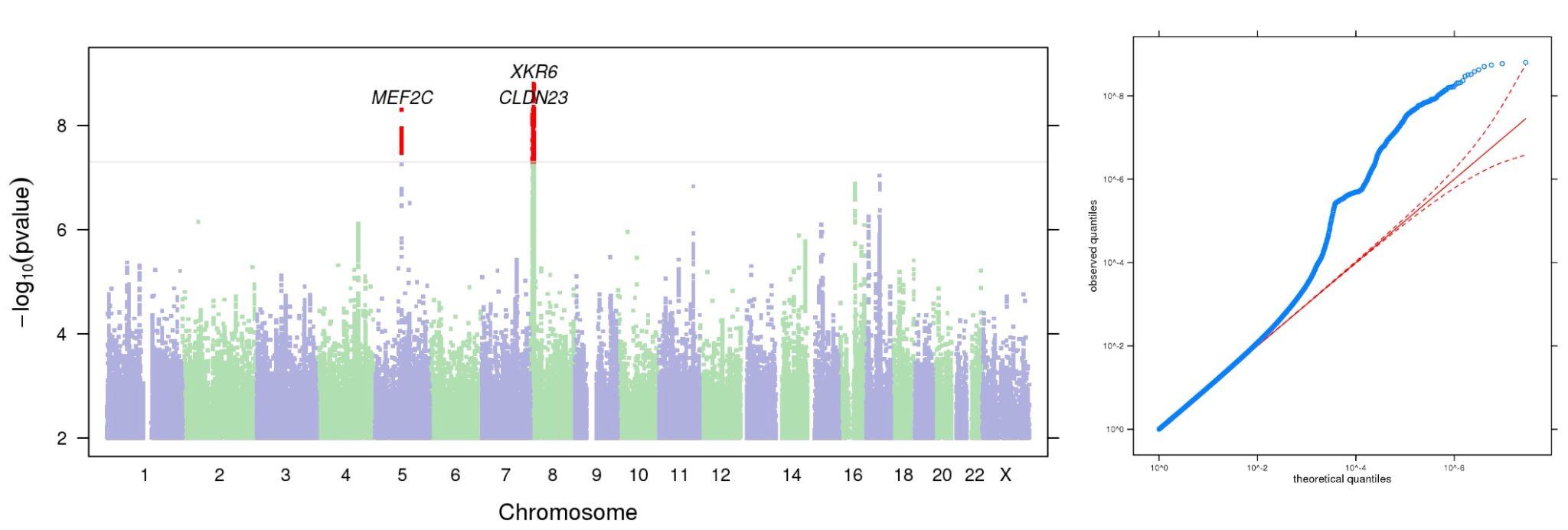

**Figure S10.** Manhattan and QQ plots of GWAS results indicating the strongest associations between the 22 autosomes, X chromosome, and **UPPS-P Negative Urgency**. The results have been adjusted for a genomic control inflation factor λ=1.002 (sample size = 132,559).

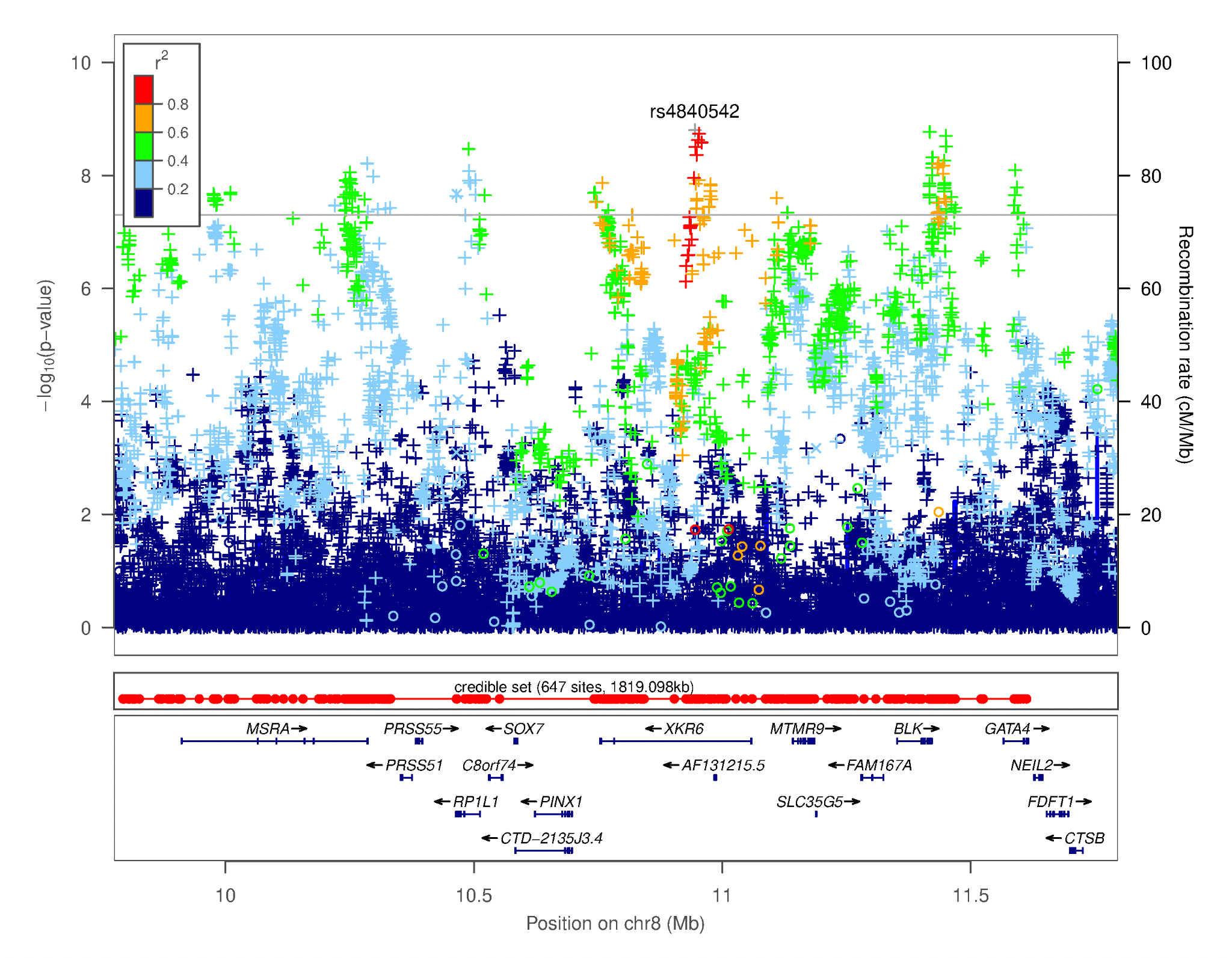

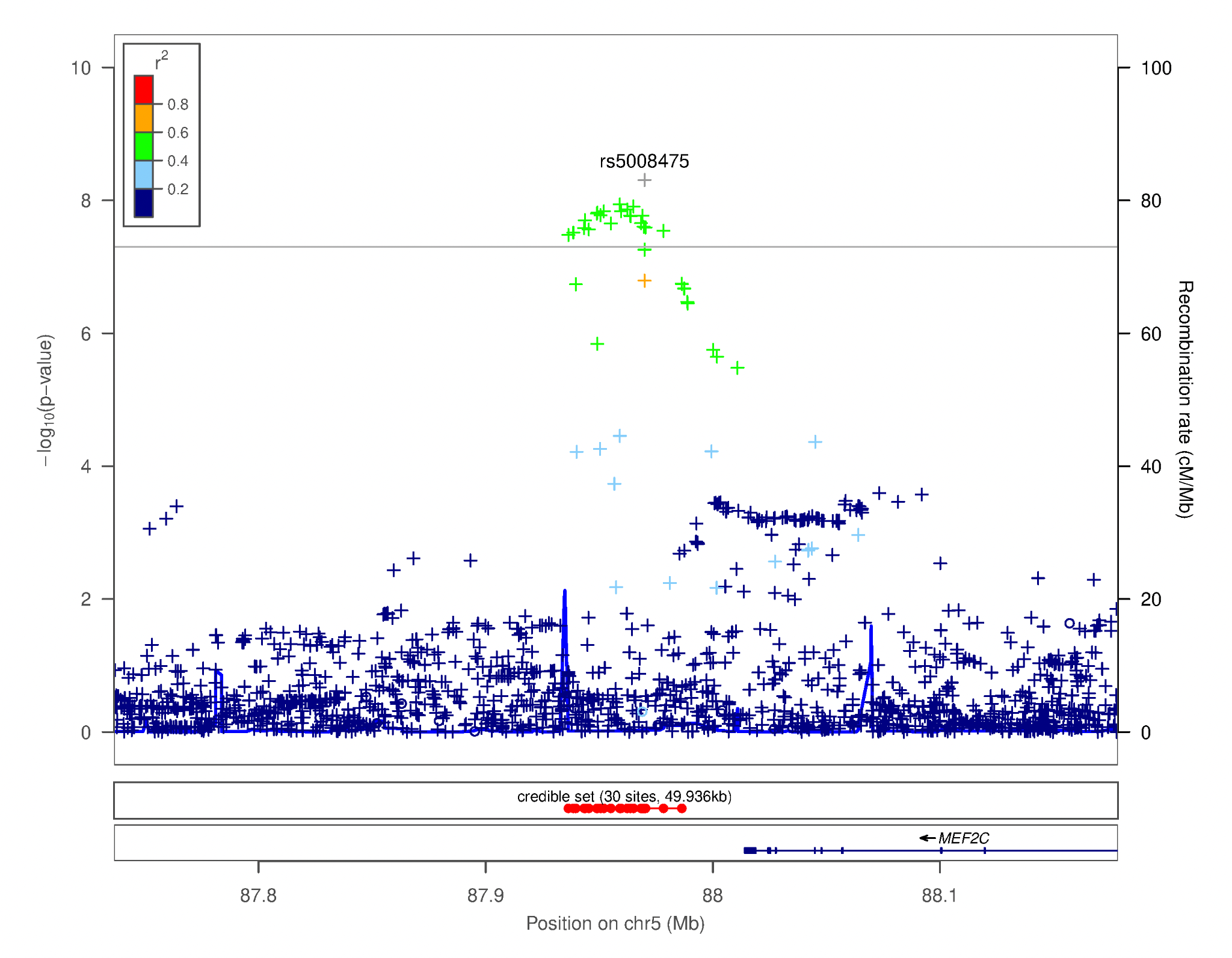

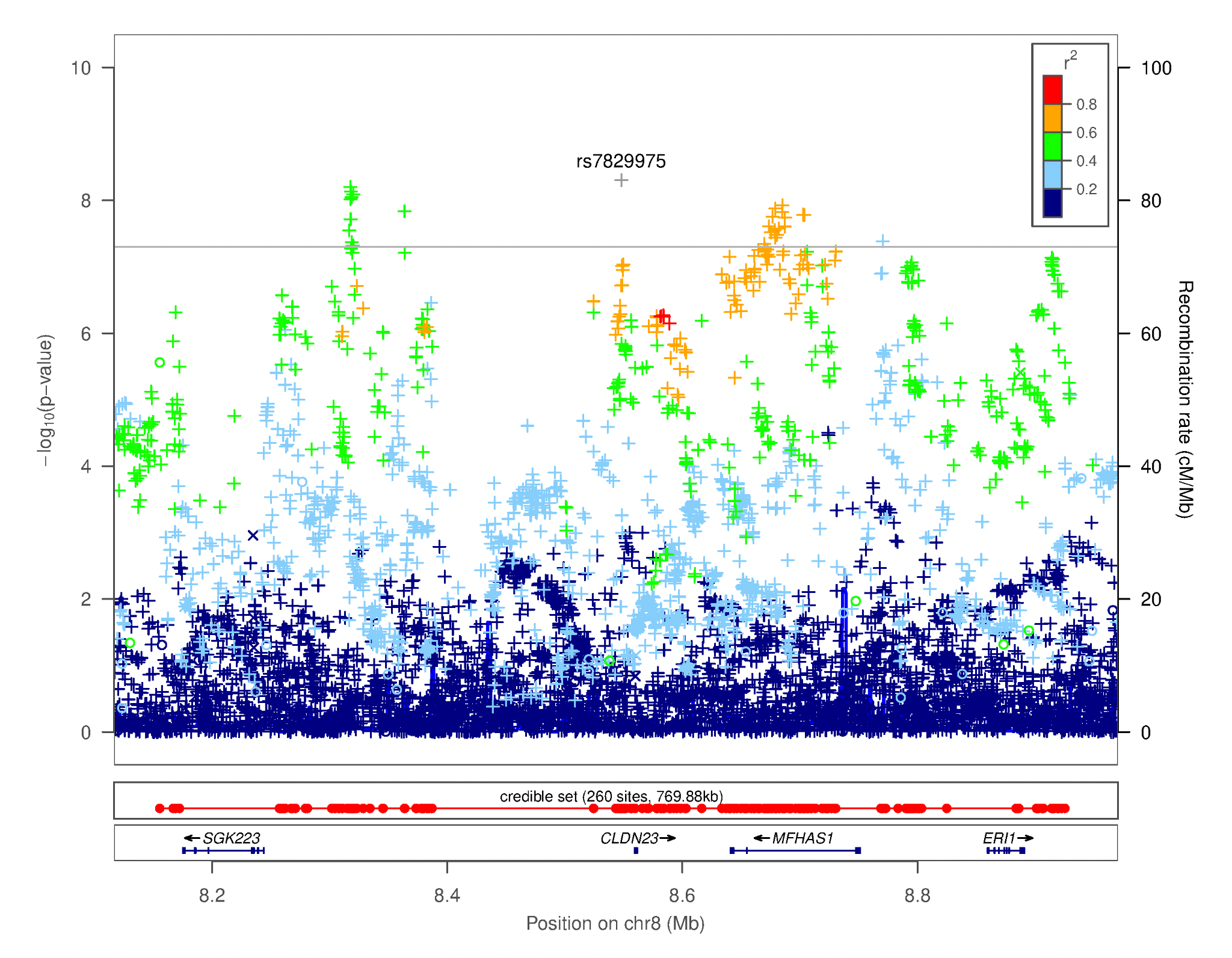

**Figure S11.** Regional association plots focusing on genetic variants associated with UPPSP Negative Urgency. This plot was generated using LocusZoom [16]. The -log_10_(*p*-value) is shown on the left *y****-***axis; position in Mb is on the *x***-**axis. Recombination rates (expressed in centiMorgans cM per Mb; NCBI Build GRCh37; highlighted in blue) are shown on the right *y****-***axis. Pairwise linkage disequilibrium (r^2^) of each SNP with the top SNP in the region is indicated by its color. Crossed points represent imputed SNPs, circles represent directly genotyped SNPs. Some of these associations (e.g., rs4840542, rs7829975) could be an artifact and therefore need to be further validated.

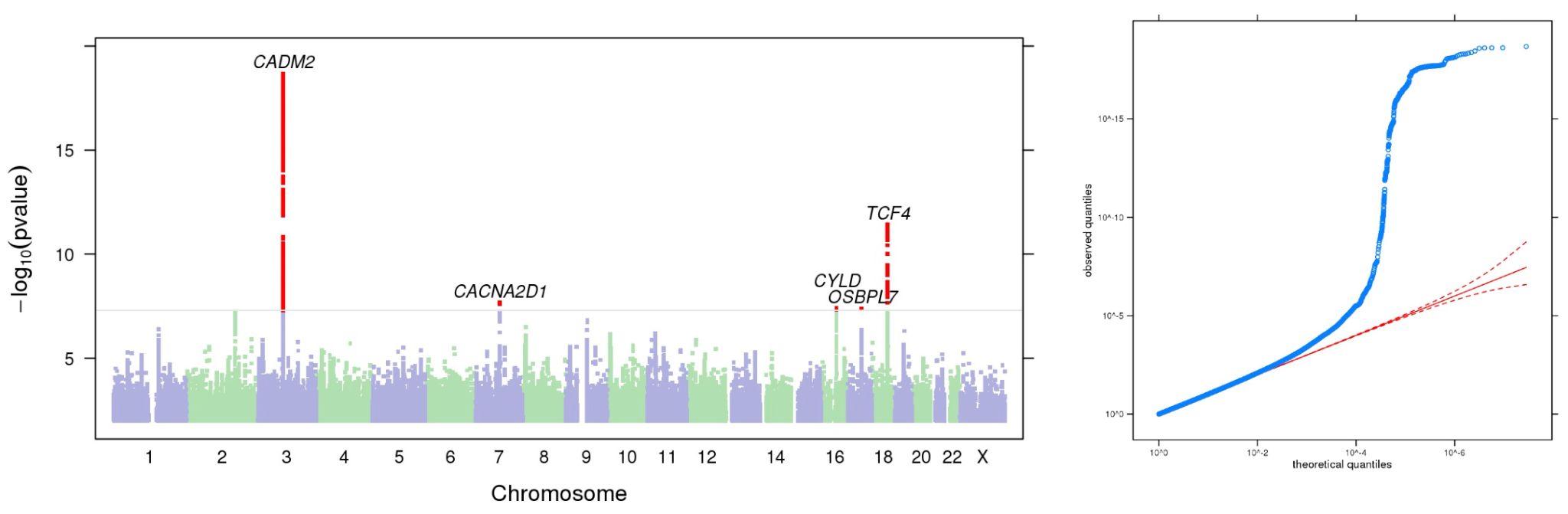

**Figure S12.** Manhattan and QQ plots of GWAS results indicating the strongest associations between the 22 autosomes, X chromosome, and **UPPS-P Sensation Seeking**. The results have been adjusted for a genomic control inflation factor λ=1.002 (sample size = 132,395).

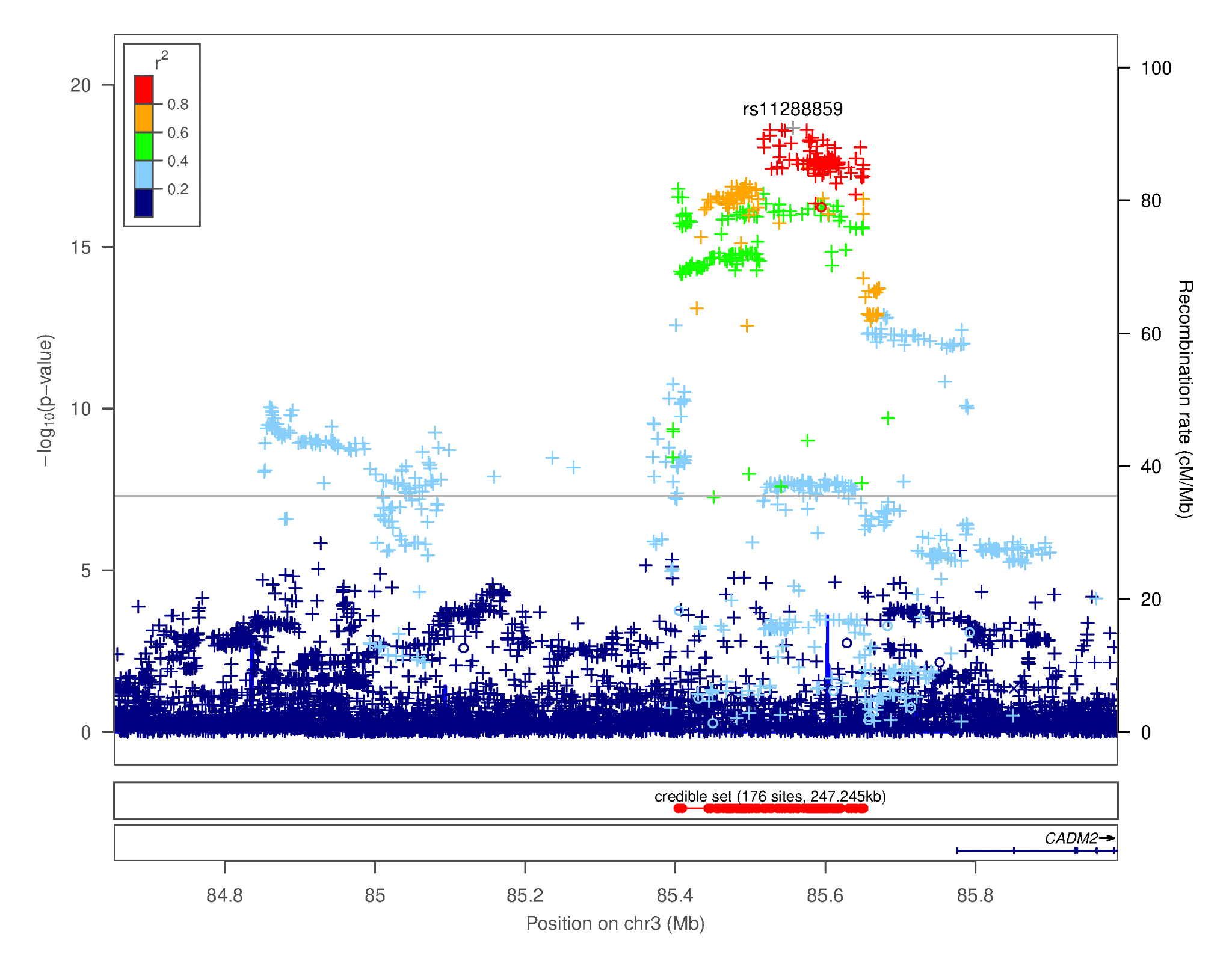

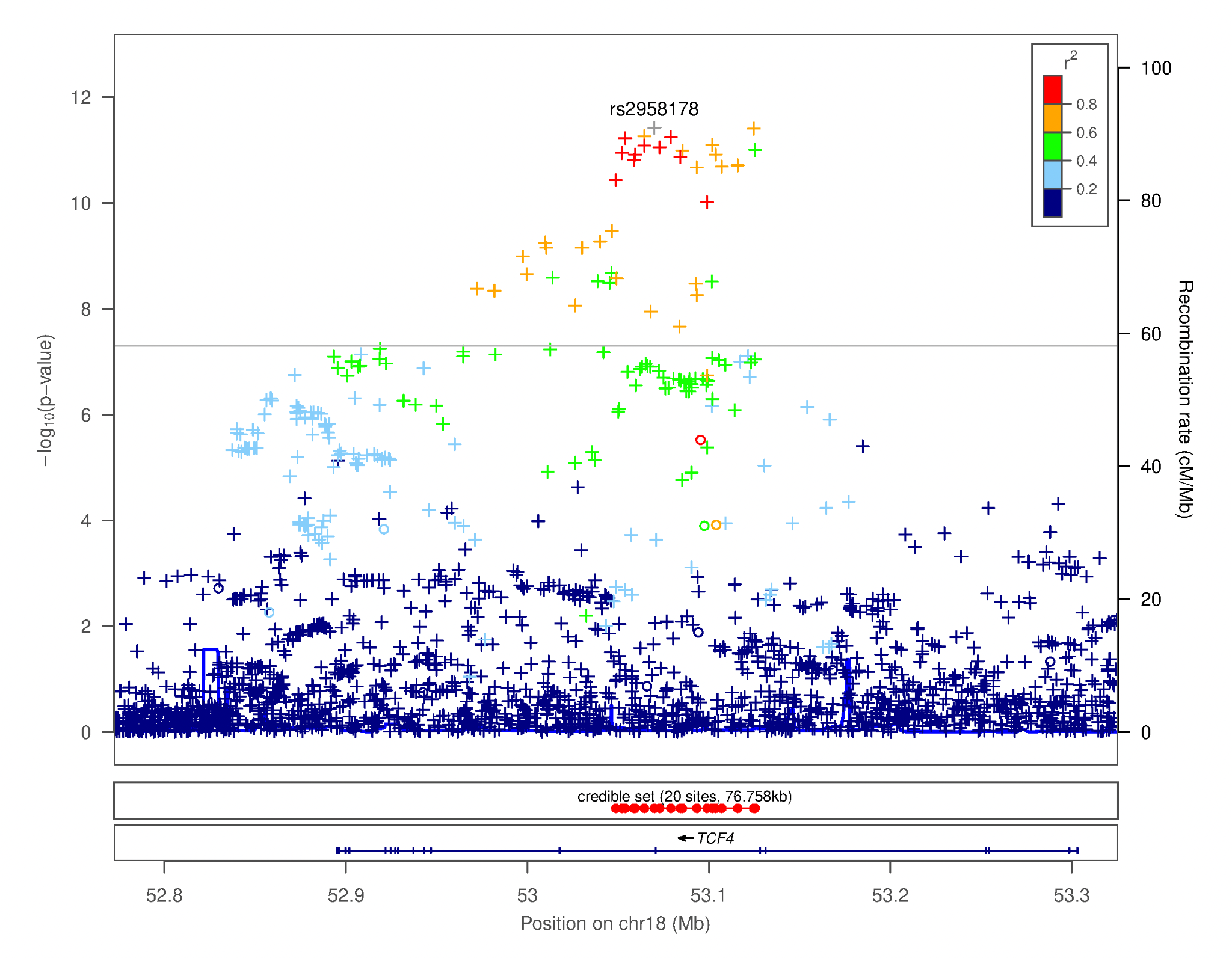

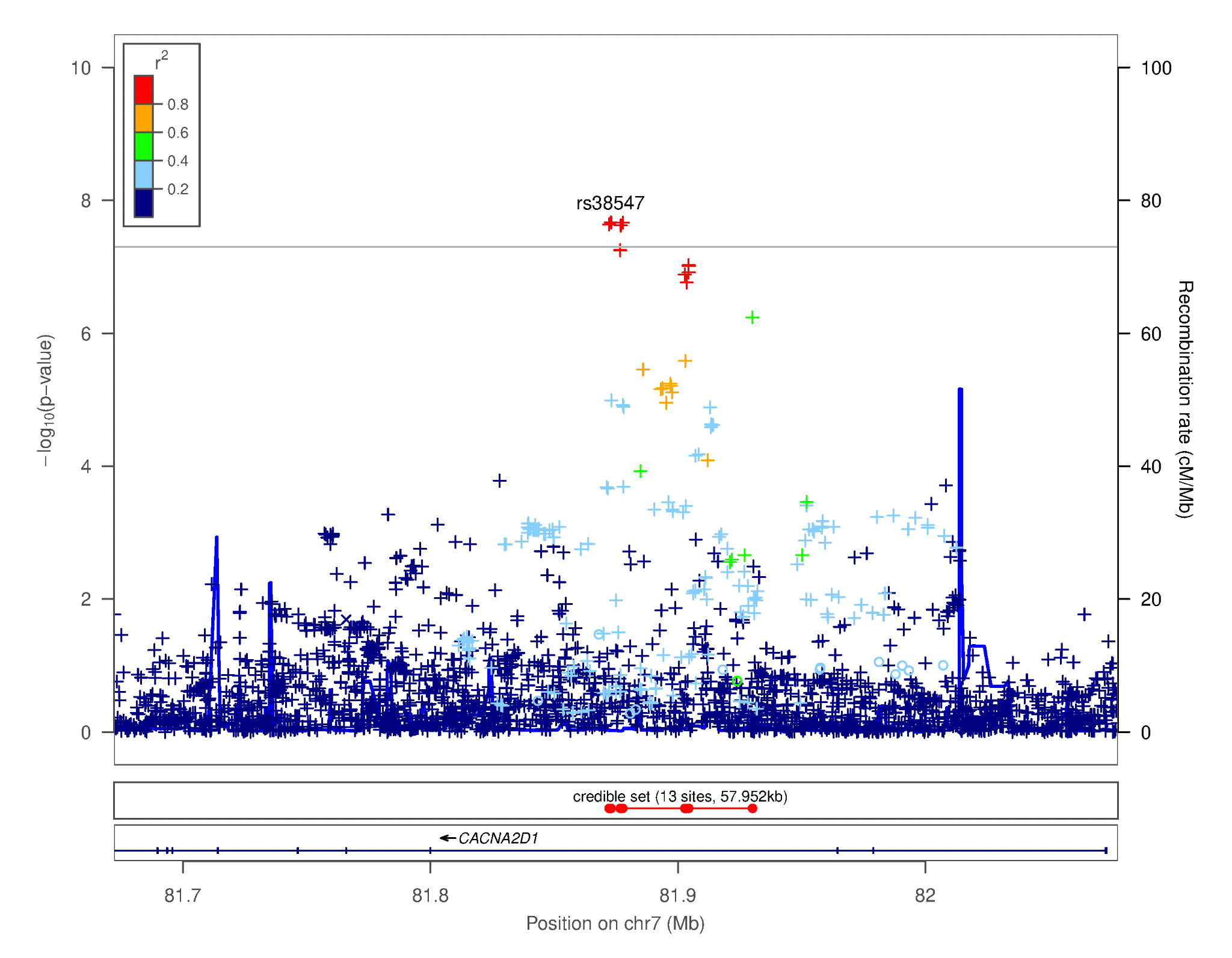

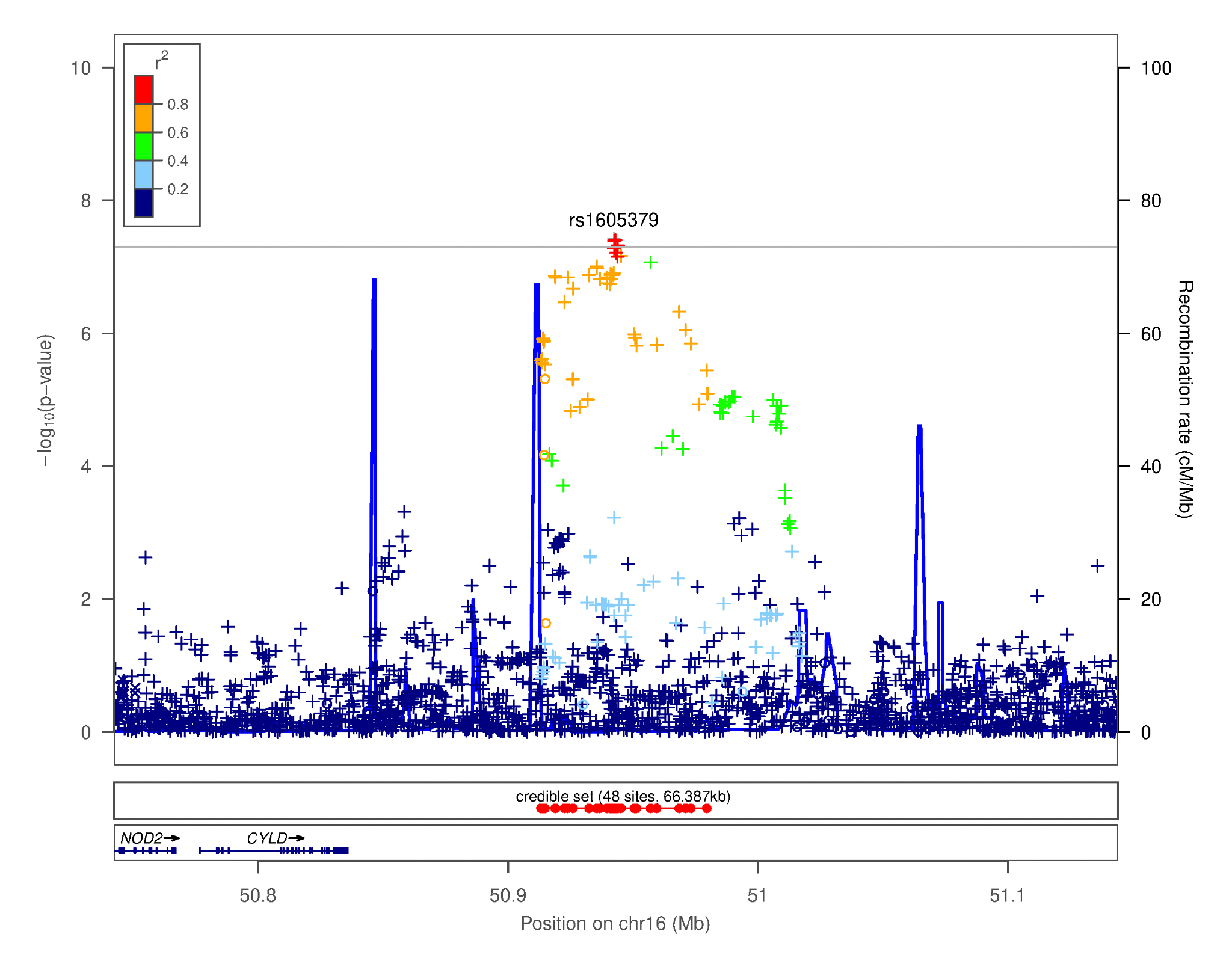

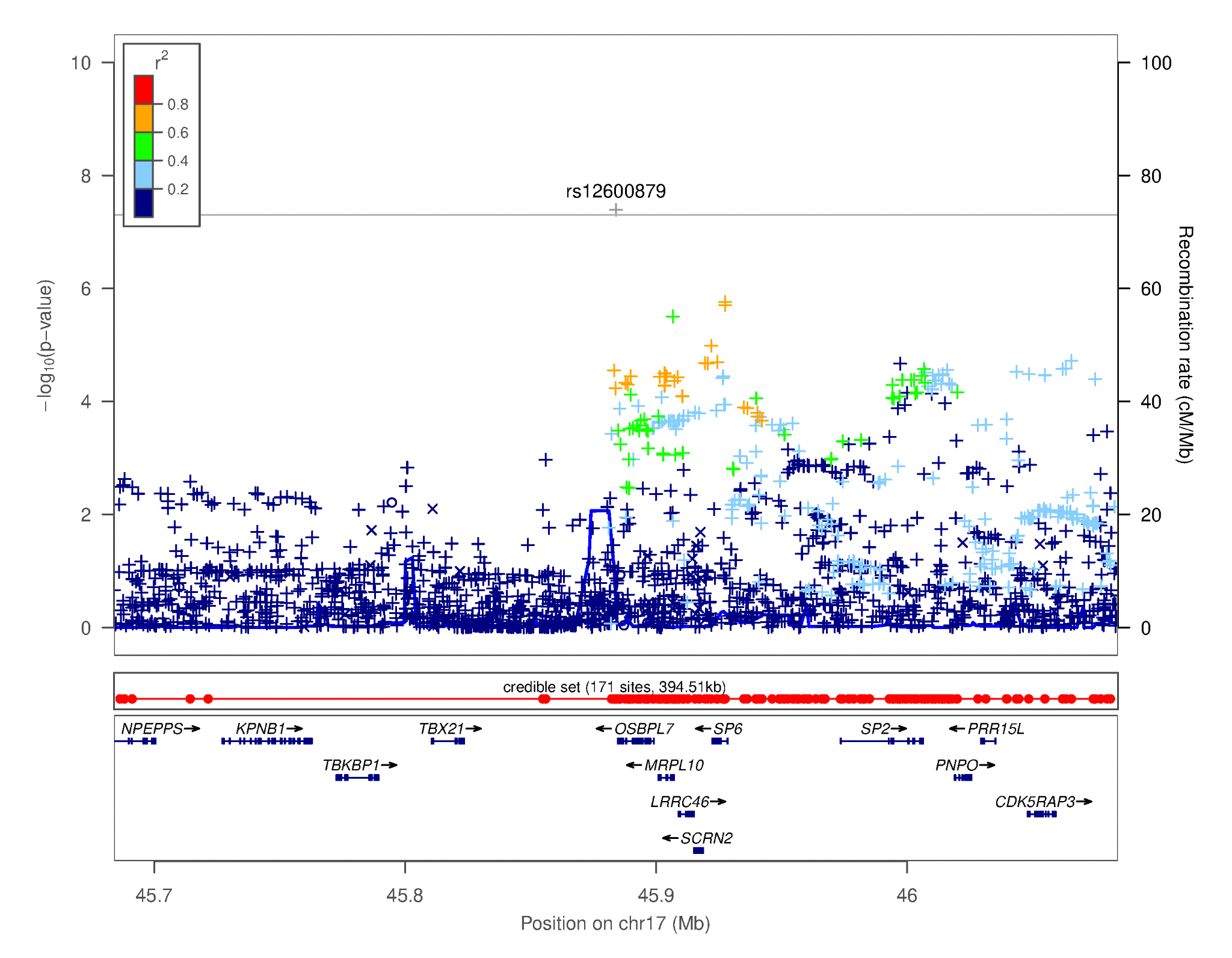

**Figure S13.** Regional association plots focusing on genetic variants associated with **UPPS-P Sensation Seeking**. This plot was generated using LocusZoom [16]. The -log_10_(*p-*value) is shown on the left *y****-***axis; position in Mb is on the *x***-**axis. Recombination rates (expressed in centiMorgans cM per Mb; NCBI Build GRCh37; highlighted in blue) are shown on the right *y****-***axis. Pairwise linkage disequilibrium (r^2^) of each SNP with the top SNP in the region is indicated by its color. Crossed points represent imputed SNPs, circles represent directly genotyped SNPs.

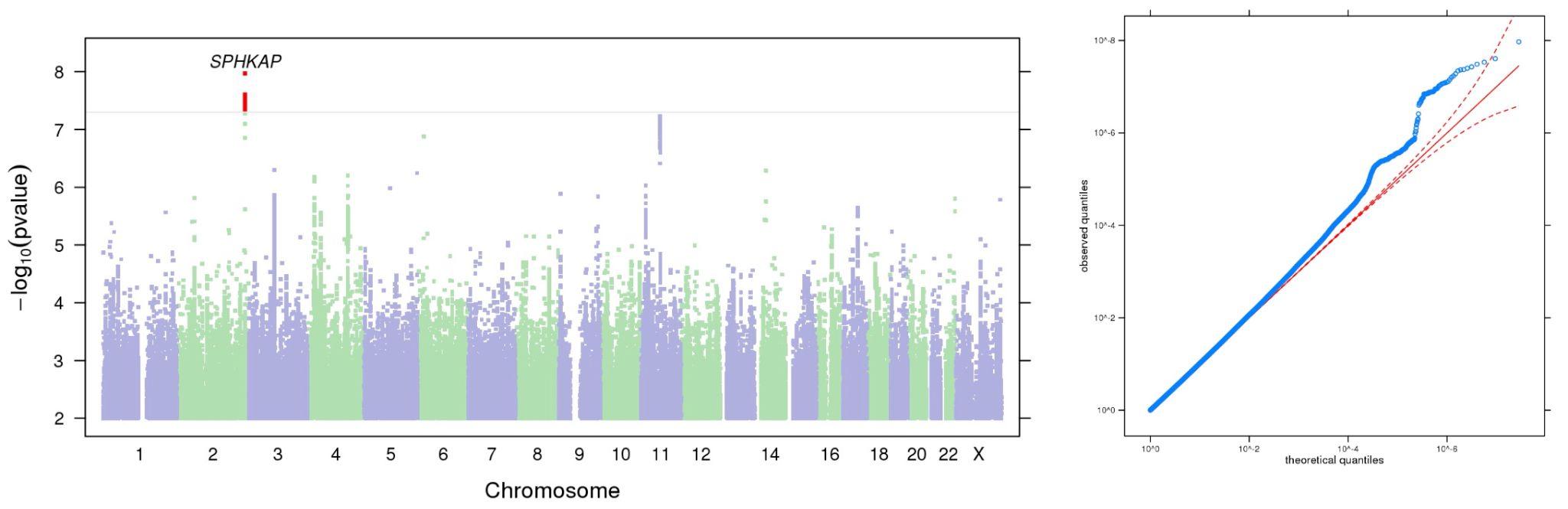
**Figure S14.** Manhattan and QQ plots of GWAS results indicating the strongest associations between the 22 autosomes, X chromosome, and **BIS Attentional**. The results have been adjusted for a genomic control inflation factor λ= 1.097 (sample size = 124,739).

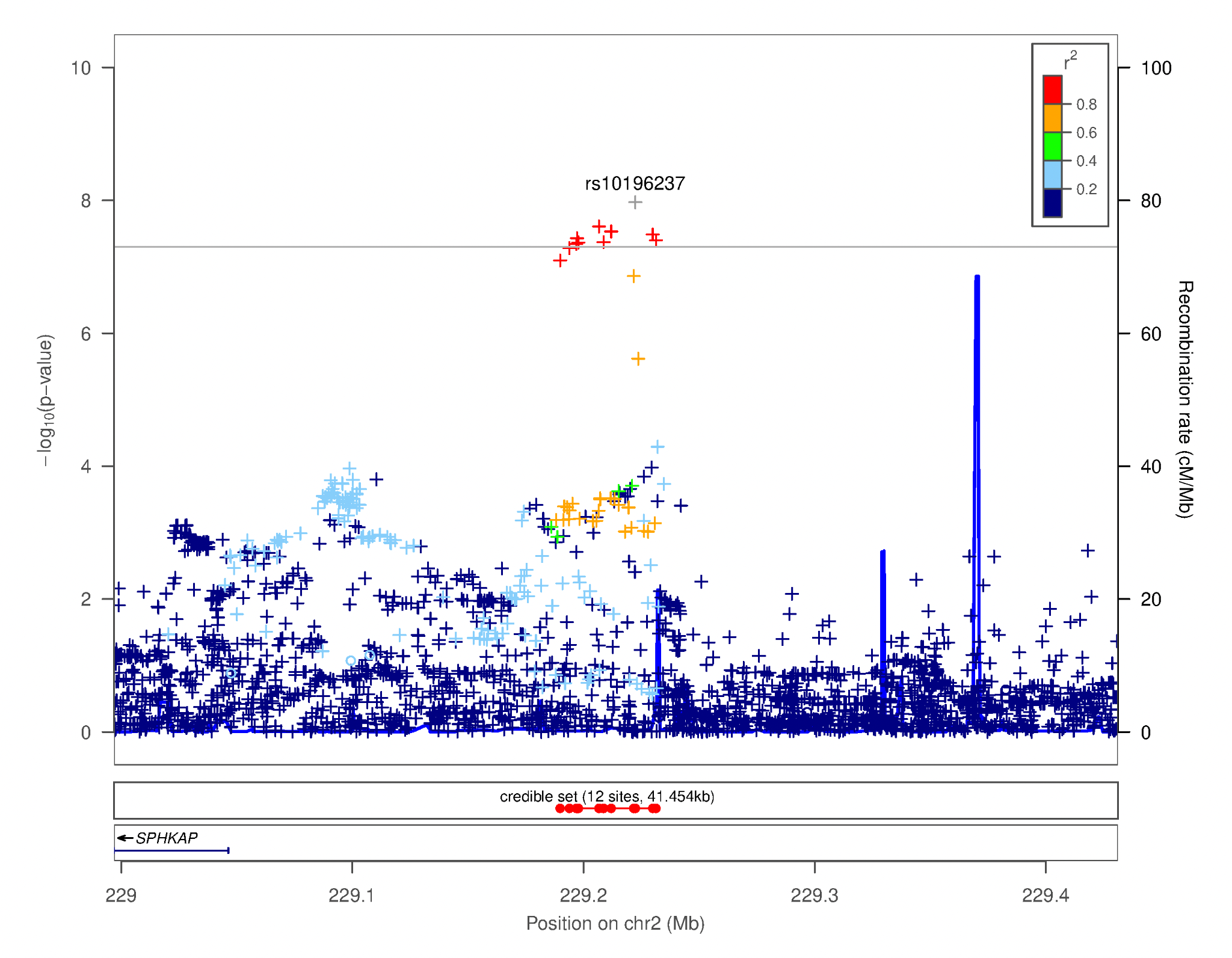

**Figure S15.** Regional association plot focusing on genetic variants associated with **BIS Attentional**. This plot was generated using LocusZoom [16]. The -log_10_(*p-*value) is shown on the left *y****-***axis; position in Mb is on the *x***-**axis. Recombination rates (expressed in centiMorgans cM per Mb; NCBI Build GRCh37; highlighted in blue) are shown on the right *y****-***axis. Pairwise linkage disequilibrium (r^2^) of each SNP with the top SNP in the region is indicated by its color. Crossed points represent imputed SNPs, circles represent directly genotyped SNPs.

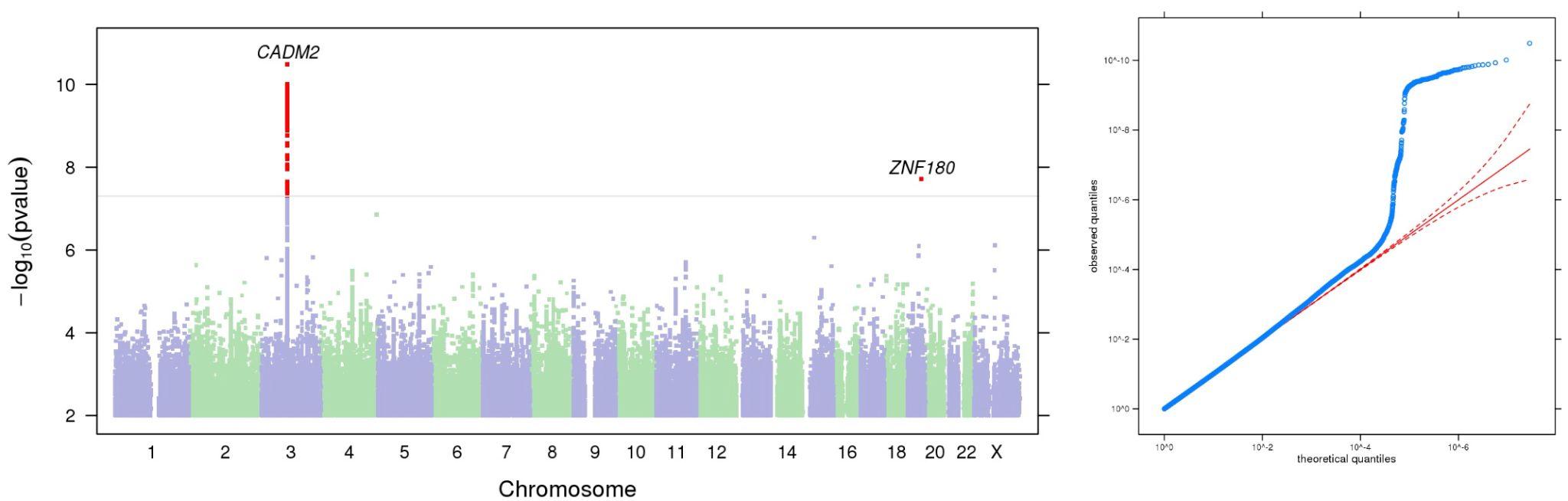

**Figure S16.** Manhattan and QQ plots of GWAS results indicating the strongest associations between the 22 autosomes, X chromosome, and **BIS Motor**. The results have been adjusted for a genomic control inflation factor λ=1.092 (sample size = 124,104).

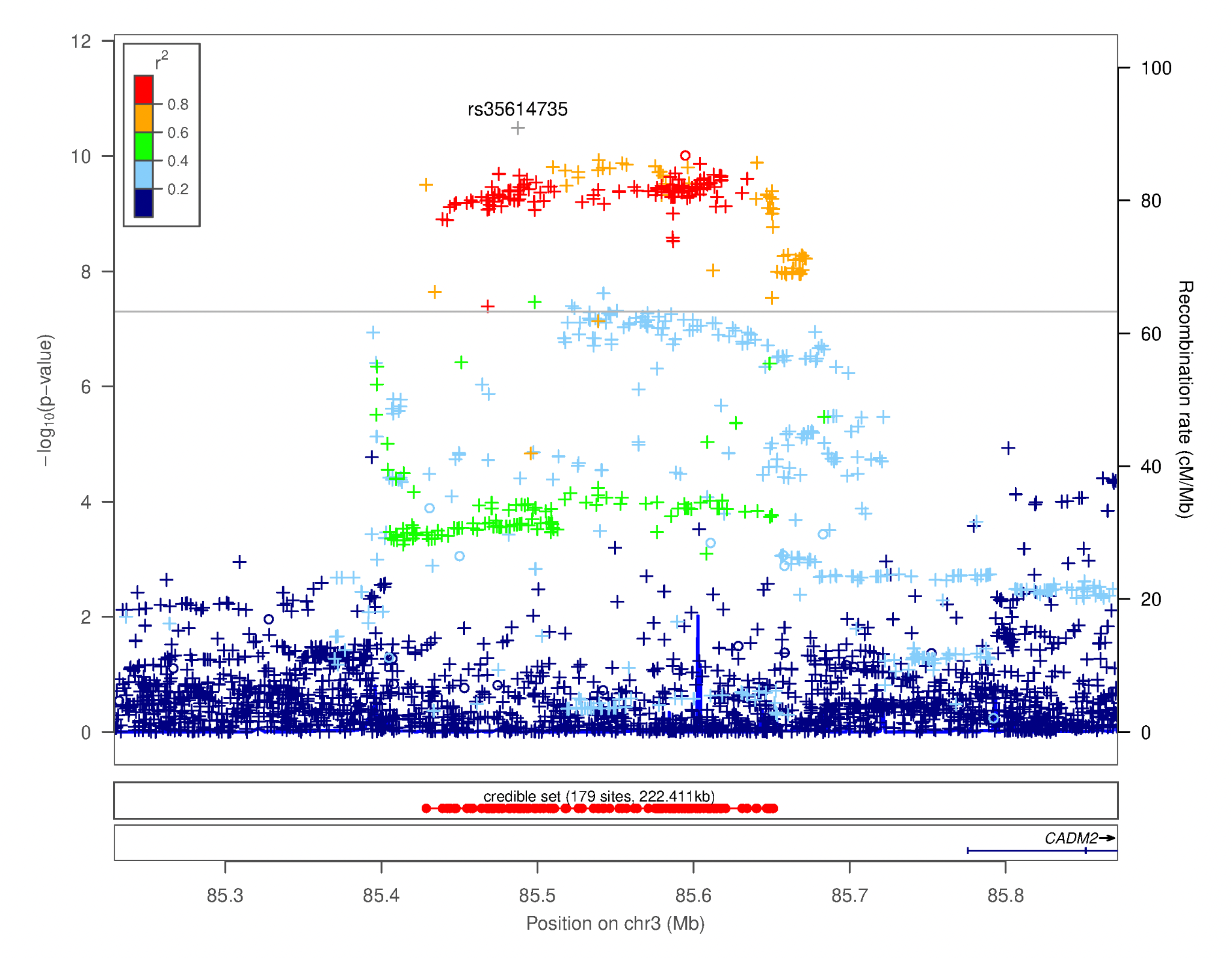

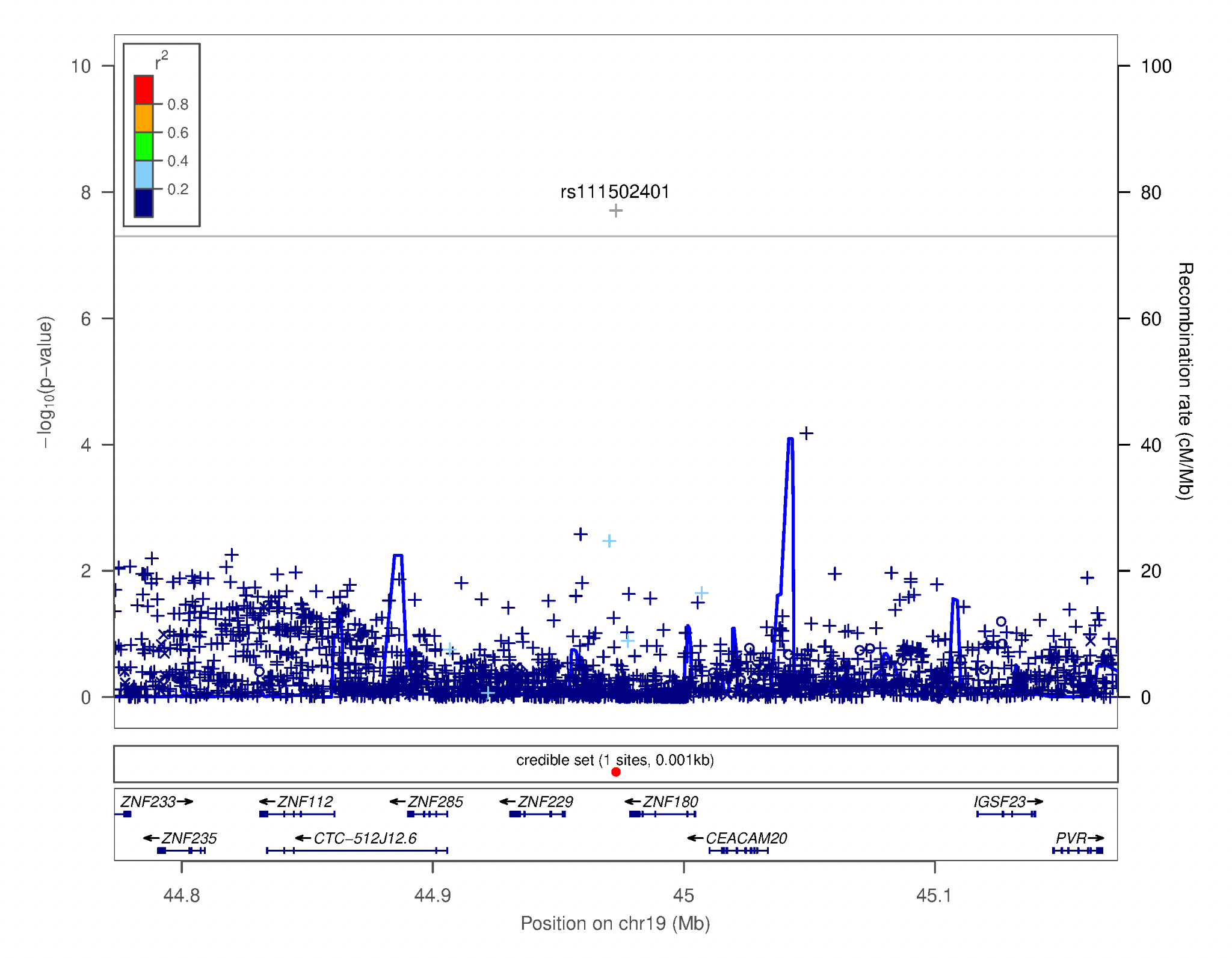

**Figure S17.** Regional association plots focusing on genetic variants associated with **BIS Motor**. This plot was generated using LocusZoom [16]. The -log_10_(*p-*value) is shown on the left *y****-***axis; position in Mb is on the *x***-**axis. Recombination rates (expressed in centiMorgans cM per Mb; NCBI Build GRCh37; highlighted in blue) are shown on the right *y****-***axis. Pairwise linkage disequilibrium (r^2^) of each SNP with the top SNP in the region is indicated by its color. Crossed points represent imputed SNPs, circles represent directly genotyped SNPs.

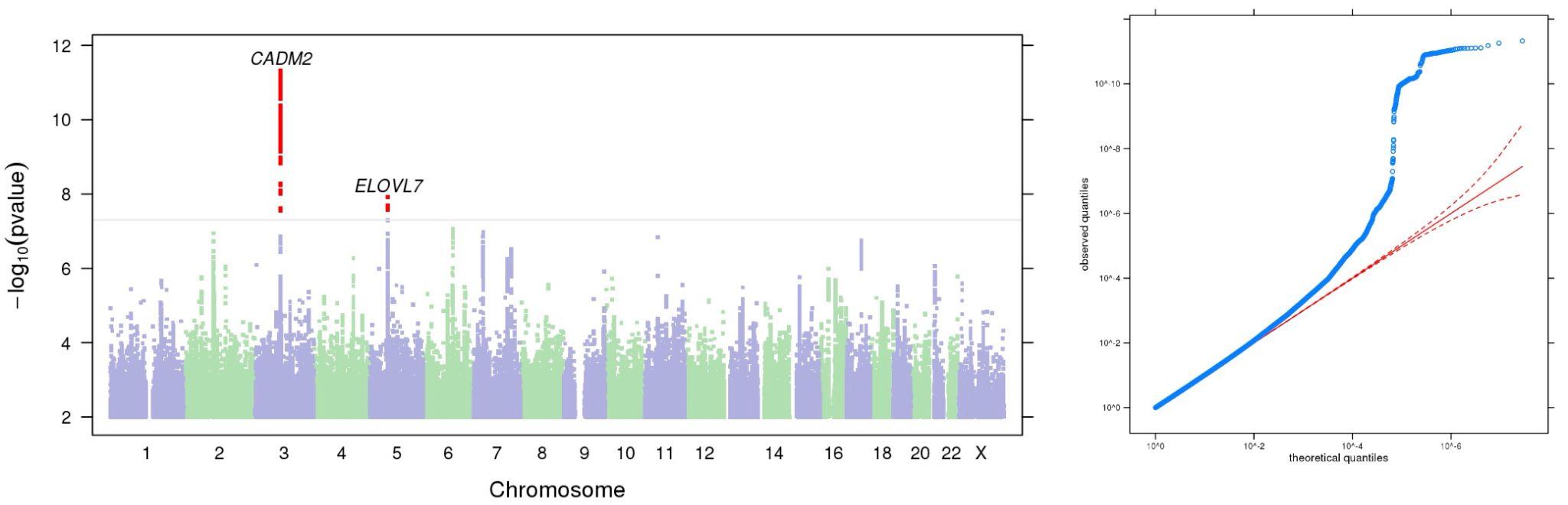

**Figure S18.** Manhattan and QQ plots of GWAS results indicating the strongest associations between the 22 autosomes, X chromosome, and **BIS Nonplanning**. The results have been adjusted for a genomic control inflation factor λ=1.123 (sample size = 123,509).

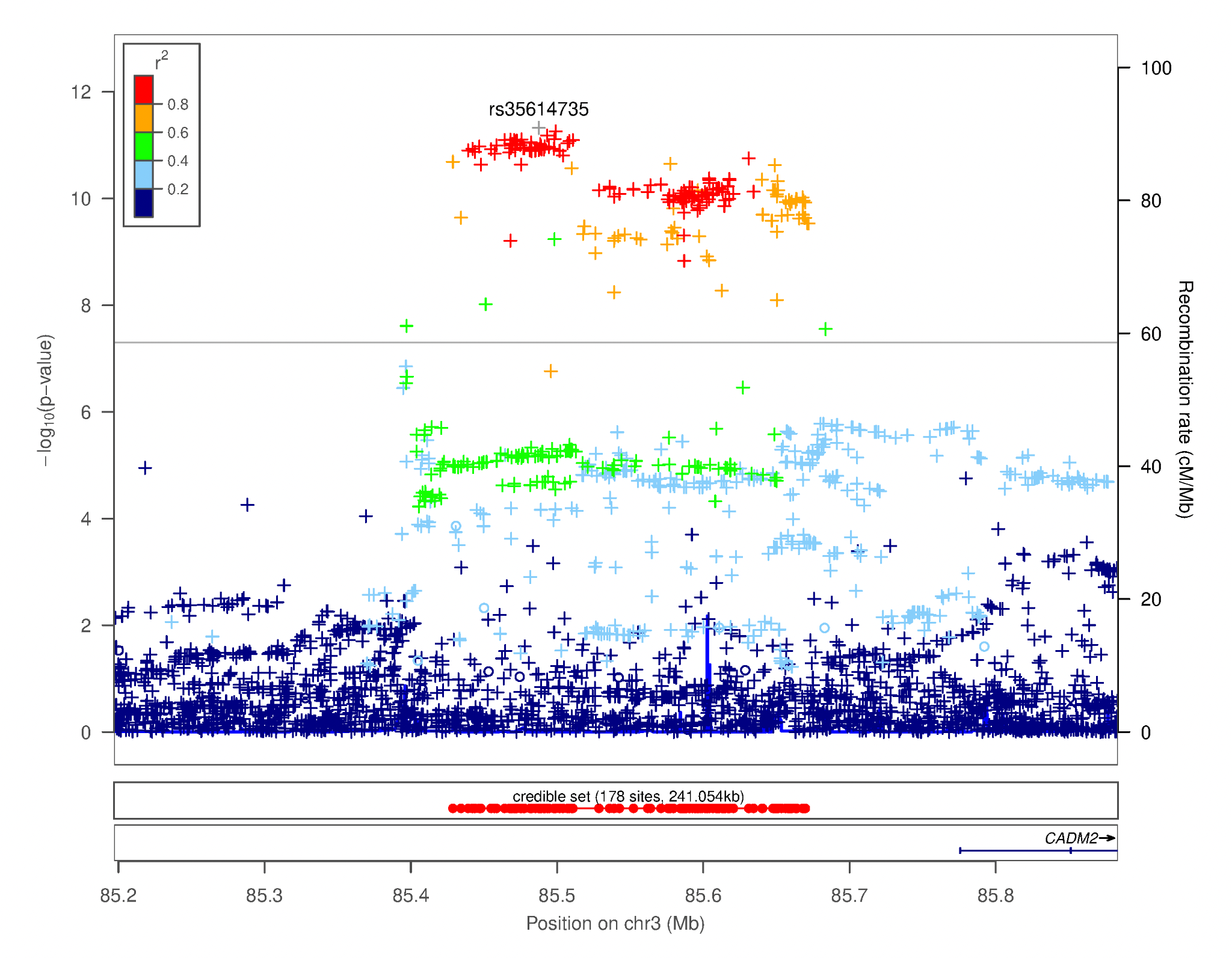

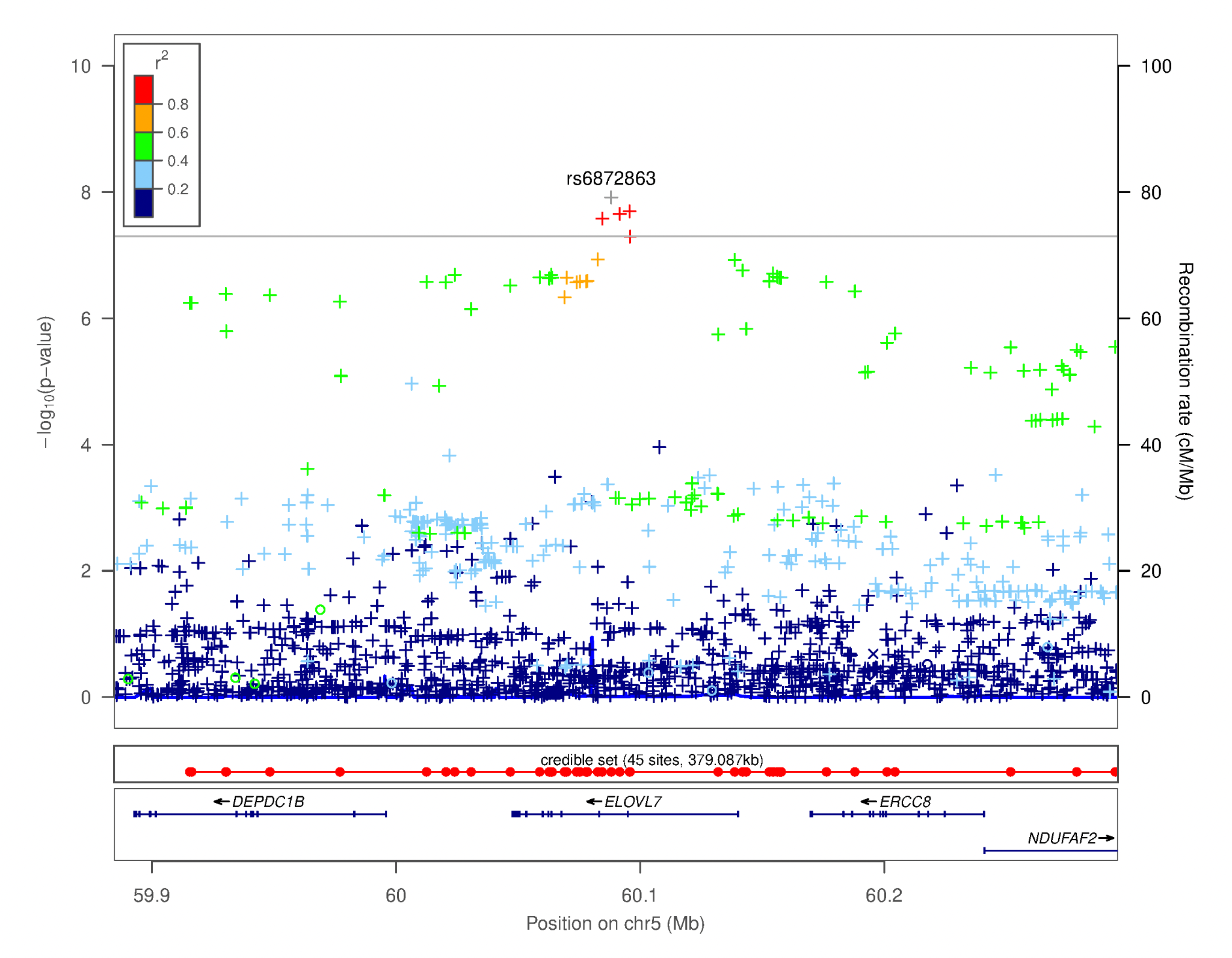

**Figure S19.** Regional association plots focusing on genetic variants associated with **BIS Nonplanning**. These plot were generated using LocusZoom [16]. The -log_10_(*p-*value) is shown on the left *y****-***axis; position in Mb is on the *x***-**axis. Recombination rates (expressed in centiMorgans cM per Mb; NCBI Build GRCh37; highlighted in blue) are shown on the right *y****-***axis. Pairwise linkage disequilibrium (r^2^) of each SNP with the top SNP in the region is indicated by its color. Crossed points represent imputed SNPs, circles represent directly genotyped SNPs.

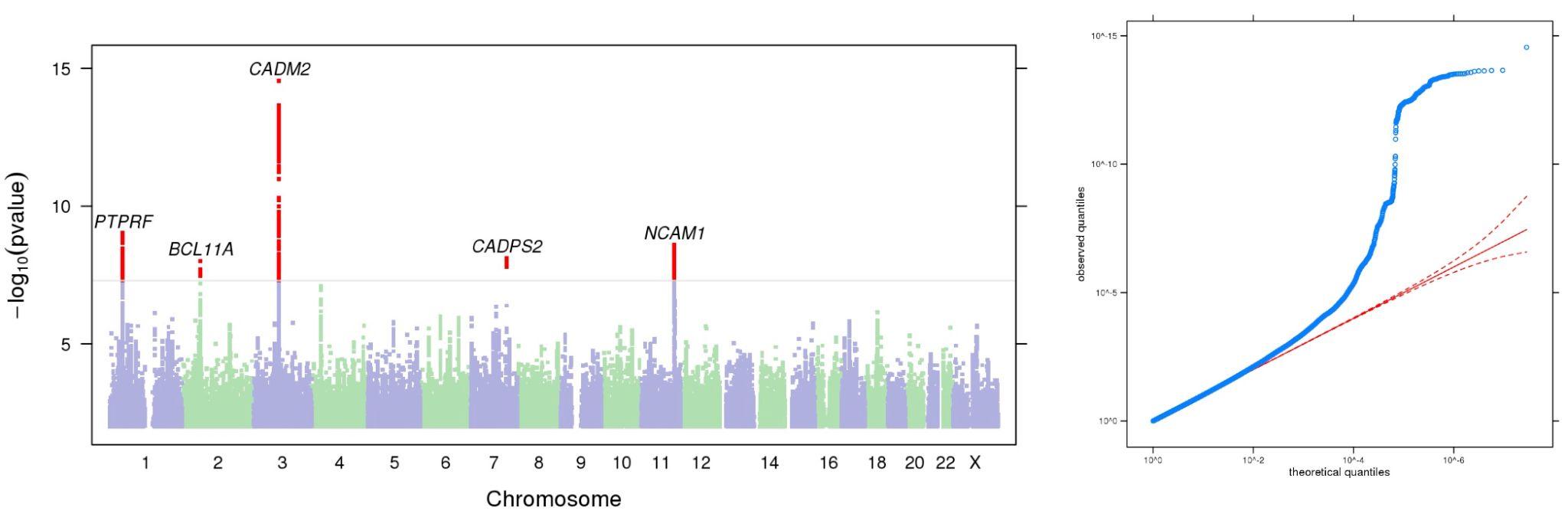

**Figure S20.** Manhattan and QQ plots of GWAS results indicating the strongest associations between the 22 autosomes, X chromosome, and **Drug Experimentation**. The results have been adjusted for a genomic control inflation factor λ=1.135 (sample size = 130,684).

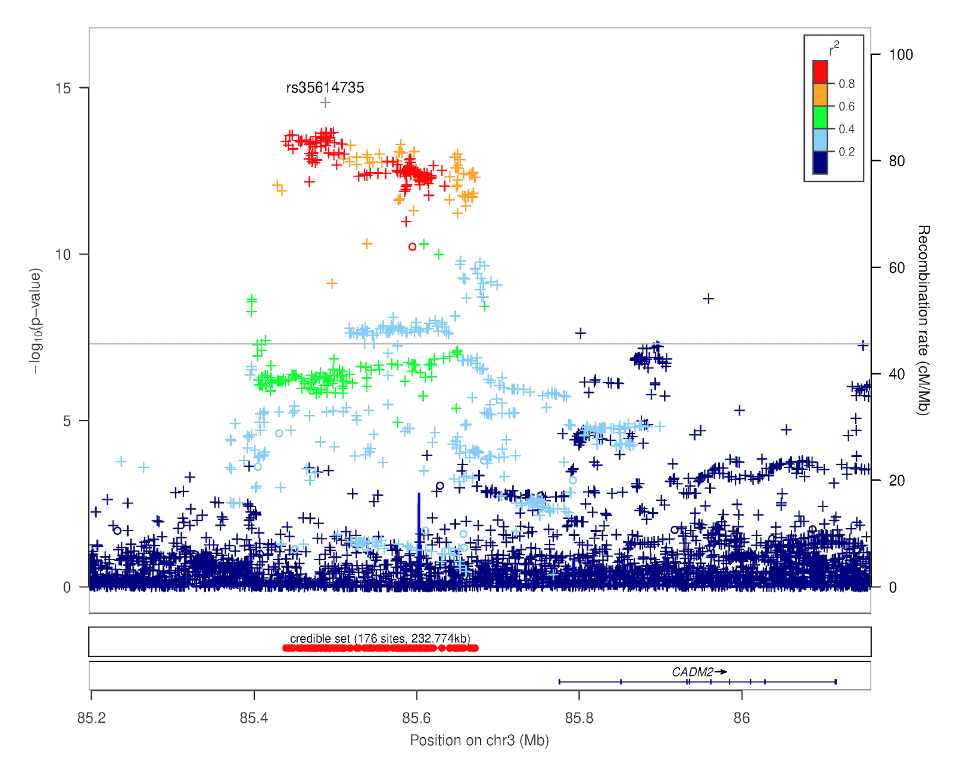

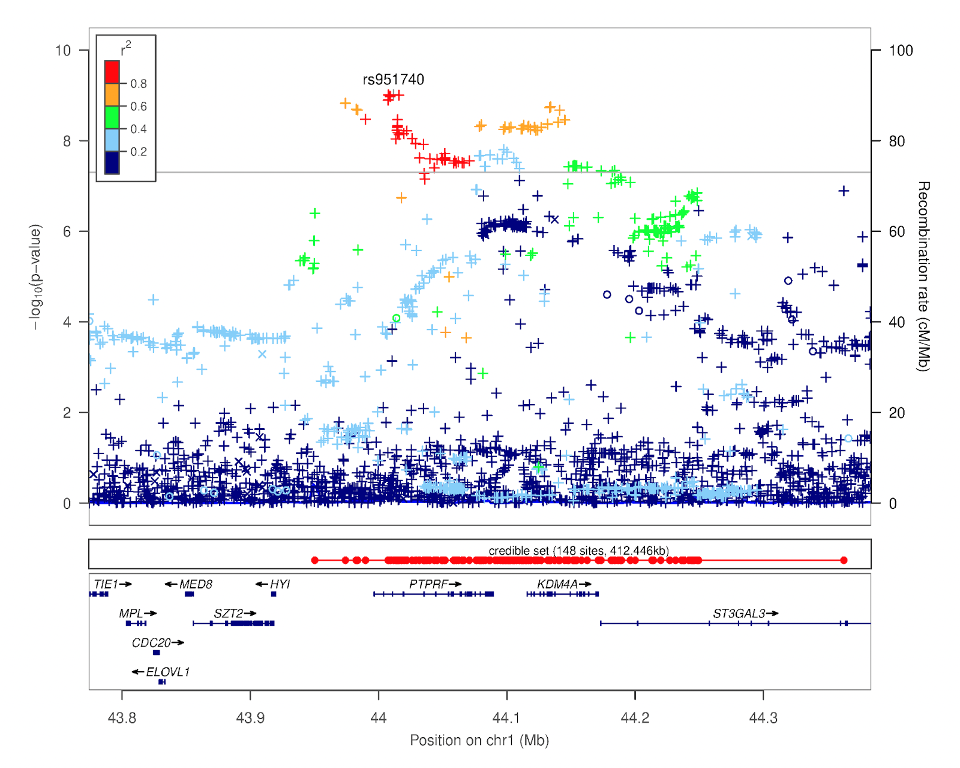

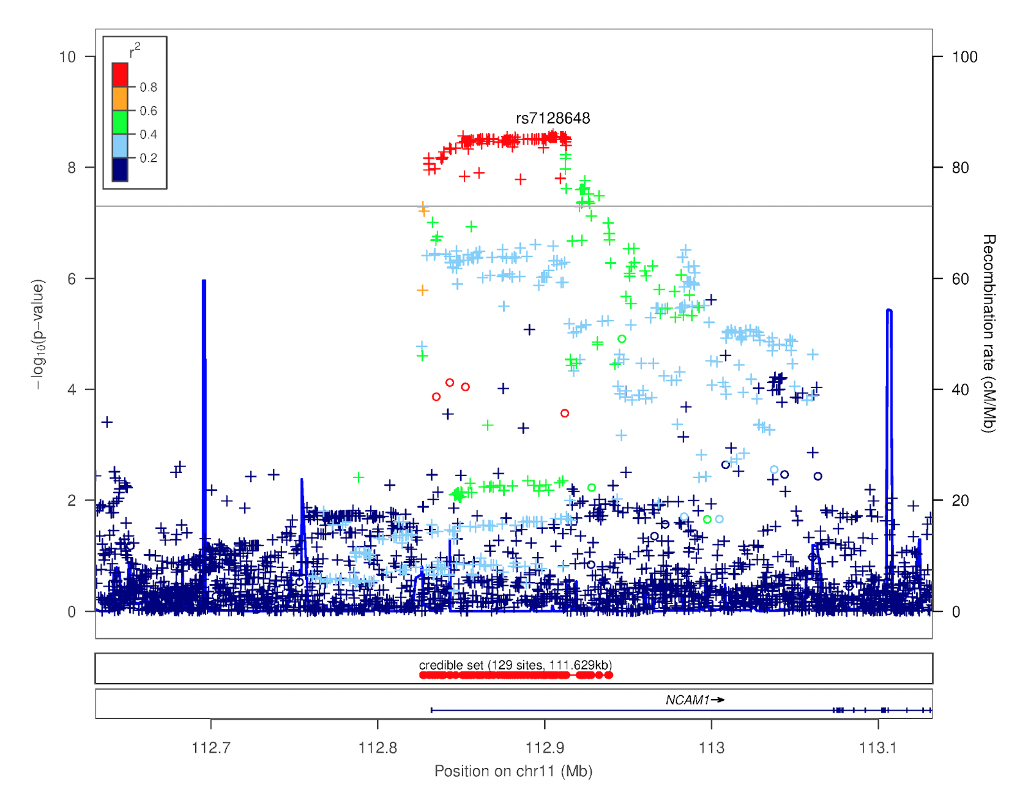

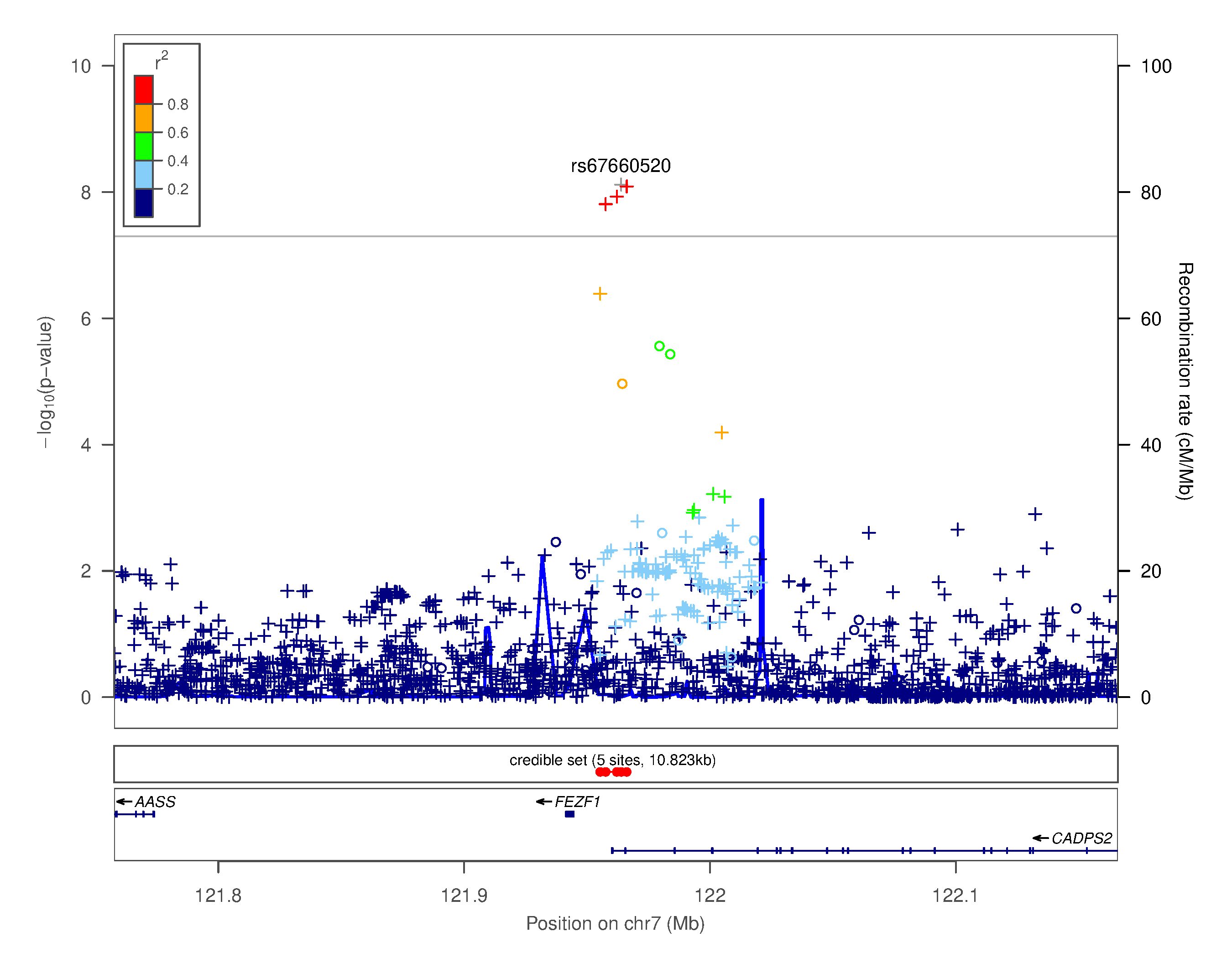

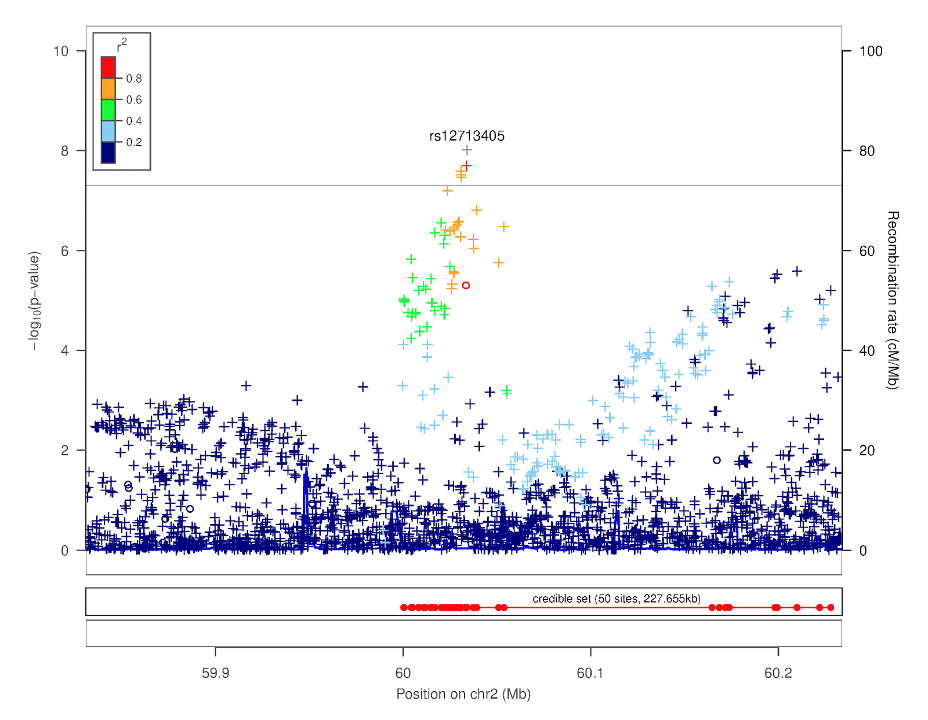

**Figure S21.** Regional association plots focusing on genetic variants associated with **Drug Experimentation**. These plots were generated using LocusZoom [16]. The -log_10_(*p-*value) is shown on the left *y****-***axis; position in Mb is on the *x***-**axis. Recombination rates (expressed in centiMorgans cM per Mb; NCBI Build GRCh37; highlighted in blue) are shown on the right *y****-***axis. Pairwise linkage disequilibrium (r^2^) of each SNP with the top SNP in the region is indicated by its color. Crossed points represent imputed SNPs, circles represent directly genotyped SNPs.

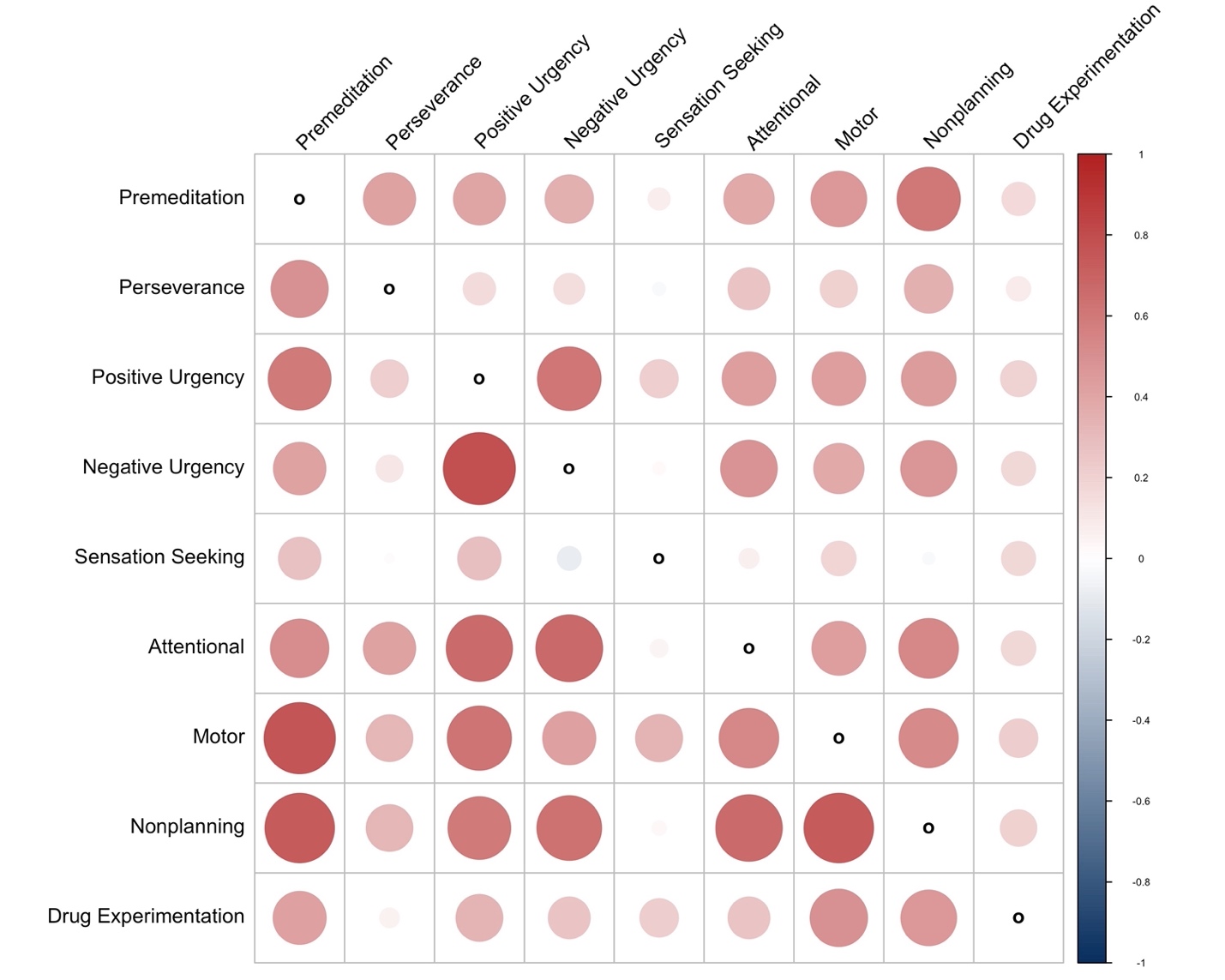

**Figure S22.** Genetic inter-correlations (lower diagonal) and phenotypic inter-correlations (upper-diagonal) between the UPPS-P, BIS-11 and Drug Experimentation traits. All inter-correlations values are shown in **Supplementary Tables 11-12**.

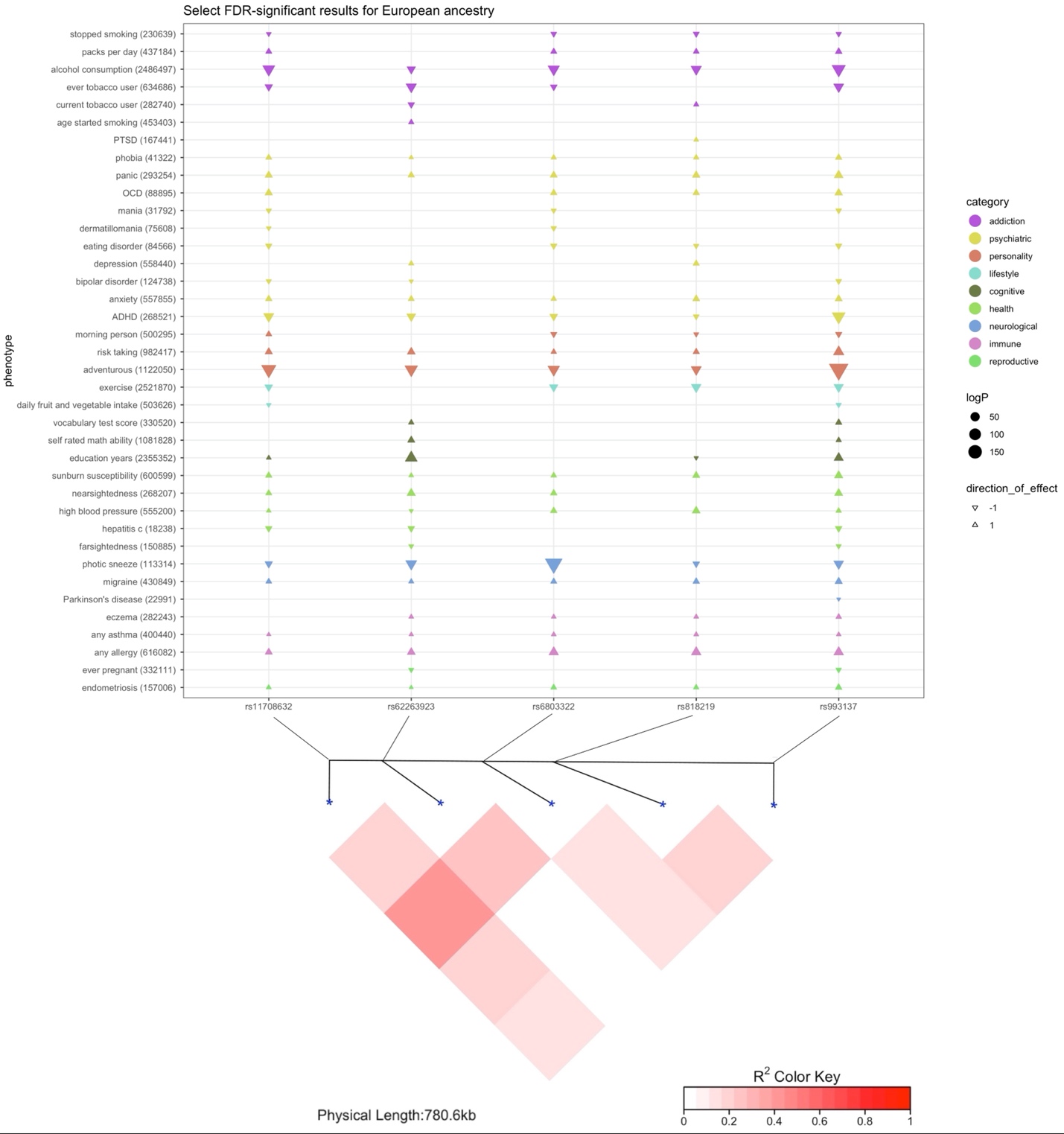

**Figure S23.** Results from PheWAS for select FDR-significant traits for each *CADM2* variant in European ancestry (top) and pairwise LD plot for all variants (bottom). Categories are identified via different colors, size of symbols represents the strength of the association (*p-*value) and the direction of the triangle indicates positive or negative effect sizes (β).

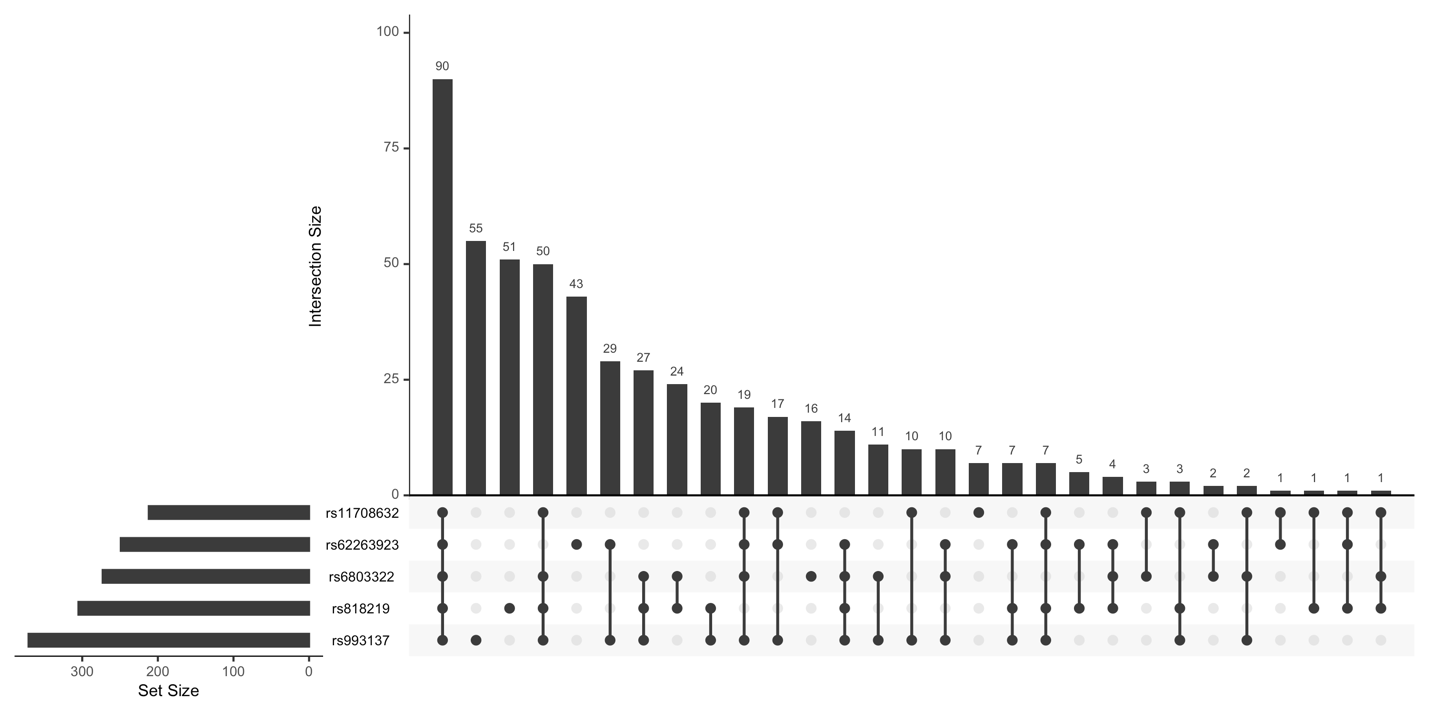

**Figure S24.** Overlap of FDR-significant traits for each of the tested variants in European ancestry. Set Size denotes the total number of FDR-significant traits for each SNP while Intersection Size shows the number of traits that overlap between the SNPs.

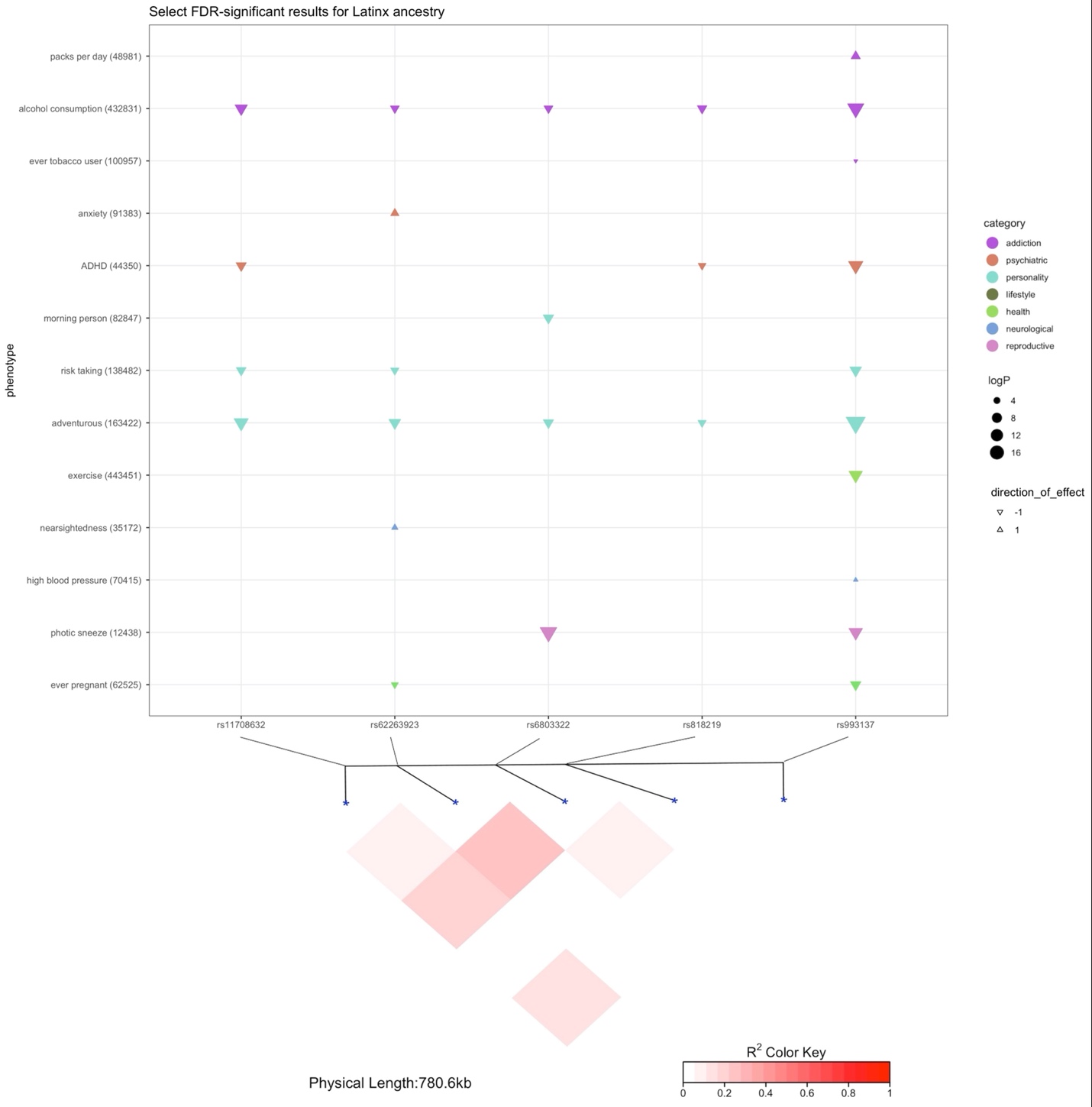

**Figure S25.** Results from PheWAS for selected FDR-significant traits for each *CADM2* variant in Latin American ancestry (top) and pairwise LD plot for all variants (bottom). Categories are identified via different colors, size of symbols represents the strength of the association (*p-*value) and the direction of the triangle indicates positive or negative effect sizes (β).

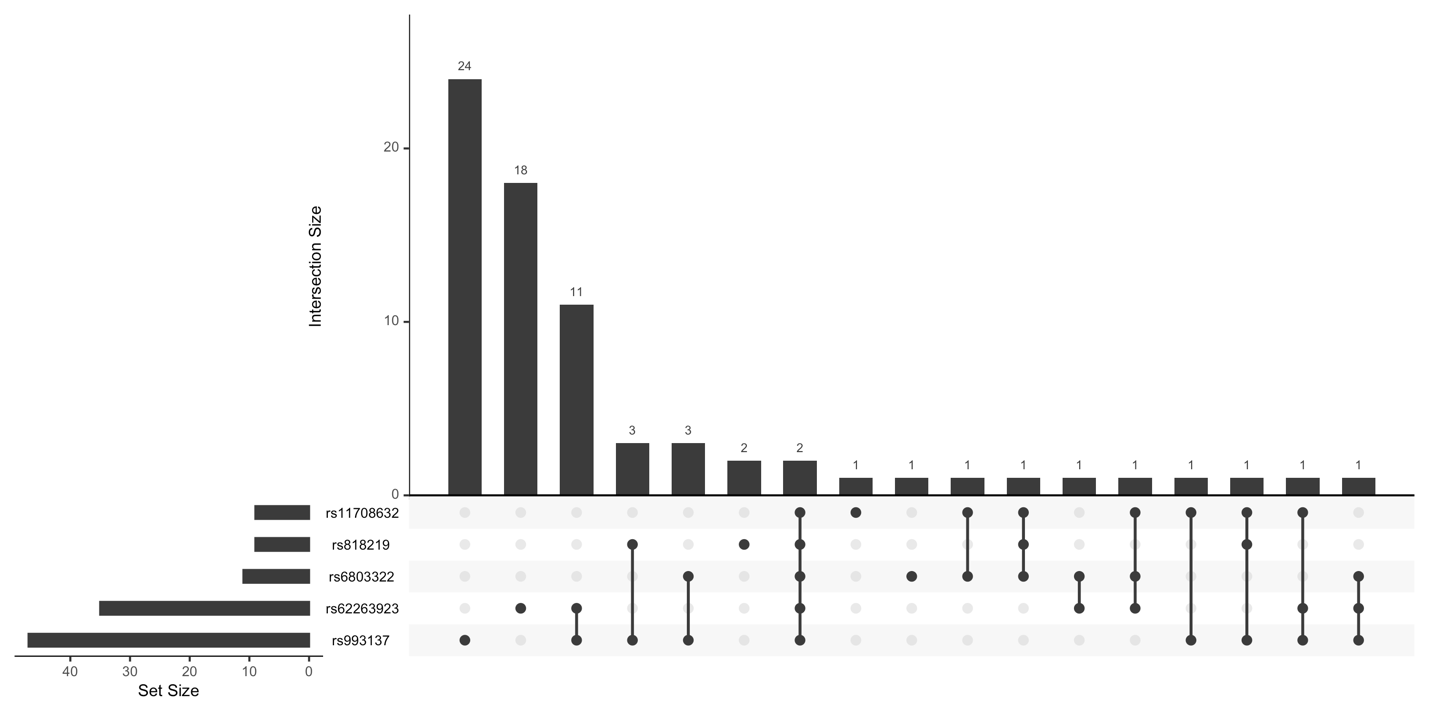

**Figure S26.** Overlap of FDR-significant traits for each of the tested variants in Latin American ancestry. Set Size denotes the total number of FDR-significant traits for each SNP while Intersection Size shows the number of traits that overlap between the SNPs.

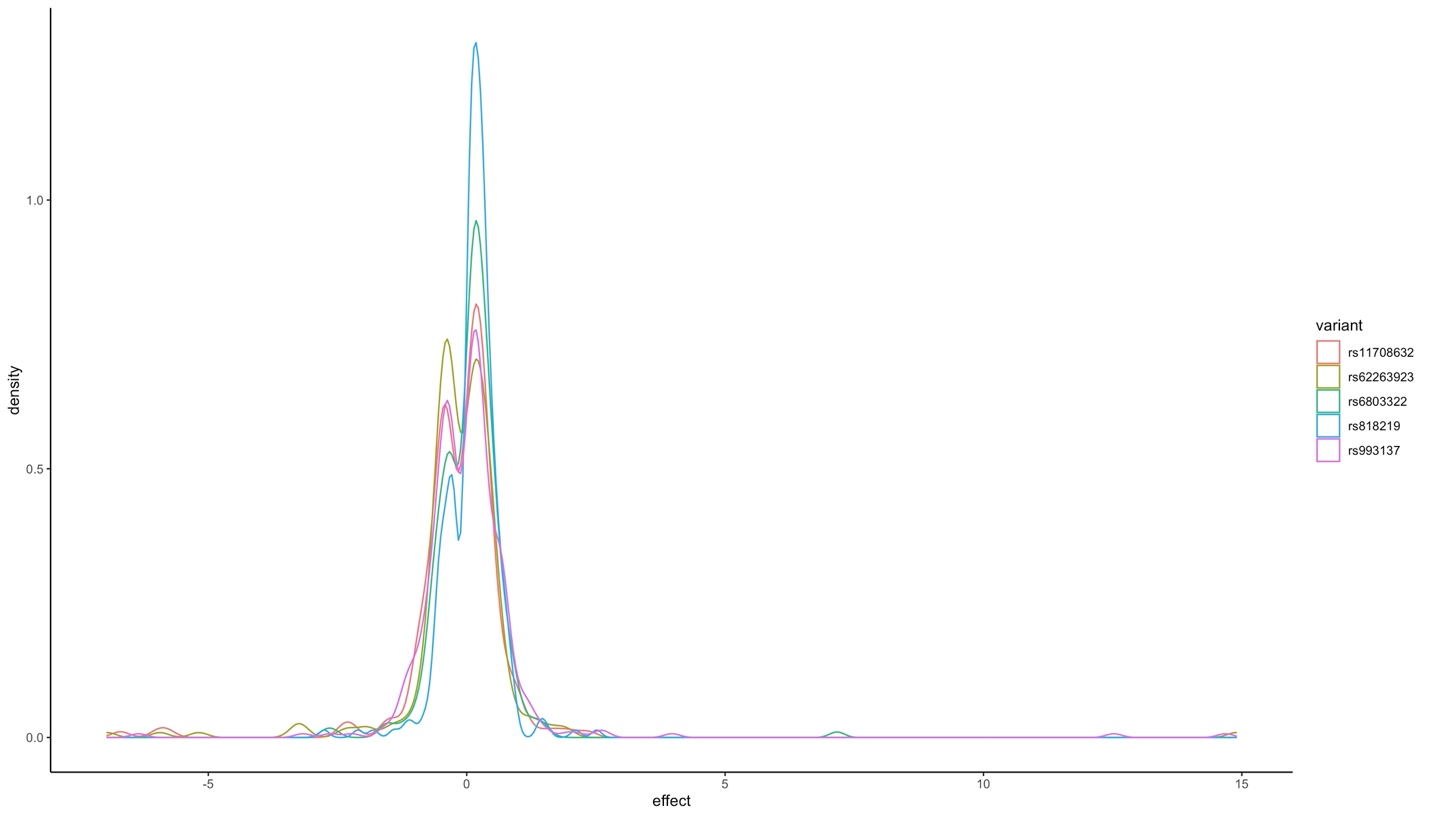

**Figure S27.** Range of standardized effect sizes (non-standardized β= -0.32 to 0.70) for FDR-significant traits in European ancestry.

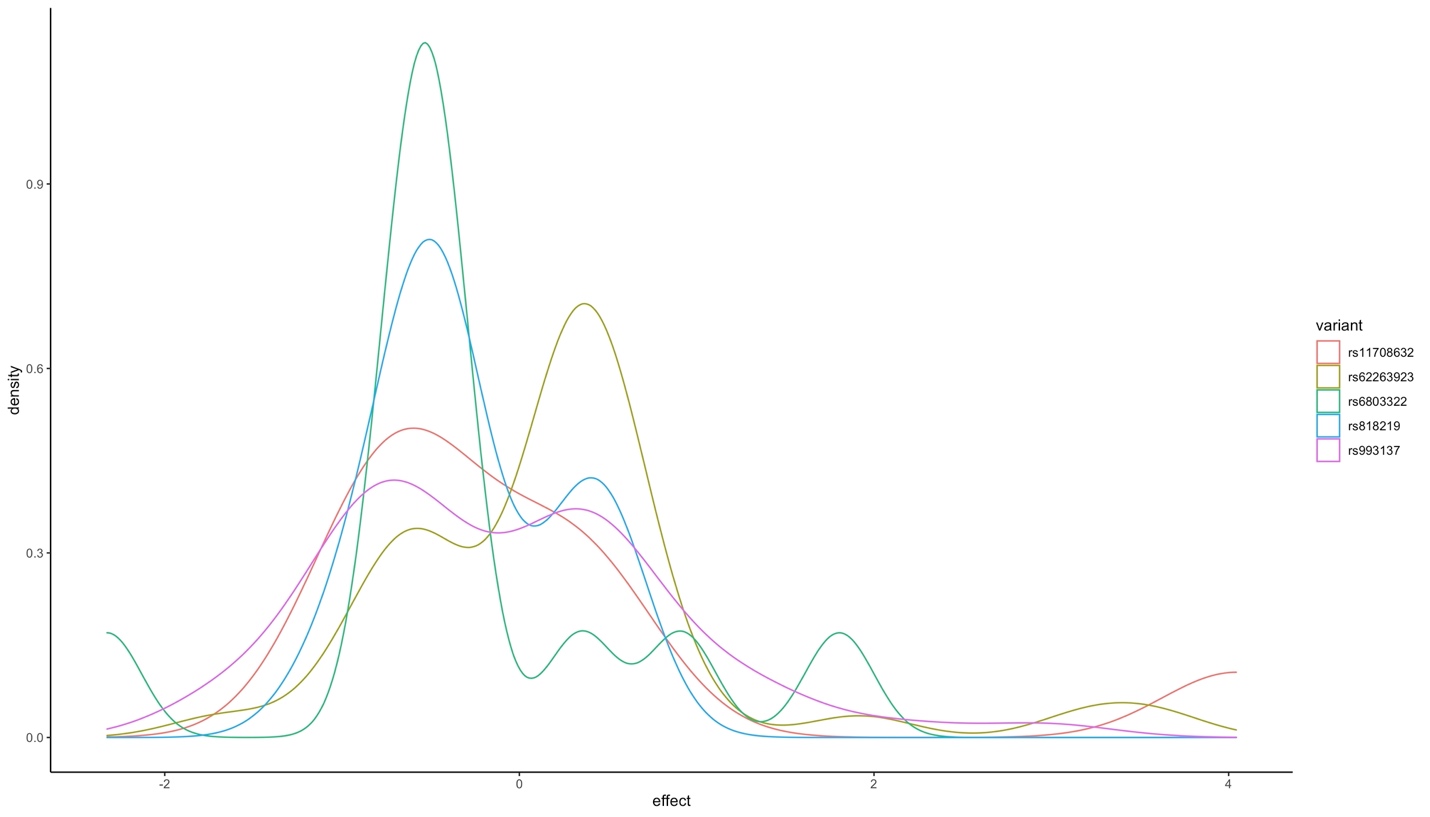

**Figure S28.** Range of standardized effect sizes (non-standardized β=-0.11 to 0.22) for FDR-significant traits in Latin American ancestry.

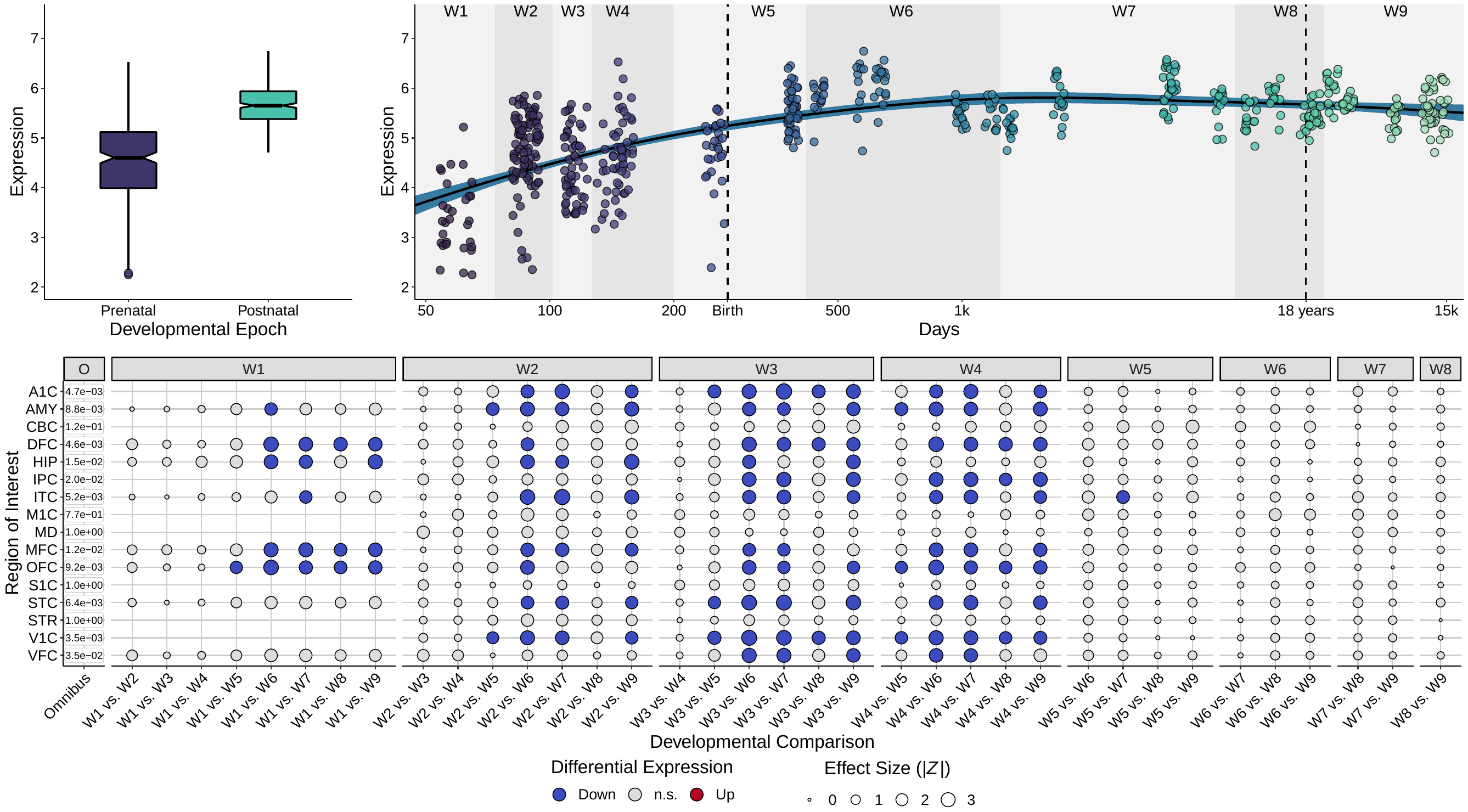

**Figure S29.** Differential expression of *CADM2* across development in human brain tissue, as measured in the PsychENCODE (PEC) dataset [17]. Normalized gene expression values are plotted as log_2_ RPKM + 1. In the top left panel, *CADM2* expression in human brain tissue is plotted as box plots stratified by prenatal and postnatal epoch. In the top right panel, the trajectory of *CADM2* expression in human brain tissue is plotted across development, as estimated with LOESS. Postconceptional days are plotted in log_2_ scale on the x-axis. In the bottom panel, Kruskal-Wallis and Dunn’s tests of differential expression for developmental windows (W1-W9) in specific regions of interest (ROIs) are plotted as a bubble plot. The *P* values for Kruskal-Wallis tests in the leftmost column are corrected for false discovery rate (FDR) across ROIs (16 tests), whereas the *P* values for Dunn’s tests are corrected within ROIs (up to 36 tests). The ROIs included were primary auditory cortex (A1C), amygdala (AMY), cerebellar cortex (CBC), dorsolateral prefrontal cortex (DFC), hippocampus (HIP), inferior parietal cortex (IPC), inferior temporal cortex (ITC), primary motor cortex (M1C), thalamus (MD) medial prefrontal cortex (MFC), orbital prefrontal cortex (OFC), primary somatosensory cortex (S1C), superior temporal cortex (STC), striatum (STR), primary visual cortex (V1C), and the ventrolateral prefrontal cortex (VFC). The methods used for this analysis have been previously extensively described [18,19].

*Mouse studies*

**Establishment of the *Cadm2* mouse line**

**KOMP mouse establishment**

We created the *Cadm2* mutant mice on a C57BL/6N background, by crossing 1) *Cadm2* Komp mouse line (retrieved from KOMP sperm; order number #KO-6555) and 2) strain B6N.Cg-Tg(Sox2-cre)1Amc/J (Stock# 014094) acquired from The Jackson Laboratory. This cross created a *Cadm2* null allele. Mice that were heterozygous (**HET**) for the null allele were mated with subsequent HET mice to yield homozygous (**HOM**), HET, and wildtype (**WT**) *Cadm2* mutants. Genotyping was performed using the following primers ATGCTACCTGAGCCTGTTTCCAAGG and GCTACCATTACCAGTTGGTCTGGTGTC (forward: WT and HOM, respectively), and AACACTTTCATTGTCACAGGACTGC and CTGTCATTCATCAGCATCCTCTGGG (reverse: WT and HOM, respectively; all sequences are shown in the 5' to 3' orientation).

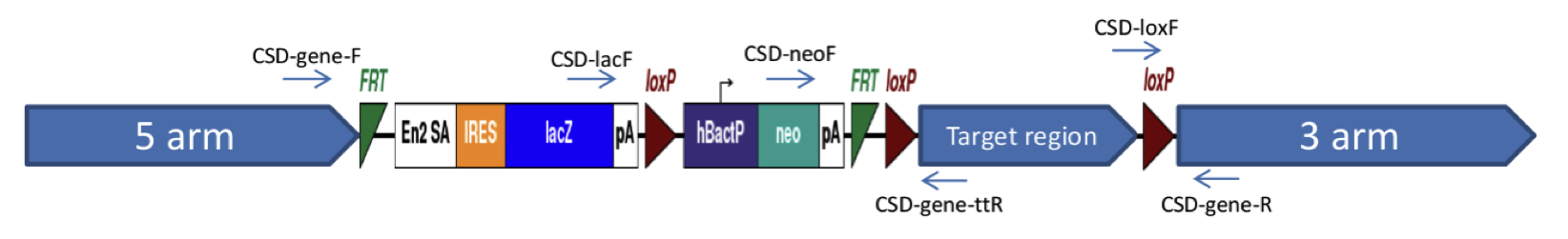

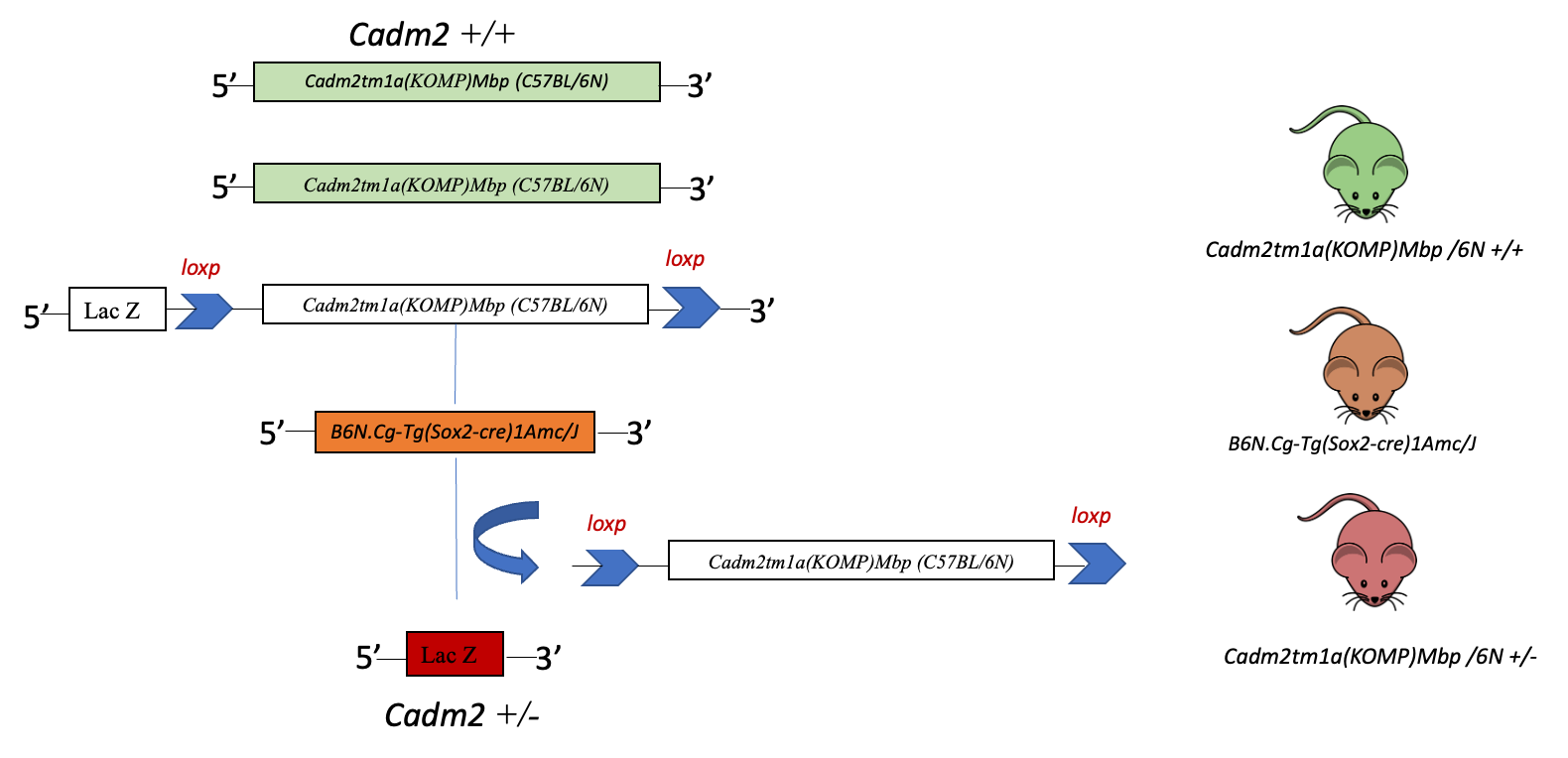

**Figure S30.** Targeted exon and mouse breeding figure.

**Western Blotting**

Our western blotting protocol was modified from Mendell et al. 2020 [20]. Animals were sacrificed at PND79 using CO_2_ and cervical dislocation. Whole brain tissues were harvested and homogenized with a Dounce homogenizer in lysis buffer (50 mM Tris-HCL (pH 7.5), 150 mM NaCl, 1% Triton-X 100) mixed with 5 μM sodium orthovanadate and protease inhibitor tablets (cOmplete™ Roche, 11836153001) according to manufacturer instructions. Lysates were briefly sonicated, incubated on ice for 20 minutes, and centrifuged at 17,530×g for 15 minutes at 4°C. Protein was quantified by Bradford assay (Alfa Aesar #J61522).

Protein samples were incubated with 4X Laemmli sample buffer (Bio-Rad #1610747) and 2-mercaptoethanol according to manufacturer instructions. Using the Mini-PROTEAN system (Bio-Rad, Mississauga, ON, Canada), 10 μg of protein was loaded into a 4% stacking, 10% resolving SDS-PAGE gel and electrophoresed at 110 V until sample buffer run-off was observed. Protein was transferred to a PVDF membrane using the Trans-Blot Turbo Transfer System (Bio-Rad #1704150) and Trans-Blot Turbo RTA kits (Bio-Rad #1704274) as directed for 7 minutes at 25 V. Membranes were blocked in 5% bovine serum albumin (BSA) in tris-buffered saline (TBS) containing 0.1% Tween-20 (TBST) for 90 minutes. Blots were cut and sections were incubated on a rocker overnight at 4°C in 1% BSA blocking solution containing primary antibody (1:1,000 CADM2, Synaptic Systems 243203; 1:10,000 GAPDH, Abcam ab181602). Blots were rinsed three times for 8 minutes each in TBST, then incubated on a rocker at room temperature in secondary antibody (1:50,000 goat anti-rabbit IgG-HRP, Bio-Rad 1706515) for 60 minutes. Blots were rinsed three times for 8 minutes each in TBST. Enhanced chemiluminescent solution was applied to blots for two minutes, briefly rinsed with TBST, and blots were imaged and quantified by densitometry using the Bio-Rad ChemiDoc MP imaging system with ImageLab software (Bio-Rad Ver 6.1.0).

Differences in whole brain CADM2 protein expression between WT and HOM (n=3/group, all males) were assessed in three pairs of littermate-matched mice, and frontal cortex and striatal expression of CADM2 protein were assessed in WT (n=5; 2 male, 3 female), HET (n=5; 3 male, 2 female) and HOM (n=3; 2 male, 1 female) littermates. CADM2 protein densitometry readings were normalized to GAPDH loading control band intensity. The difference in CADM2 protein expression was quantified as the percent difference from average WT littermate band intensity. Whole brain percent protein change was statistically assessed using a one-tailed t-test. Frontal cortex protein change was assessed by one-way ANOVA followed by Bonferroni post-hoc testing and striatal protein change was assessed by Kruskal-Wallis one-way ANOVA with Bonferroni post-hoc testing.

**CADM2 protein expression results**

Western blot analysis of protein lysate revealed an approximate 84% reduction (*t* = 4.41, *p*=6.00E-03) in CADM2 protein expression in the whole brains of WT relative to HOM brains (**Figure S31**). Expression in the frontal cortex was similarly altered by genotype [F_(2, 36)_=8.58, *p*=7.00E-03], with an approximate 95% reduction in HOM (*p*=1.30E-02) and 71% reduction in HET (*p*=2.40E-02) CADM2 expression compared to WT littermates. In the striatum, CADM2 expression was significantly different according to genotype [H_(2)_=7.48, *p*=2.40E-02], with a significant reduction in HOM mice (*p*=1.50E-02) of approximately 83% and a marginal reduction of 54% (*p*=7.90E-01) in HET mice relative to WT.

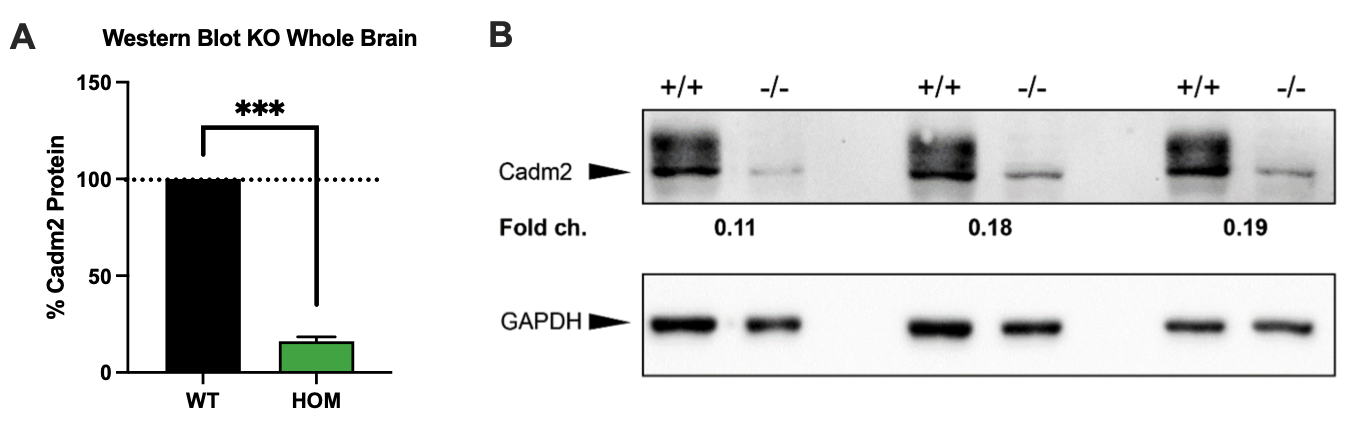

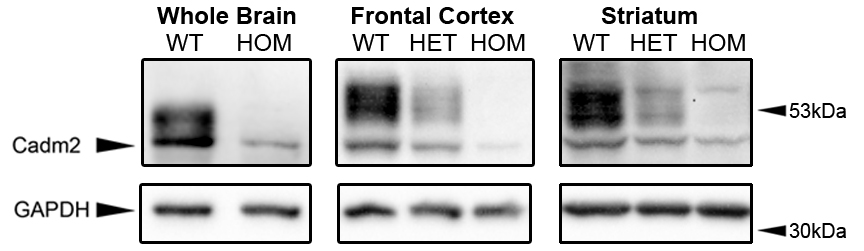

**Figure S31.** Cadm2 protein expression is significantly reduced in HOM mice relative to WT littermates. The left panel illustrates how knockout of the *Cadm2* gene depletes whole brain CADM2 protein expression by approximately 84% relative to WT littermates. The right panel shows representative Western blots of whole brain, frontal cortex, and striatal lysates probed for CADM2 protein.

**Mouse behavioral testing – General Overview**

HOM, HET, and WT littermates were tested in parallel. Mice were between 4 and 6 months of age at the time of testing, unless otherwise stated. Lighting, housing and feeding conditions for each cohort are shown in the next sections below.

*Cohort 1 - Motivation, inhibition, and risk-taking behavior.* The first cohort (WT=25, HET=30, HOM=3) was used to examine risky behavior or “choice” impulsivity, behavioral flexibility, prepulse inhibition (**PPI**) of the acoustic startle response, and general exploration. The primary measure of choice impulsivity included risky behavior via the mouse Iowa gambling task (**IGT** [21,22]). Because *Cadm2* has been previously associated with alcohol consumption, we used a within-subjects design to assess risky behavior under acute doses of ethanol (0, 0.5, 1g/kg), as previously described [23]. In addition, we evaluated motivation, as measured by a Progressive Ratio Breakpoint task (**PBRT** [24,25]), and behavioral flexibility, as measured by a Probabilistic Reversal Learning task (**PRL** [26]). PPI took place in eight startle chambers (SR-LAB; San Diego Instruments, San Diego, California, USA), using previously published protocols [27,28]. General exploration was measured via the Behavioral pattern monitor (**BPM** [29,30]). Data from PRBT, PRL, IGT, BPM and PPI were subjected to a univariate ANOVA with sex and genotype as between-subject factors. Data from the IGT ethanol challenge were analyzed using a repeated measure ANOVA with drug as a within-subject factor and genotype and sex as between-subject factors. The sample size of the HOM mice deviated from the expected Mendelian frequency (N=3) for no known reason; therefore, we excluded this group from the analyses of this cohort.

*Cohort 2 - Motoric impulsivity.* The second cohort (WT=13; HET=14, HOM=12) was used to examine “motoric” impulsivity via the 5-choice serial reaction time task [**5CSRTT** [23,31]]. Reinforcer preference, baseline and long ITI performance were analyzed using univariate ANOVA for parametric data or Kruskal-Wallis ANOVA for nonparametric data with genotype as the between-subject factor. Reduced stimulus duration (**RSD**) and variable ITI (**vITI**) performance were analyzed with genotype and stimulus duration (RSD) or stimulus delay (vITI) as the between- and within-subject factors, respectively, using two-way repeated-measures ANOVA for parametric data or two-way between-within subjects ANOVA on trimmed means for nonparametric data. Due to the low number of HOM females (N=4), sex was not included in the analyses of this cohort.

*Cohort 3 - General locomotion, anxiety-like behavior, and ethanol consumption.* The third cohort (WT=22; HET=44, HOM=12) was used to measure general locomotion, anxiety-like behavior, and ethanol consumption. Precisely, we assessed general locomotion via the Open Field [32–35], and anxiety-like behavior via the elevated plus maze [**EPM** [36]] and light-dark box [**LDB** [37]]. Lastly, we measured acute voluntary ethanol consumption using a drinking-in-the-dark paradigm [**DID**, [38]]. OFT, EPM and LDB data were assessed for normality and genotype effects were analyzed by ANOVA, with between-subject factors of genotype and sex. For the DID, ethanol drinking data were averaged and converted from milliliters to g ethanol/kg body weight for analysis. Drinking data were analyzed using ANOVA, with between-subject factors of genotype and sex.

*Cohort 4 - Body weight.* Considering the previous role of *Cadm2* on energy homeostasis [39], the fourth cohort (WT=29, HT=54, HOM=17) was used to assess body weight changes from adolescence (week 5) to late adulthood (week 35). Data were analyzed using linear mixed models with genotype and sex as dependent variables, and subject as a random factor.

*Cohort 5 - Dendrite morphology.* As *Cadm2* is important for synapse organization [39], the fifth cohort (WT=3, HET=3, HOM=3) was used to quantify dendritic spines from medium spiny neurons in the nucleus accumbens (**NAc**), which is one of the core regions for impulsive behavior [40]. Only male mice were tested. ImageJ (Version 2.0.0-rc-69/1.52p) was used to analyze secondary and tertiary dendrites and score spine types. The results of the dendritic spine were averaged per 10μm for each mouse to standardize measures [41]. Statistical analyses were performed using ANOVA with genotype as a between-subjects factor.

Statistical analysis was conducted using SPSS 26.0 or 28.0 (IBM, Armonk, NY), the Bio-Medical Data Package (for BPM data) or RStudio (RStudio, PBC, Boston, MA). For all analyses, outliers deviating more than three times the interquartile range from quartile 1 or quartile 3 were excluded from the analysis. A *p*<0.05 was required for results to be considered statistically significant. Post-hoc differences were assessed using Bonferroni or Tukey’s honestly significant difference with a *p*<0.05. All analyses were performed with the researchers blind to the genotype.

**COHORT 1**

**Subjects and behavioral testing**

Mice (WT: 15 male, 10 female; HET: 16 male, 14 female; HOM: 1 male, 2 females) were administered a battery task in the following order on separate days (**Figure S30A**): Progressive Ratio Breakpoint Task (**PRBT**), Probabilistic Reversal Learning Task (**PRLT**), Iowa Gambling Task (**IGT**), Behavioral Pattern Monitor (**BPM**), and Pre-Pulse Inhibition (**PPI**). In all experiments, mice were littermate matched.

Mice were between 4 and 6 months of age at the time of testing. Mice were food restricted and maintained at 85% of their free-feeding weight during the periods of training and testing for PRBT, PRLT and IGT. All animals were housed with a maximum of five animals per cage and held in a temperature controlled reversed 12-h light cycle room (lights on at 7:00 P.M. and off at 7:00 A.M.) in a UCSD-operated vivarium. Mice were food deprived at 85% of their baseline body weight for IGT, PRBT, PRLT, and water was available ad-libitum. All procedures were approved by the UCSD Institutional Animal Care and Use Committee. The UCSD animal facility meets all federal and state requirements for animal care. Training and testing occurred between 12 PM and 5 PM. Food restriction ceased after two weeks of PRBT, PRLT, and IGT testing. Mice were tested in BPM and PPI twelve days later.

**Apparatus**

Training and testing for PRBT, PRLT and IGT took place in 5-hole-operant-chambers (25 × 25 × 25 cm, Med Associates Inc., St. Albans, VT). Each chamber consisted of an array of five square holes (2.5 x 2.5 x 2.5 cm) arranged horizontally on a curved wall 2.5 cm above the grid floor opposite a food delivery magazine (Lafayette Instruments, Lafayette, IN), at floor level and a magazine light near the ceiling. The chamber was in a sound attenuating box, ventilated by a fan that also provided a low level of background noise. An infrared camera installed in each chamber enabled the monitoring of performance during training and testing. Mice were trained to respond with a nose-poke to an illuminated LED recessed into the hole. Responses were detected by infrared beams mounted vertically located 3 mm from the opening of the hole. Liquid reinforcement in the form of strawberry milkshake (Nesquik® plus non-fat milk, 30 μl) was utilized and was delivered by peristaltic pump (Lafayette Instruments, Lafayette, IN), to a well located in the magazine opposite the 5-hole wall. Magazine entries were monitored using an infrared beam mounted horizontally, 5 mm from the floor and recessed 6 mm into the magazine. The control of stimuli and recording of responses were managed by a SmartCtrl Package 8-In/16-Out with additional interfacing by MED-PC for Windows (Med Associates Inc., St. Albans, VT) using custom programming.

**Habituation and Training**

Prior to training and testing for PRBT, PRLT, and IGT, mice were acclimated to the food reward by an overnight exposure to strawberry milkshake.

During the first training phase, mice were initially trained to nose-poke in the central aperture for a single food reward (fixed ratio, **FR1**). At initial training, mice were placed in the five-hole chambers for 10 minutes with milkshake dispensed every 15 seconds into the well of the lit magazine. Mice initiated each trial by nose-poking in the reward delivery area, after which the central light was immediately illuminated and a single nose-poke in the central aperture resulted in a single food reward. They were required to recognize magazine illumination and delivery of 30 μl strawberry milkshake as a reward and collect it every 15 seconds for 10 minutes (criterion was 30 collection responses per session for two consecutive days). Magazine entry resulted in the light being extinguished until the next reinforcement was delivered. Acquisition criterion was ≥30 entries in the reward magazine per session for two consecutive days. FR1 training then began wherein after entering the magazine, all 5 holes were illuminated and nose-poking in any of the 5 resulted in all five being extinguished, the magazine illuminated, and a reward being delivered. This FR1 training session lasted 30 minutes. To minimize biased responses in specific holes, five consecutive nose-pokes in one hole resulted in that hole being extinguished and inactive until two other holes were poked. This session was repeated daily until >70 responses were recorded within 30 minutes for two consecutive days. Once responding consistently, mice were baseline-matched on total responses and trained in a 30-minute habituation session prior to testing.

During the second training phase, mice were trained to nose-poke into one (PRBT), two (PRLT) or four (IGT) lit holes to obtain the reward, prior to testing in the respective test.

**Progressive Ratio Breakpoint Task**

During the 60 minutes of the PRBT, mice were required to nose-poke in the central illuminated hole in an incremental fashion in order to obtain a food reward. The number of nose-pokes required to gain a reward increased by one more than the previous addition every three trials, following the progression: 1, 1, 1, 2, 2, 2, 4, 4, 4, 7, 7, 7, 11, 11, 11, 16, 16, 16, etc. PRBT began when mice nose-poked into the illuminated magazine entry. Afterwards, mice had unlimited time to nose-poke into the central illuminated hole. The primary outcome measure of this task was the ‘breakpoint’, defined as the last ratio to be completed before the end of the session and quantified the willingness of an animal to work for a reward.

**Probabilistic Reversal Learning Task**

PRLT lasted for 60 minutes. Subjects were required to select the target hole between two illuminated holes within 10 seconds after the trial was initiated. The target hole provided a high probability of reward (80%) and low probability (20%) of punishment. The non-target hole provided a low probability of reward (20%) and high probability of punishment (80%). After 8 consecutive responses at the target hole, criterion was met and the target hole became the non-target hole and vice versa (reversals). The two holes were located at the left or right of the chamber and the target/non-target holes equally distributed within the cohort. The primary outcome measures of the task were total trials to criterion and number of reversals. The secondary outcome measures recorded in seconds included the mean latency to nose-poke a target hole (mean target latency), mean latency to nose-poke a non-target hole (mean-nontarget latency), and mean latency to collect a strawberry milkshake reinforcement when rewarded (mean reward latency). The final secondary outcome was accuracy, which was calculated by nose-pokes target hole/(nose-pokes target + non-target hole).

**Iowa Gambling Task**

Five-hole operant chambers were used for the IGT to provide four illuminated options (central hole was not illuminated). Mice had 10 s to nose-poke in one of the four illuminated holes. Mice were rewarded with strawberry milkshake (30 or 60 μl for safe and risky side, respectively), or punished with flashing light, which had a frequency of 0.5 Hz, for varying stimulus durations (6 or 12 seconds for safe side; 32 or 64 seconds for risky side). Two options delivered large rewards but long time-out penalties (disadvantageous/risky), whereas the other two options delivered smaller rewards but shorter time-out penalties (advantageous/safe).

Sessions lasted 60 minutes or 250 trials, whichever was completed first. Mice were required to nose-poke into the illuminated magazine entry to initiate the first trial. An intertrial interval (**ITI**) of 5 seconds preceded illumination of the cue array. If the mouse nose-poked in any cue hole during this 5 seconds ITI, a premature response was recorded and did not count as a completed trial, stimuli were not presented, and the magazine light illuminated for a 5 seconds time-out period in which all holes were unresponsive. The next trial began when the magazine light was extinguished and the mouse nose-poked into the magazine. If the mouse withheld from responding during the ITI period, holes 1, 2, 4, and 5 were illuminated. These lights remained lit until the mouse nose-poked in one of these holes or until 10 seconds had passed. Failure to respond in any hole during the light stimuli was registered as an omission. Omissions did not trigger a time-out period but resulted in the cue lights being extinguished and the magazine being illuminated so that another trial could be started. If the animal did nose-poke in 1 of the 4 lit holes during the stimulus, a ‘correct’ response and the hole choice were recorded. All cue lights were then extinguished and the mouse was rewarded or punished depending on the reward schedule. For rewards, the magazine light was illuminated and delivered the appropriate level of reinforcer. Retrieving the reward initiated the next trial. If a punishment occurred, no reward was given and a punishing time-out was triggered whereby the light stimulus of the chosen hole flashed at a frequency of 0.5 Hz for the duration of the time-out period, during which all apertures were unresponsive. After the time-out period, the flashing light was extinguished and the magazine light illuminated for a new trial to begin. These data were recorded as the total punishment duration in seconds. The time taken to make a choice (mean choice latency) and latency to collect rewards (mean reward latency) was also recorded.

The main outcome variable of this task was the difference score, which was determined by advantageous choices minus disadvantageous choices selected. Data were collected as P1, P2, P3, and P4 responses corresponding to different reward schedules. Responses were measured as a percentage of the total trials completed. Data were grouped by advantageous (P1 and P2) and disadvantageous options (P3 and P4) and response options were measured as a percentage of the advantageous choices (% Advantageous Choices = (P1+P2)/(P1+P2+P3+P4) × 100%), as described previously [27].

**Drug Preparation and Testing**

Drug solutions were prepared prior to testing. Ethanol (98%) was diluted with saline to 20% v/v; the injected volume was determined according to a body weight-dose schedule.

To evaluate the effect of acute doses of ethanol on the IGT in WT and HET mice, mice were injected i.p. with two different doses of ethanol (0.5 g/kg and 1.0 g/kg) or saline on three drug testing days with saline days in between. The drug dosages were selected based on our prior work [42]. There was one week between each testing session to reduce the risk of tissue damage. Each animal received a higher or lower dose next testing session to minimize the effect of ascending or descending. A pre-injection time of 15 minutes was applied. Ethanol challenge was administered after BPM and PPI (described below).

**Behavioral pattern monitor**

Methods and testing equipment for the mouse BPM have been described in detail previously [29,30]. BPM took place in a room with red light and eight BPM chambers (30,5 x 60 x 38 cm) with white light. Animals were transported to the testing room and allowed to acclimate for 1 hr prior to testing. Male mice were tested first on day 1 and females were tested second on day 2. Mice were placed in the upper left corner of the chamber. BPM lasted for 45 minutes.

Primary dependent measures were total activity, exploratory behavior, and locomotor patterns. Activity was measured by distance, counts (number of observed micro-events), and transitions (period when a mouse crossed an imaginary square). Exploratory behavior was measured by nose-pokes, repeated nose-pokes, and rearings. Locomotor patterns were measured by spatial d and spatial CV. Spatial d quantified straight-forward locomotor path of the mice. Spatial CV quantified consistent movement of the mice, in which a lower spatial CV constitutes a more random distribution of movement. Data were analyzed using ANOVA, with between-subjects factors of sex and genotype. Dependent variables were collapsed across sex where no main effect of sex was observed. Data were analyzed using Biomedical Data Programs software (Statistical Solutions Inc., Saugus, MA, USA).

**Pre-pulse Inhibition**

PPI took place in eight startle chambers (SR-LAB; San Diego Instruments, San Diego, California, USA). Each chamber contained a 5 cm diameter, clear, and nonrestrictive Plexiglas cylinder resting on a platform below high-frequency speakers that produced a constant background noise of 65 dB(A) and emitted the acoustic stimuli during the test. Mouse startle responses produce cylinder vibrations, which were converted to analog signals by an attached piezoelectric unit and stored as digitized data on a computer. At each stimulus onset, 65 consecutive 1 ms readings were obtained to determine the average amplitude of the acoustic startle response. SR-LAB equipment was calibrated regularly to ensure consistently accurate measurement.

The test session was designed to assess variations in both the pre-pulse intensity and the interstimulus interval (**ISI**) on the basis of previously published protocols [27,28]. Each session was initiated with 5 minutes of acclimation period during which the animals were habituated to the 65 dB(A) background noise for 25 minutes before the trials started. Each trial recording took 65 ms. The different PPI trials included pre-pulse, ISI and no stimuli. The PP trial consisted of a pre-pulse sound with varying intensity of 4,5, or 16 dB above background noise followed by a 120 dB pulse. The ISI trial was tested from the duration of onset pre-pulse to onset pulse which varied by 25, 50, 100, 200, or 500 ms; pre-pulse sound was 16 dB above background noise, followed by a 120 dB pulse. Every other trial was a no stimulus trial, in which no acoustic stimulus was presented. Startle pulses were presented for 40 ms, pre-pulse stimuli were presented for 20 ms, and the average intertrial interval between stimulus presentations was 15 seconds (range 7–23 s). The startle session was divided into five blocks. Blocks 1 and 5 each included five pulse-only trials, in which a 120 dB(A) pulse was presented alone. Block 2 assessed PPI and included four trial types (10 of each), including 120 dB(A) startle pulse intensities presented alone or preceded by 69, 73, or 81 dB(A) pre-pulse stimuli. Pre-pulses were administered 100 ms before the pulse stimulus. Block 3 assessed the startle response to different pulse intensities [80, 90, 100, 110, 120 dB(A)], but did not include any pre-pulse trials. In block 4, the ISI between pre-pulse and pulse varied; mice were presented with 120 dB(A) pulses alone or preceded by a 73 dB(A) pre-pulse separated by a 25, 50, 100, 200, or 500-ms interval (four trials for each interval).

The amplitude of the startle response was quantified as the average startle magnitude during the 65-ms recording window. Habituation to the startle response was assessed as the percentage decrease in startle amplitude in pulse-alone 120-dB(A) trials from block 1 to blocks 2, 3, 4, and 5. The percentage of PPI for each type of pre-pulse intensity was calculated as [100 − (pre-pulse amplitude/pulse amplitude) × 100].

**Statistical analyses for cohort 1**

Data were subjected to a univariate ANOVA with sex and genotype as between subject factors. The sample size of the HOM mice was low (N=3); therefore, we excluded this group from the analyses. Eight animals (5 WT and 3 HET) did not attain first criterion (>70 responses over 2 consecutive days) and were excluded from the analyses of trials to first criterion (PRLT). For the IGT, although performance was assessed over 3 trial blocks, as published previously and consistent with human testing, the overall data was analyzed and collapsed across blocks due to no block by genotype interaction. IGT, PRBT, and PRLT data were analyzed using IBM SPSS Statistics 25. BPM and PPI data were analyzed using Bio-Medical Data Package. Data obtained from the ethanol study were carried out using a repeated measure ANOVA with drug as a within subject factor and genotype and sex as between subject factors.

Startle responding and PPI were assessed separately for blocks 2, 3, and 4 using mixed analysis of variance (ANOVA; drug × genotype × sex) with pre-pulse intensity (block 2), pulse intensity (block 3), and ISI (block 4) as within-subjects factors. Further assessments used analysis of covariance (ANCOVA) with startle reactivity as a covariate to determine whether genotype differences in PPI may have been impacted by startle responding. Post-hoc differences were assessed using Tukey’s honestly significant difference (HSD) with a *p*<0.05.

Although some variables were affected by sex (**Table S21**), no sex by genotype interaction was observed for the primary outcomes. Therefore, analyses use the combined data.

**Results for cohort 1**

**PRBT**

We noted a trend for HET mice learning the PRBT task (>70 responses for 2 consecutive days) faster than WT mice [F_(2,74)_=4.01, *p*=5.10E-02], but we observed no effect of genotype on breakpoint [F_(1,51)_=0.003, *p*=9.57E-01; **Figure S32B**], which is the main measure of motivation examined in this task (**Table S21**).

**PRLT**

We observed a trend for a genotype effect on the number of trials completed [F_(1,42)_=3.095, *p*=8.50E-02], and a significant effect on the total number of trials completed to first reversals [F_(1,42)_=4.27, *p*=4.50E-02], HET mice requiring fewer trials to reach criterion than WT mice (**Figure S32C**). We also detected a significant effect of genotype on non-target latency [F_(1,51)_=5.15, *p*=2.80E-02], HET mice responding faster to a non-target than WT mice (**Figure S32D**). No effect of genotype was observed on any of the other PRLT measures (**Table S21**).
**IGT**	HET mice exhibited less risky behavior in the IGT (**Figure S33E**), as measured by a higher difference score [F_(1,51)_=4.70, *p*=3.50E-02] and total number of risky (disadvantageous) choices [F_(1,51)_=7.51, *p*=8.00E-03] compared to WT mice. Intriguingly, we observed a genotype effect on premature responses [F_(1,51)_=5.78, *p*=2.00E-02], which is another form of impulsivity, HET mice showing higher percentage of premature responses than WT mice.

No main effect of sex or interaction between sex by genotype was observed across any of the other IGT variables (e.g., percentage omissions, mean choice latency, main reward latency and total trials; **Table S21**).

Acute doses of ethanol did not differentially affect IGT performance (lack of ethanol effect, genotype effect or any interaction on any of the main outcome measures; **Table S21**).

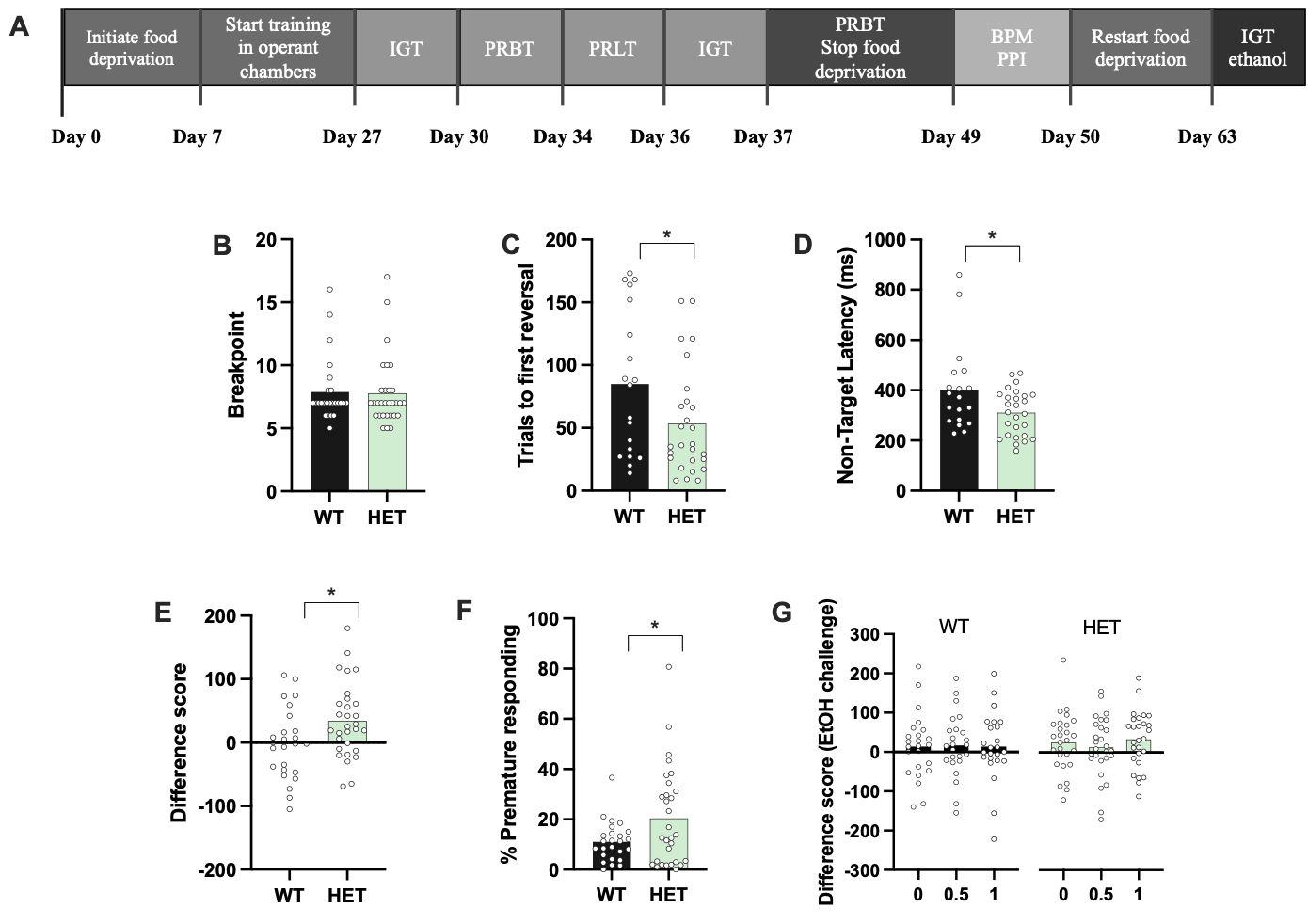

**Figure S32. *Cadm2* effects on motivation, behavioral flexibility and impulsive behavior as measured in the PRBT, PRLT and IGT tasks.** Experimental timeline for cohort 1 (**A**). *Cadm2* did not alter motivational behavior measured by breakpoint in the PRBT (**B**). In the PRLT, HET mice attained criterion faster (**C**) and responded to non-target stimuli quicker (**D**) than WT mice. In the IGT, HET mice showed lower risky responding, as measured via the difference score (**E**), but also showed higher premature responses (**F**) than WT mice. Acute doses of ethanol (0.5, 1 g/kg) did not modify IGT performance (**G**). Data are shown as mean ± S.E.M., * *p*<0.05.

**BPM**	HET mice exhibited more exploratory behavior than WT mice, as shown by an increase in nose-pokes [F_(1,53)_= 4.88, *p*=3.20E-02] and repeated nose-pokes [F_(1,53)_= 5.12, *p*=2.70E-02; **Figures S33A-B**]. There were no main effects of genotype on any of the additional variables tested, including rearings, distance, counts, transitions, spatial CV and spatial d (**Figures S33C-G**, **Table S21**). No sex by genotype interaction was observed on any measurement.

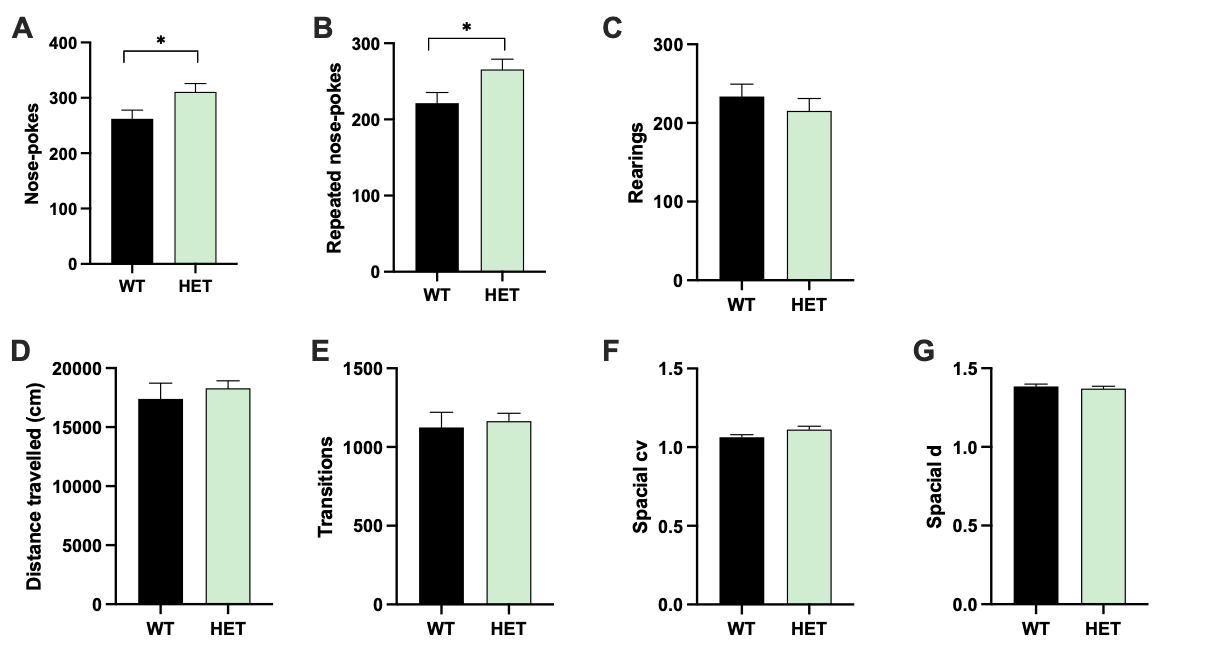

**Figure S33.** ***Cadm2* effects on activity, exploration and locomotor patterns.** HET mice exhibited higher exploration than WT mice in the BPM based on nose-pokes (**A**) and repeated nose-pokes (**B**), but showed no differences in other measures of exploration, such as rearings (**C**). No effect of genotype was observed on activity in BPM based on distance (**D**) or transitions (**E**). Mice exhibited equal amounts of repeated behavior in the BPM based on spatial CV (**F**). No difference in spatial d was observed (**G**). Data are shown as mean ± S.E.M, * *p*<0.05.

**PPI**
We detected a trend for a genotype effect on startle response [F_(1,51)_=3.99, *p*=5.00E-02], HET mice tended to startle more than WT mice (**Figure S34A**). Furthermore, a trend towards a pulse by genotype interaction was observed [F_(4,204)_=2.05, *p*=8.90E-02], where HET mice tended to startle relatively more at a higher pulse compared to WT. No sex by genotype interaction on startle response was observed [F_(1,51)_<1.0, ns].
There were no effects of genotype or sex on PPI (%) at different pre-pulse intensity, nor a genotype by sex or genotype by intensity interaction (**Figure S34B**, **Table S21**). However, when PPI was assessed across varying ISIs, a trend towards a genotype effect [F_(1,51)_=3.27, *p*=7.70E-02] and a significant ISI by genotype interaction [F_(4,204)_=2.93, *p*=2.20E-02] were found. Post-hoc analysis revealed a significant effect of genotype at the 25-ms ISI [F_(1,53)_=8.23, *p*=6.00E-03] and 100-ms ISI [F_(1,53)_=4.50, *p*=3.90E-02]. At those ISIs, HET mice exhibited higher % PPI compared to WT mice (**Table S21**). No differences were observed at 50-, 200- and 500-ms ISI (**Table S21**).
The effect of genotype on habituation was not significant and no genotype by sex interactions were observed (**Table S21**). However, we noted a significant effect of sex [F_(1,51)_=27.17, *p*<1.0E-04] and a habituation by genotype interaction [F_(4,204)_=2.14, *p*=1.6E-02]. Post-hoc analysis showed only a significant difference in habituation at the second 120 dB pulse [F_(1,53)_=4.80, *p*<0.05].

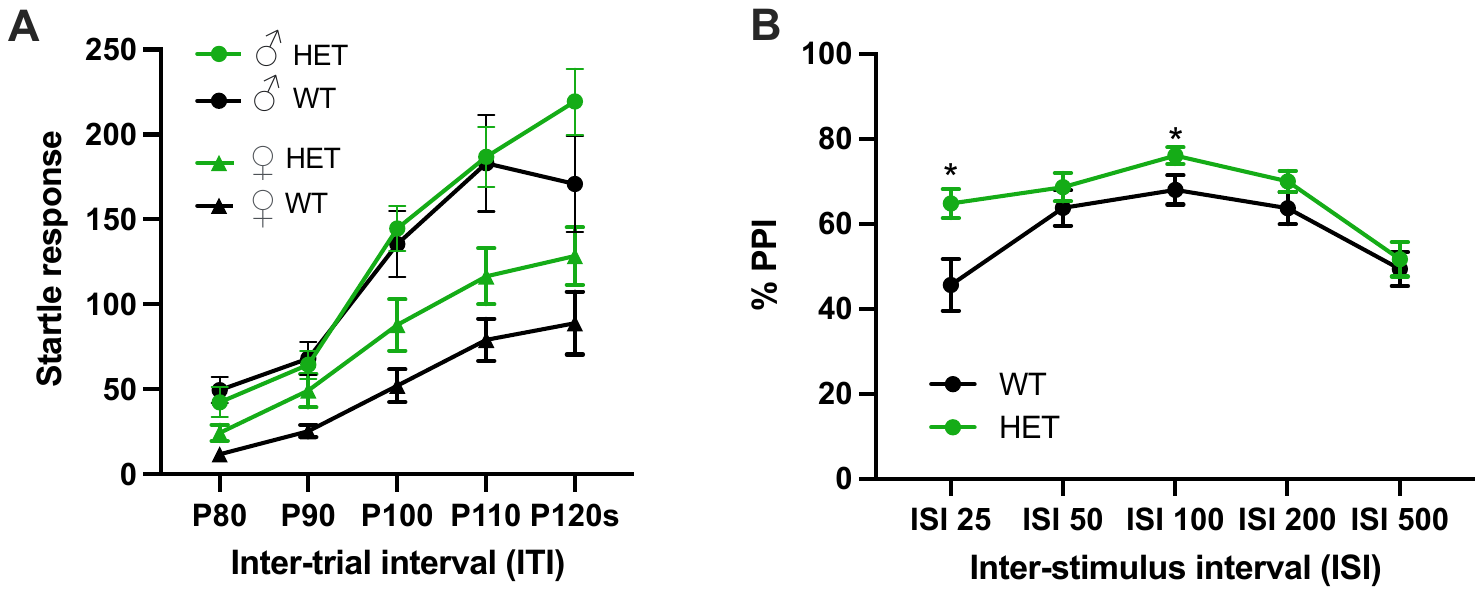

**Figure S34.** ***Cadm2* effects on sensorimotor gating behavior.** Genotype trend on startle response, with greater startle response in HET female mice compared to WT female mice (**A**). Percentage PPI shown at different ISIs (in ms). HET mice exhibited higher percentage PPI at ISI 25 and 100 ms compared to their WT littermates (**B**). Data are shown as the mean ± S.E.M. * *p*<0.05.

**COHORT 2**

**Subjects**

Mice (WT: 6 males, 7 females; HET: 7 males, 7 females; HOM: 8 males, 4 females) were tested on the 5-choice serial reaction time task (**5CSRTT**). In all experiments, mice were littermate matched. Mice were group-housed in pairs of 2-4. Mice were handled daily at least three days prior to behavioural testing. Animals were maintained on a 12h:12h light:dark cycle and had ad libitum access to food chow and water unless otherwise stated. The housing room was maintained at 21±2°C and at 50±10% humidity.

Beginning at eight to ten weeks of age, mice were food restricted and maintained at no less than 85% of their baseline weights prior to testing. Mice were given 18% protein food chow at 2.5-3.0g/day for males and 2.0-2.5g/day for females for the duration of behavioral testing. Weights were monitored daily and animals were given an extra 0.5-1.0g of chow if weight fell below the 85% threshold. All animal care, behavioral testing, and euthanasia were conducted in accordance with the Animal Use Protocol approved by the Animal Care Committee at the University of Guelph (AUP4194 and AUP3922).

**Behavioral testing**

***Touchscreen Apparatus: 5-choice serial reaction time task (5CSRTT)***

Each apparatus consisted of 4 mouse operant touchscreen chambers [43]. All procedures were conducted in the automated Bussey-Saksida Mouse Touchscreen System model 81426 (Campden Instruments Limited, Loughborough, EN). Each chamber was outfitted with a touchscreen that was partitioned with a plastic insert into five windows (132 x 132 pixels each) located 50 pixels from floor height. Chambers included a LED house light, a tone generator, a magazine unit with a light and photobeam to detect entries, and a pump connected to a bottle of liquid reinforcer (Neilson strawberry milkshake, Saputo Inc., Canada). ABET II Touch software v.2.20.3 was used to execute schedules and collect data, as we previously described [43–45].

**Behavioral sequence**

*Habituation to the reinforcer and to the 5CSRTT boxes.* Mice (10-12 weeks old) were food restricted and maintained at no less than 85% of their baseline body mass with 18% protein food chow during task training and testing. Training and testing were conducted 6 days per week as described by Beraldo et al. 2019 [31]. Habituation was conducted from days 1-3. On day 1, mice were placed in the testing chamber and were allowed to explore the apparatus for 10 minutes. The magazine light was turned off and no stimuli or reinforcer was presented. On days 2-4, mice were placed in the testing chamber for 20 minutes and 150 μL of reinforcer was delivered to the food magazine at session onset. Every 10 s, 7 μL of reinforcer was delivered to the food magazine. Reinforcer delivery was accompanied by a 1 second tone (3 kHz) and illumination of the food magazine. Day 3 was conducted in a similar fashion but for a longer session of 40 minutes.

*5CSRTT Training.* Training consisted of four stages: Initial Touch, Must Touch, Must Initiate, and Punish Incorrect. Mice underwent Initial Touch training for a maximum of 60 minutes or 30 trials per session, whichever occurred first. The Initial Touch session commenced with the magazine light off and the pseudorandom presentation of a stimulus light (white square) to one of the five touchscreen windows for a brief time (30 seconds). Following stimulus presentation, 7 μL of reinforcer was delivered to the illuminated food magazine and accompanied by a 1 second tone. A 5 seconds ITI between the next stimulus presentation began when the animal exited the food magazine. Delivery of the reinforcer was not contingent on a response, but if the mouse touched the screen while the stimulus was presented and where it was presented, the stimulus was removed and 21 μL of reinforcer was immediately delivered as described. Mice progressed to the Must Touch training phase when 30 trials were completed within one 60 minutes session. Must Touch was conducted as described for Initial Touch, except that subjects were required to touch the stimulus to receive the reinforcer. Mice progressed to the Must Initiate training phase when 30 trials were completed in one 60 minutes session. In Must Initiate, trials commenced with illumination of the magazine. Upon a nose-poke to the magazine, the light was extinguished, and a stimulus/reinforcer presentation was delivered as described for Must Touch. Each trial was separated by a 5 seconds ITI. Subjects progressed to the Punish Incorrect training phase upon completing 30 trials within 60 minutes. Punish Incorrect was conducted as described for Must Initiate, except that incorrect responses to the stimulus prompted a 5 seconds timeout period in which the house light was turned on and no reinforcer was delivered. After a 5 seconds ITI, the house light was turned off and the mouse was required to repeat the trial until a correct response was achieved. Mice progressed to the baseline training stage upon completing a minimum of 23 out of 30 correct trials across two consecutive 60 minutes sessions.

*Baseline Training:* The training phases were conducted in sessions that were a maximum of 60 minutes or 50 trials, whichever occurred first. Mice were placed into the testing chamber with the magazine illuminated and were requiring a nose poke into the magazine to initiate the first trial. Following a 5-8 seconds variable delay, the stimulus was presented pseudo-randomly in one of the five touchscreen windows for 4 s. The mice had an additional 5 seconds (limited hold) to respond on the screen following stimulus removal. A response during the stimulus presentation delay was registered as a “premature response” and initiated a timeout period as described above. Reinforcer or a timeout period were delivered following correct and incorrect responses, respectively. Trials in which the mouse failed to respond before termination of the limited hold period were considered “omissions” and followed by a timeout. Omissions and premature responses did not count towards performance accuracy. Mice proceeded to the 2 seconds baseline training when they completed three consecutive sessions with ≥ 80% accuracy, ≤ 20% omissions, and ≥ 30 trials completed. The 2 seconds baseline training sessions were conducted as described for 4 seconds baseline training, except that the stimulus was presented for 2 seconds. Baseline performance criterion was met when mice finished three consecutive sessions with ≥ 80% accuracy, ≤ 20% omissions, and 50 trials completed. Mice that failed to baseline performance criterion within 30 sessions were removed from the study. Following exclusion of poor performers (n=1 WT male, 1 HOM male), 13 WT (n=6 male, 7 female), 14 HET (n=7 male, 7 female), and 12 HOM (n=9 male, 4 female) were analyzed for 5CSRTT performance.

*Testing:* Performance in the following 5CSRTT parameters was evaluated: *Accuracy* – the percentage of trials in which a correct response was give; *Omissions* – the percentage of trials in which a response was omitted; *Session* *Time* – the length of time of the session; *Correct* and *Incorrect Response Latency* – the length of time for the mouse to input a response on the touchscreen; *Reward Collection Latency* – the length of time for the mouse to retrieve the reinforcer following a correct response; *Premature Response Percentage* – the percentage of premature responses in the total number of responses (%Prematures = Premature Responses / (Accurate Responses + Inaccurate Responses + Omitted Responses + Premature Responses) × 100%); and *Perseverative Correct* and *Perseverative Incorrect Responses* – the number of touchscreen inputs provided to the correct or incorrect touchscreen window after a response was already committed by the animal.

Modifying the conditions of the 5CSRTT can result in subtle differences in performance. For example, greater attentional demand can be evoked by shortening the duration of stimulus presentation [43–45]. Following establishment of the baseline testing, reduced stimulus duration (**RSD**) performance was evaluated using the same schedule parameters as described for 2 seconds baseline testing, except that the duration of stimulus presentation was fixed at 0.6, 0.8, 1.0 or 1.5 seconds within a session. Average performance was evaluated across two consecutive sessions of each stimulus duration. Between different RSD schedules, mice were returned to the 2 seconds baseline schedule for two consecutive sessions. The order of RSD schedules was counterbalanced within each genotype and sex.

Following RSD performance evaluation, the 2 seconds baseline criterion was re-established, and mice were tested in a 10 seconds stimulus delay session, commonly known as ‘long ITI’ [45]. This session was conducted as described for the baseline sessions, except that the delay between trial initiation and stimulus presentation was fixed at 10 seconds for all trials. Following this session, the 2 seconds baseline criterion was re-established, and performance was evaluated in an altered variable inter-trial interval (**vITI**) session. Within this session, the delay between trial initiation and stimulus presentation was 2, 5, 10 or 15 seconds. The delay used for each trial was pseudo-randomly determined.

*Two-bottle choice:* To evaluate any confounds in 5CSRTT performance that may arise from differences in preference for the reinforcer, a two-bottle choice preference test was conducted. Mice were singly housed in Allentown mouse cages containing corncob bedding and presented with two 10 mL serological pipettes outfitted with ball bearing sippers [46]. One pipette was filled with tap water and the other was filled with the strawberry milkshake reinforcer used for the 5CSRTT. After two hours, mice were returned to their home cages. Reinforcer preference was calculated as % Preference = Volume of Reinforcer Consumed / Total Volume of Liquid Consumed x 100%.

**Statistical Analysis for cohort 2**

Statistical analysis was conducted using SPSS 26 (IBM, Armonk, NY) or RStudio (RStudio, PBC, Boston, MA). For all analysis, outliers deviating more than three times the interquartile range from quartile 1 or quartile 3 were excluded from the analysis, and a *p* value < 0.05 was considered significant. Results are presented as the mean ± S.E.M. and graphs were generated in Prism 9.

Parametric data acquired from 5CSRTT and reinforcer preference testing were analyzed using one-way ANOVA or repeated measures, two-way ANOVA followed by Bonferroni post-hoc testing where appropriate. Non-parametric data were analyzed with Kruskal-Wallis one-way ANOVA or a two-way between-within subjects ANOVA on the trimmed means followed by Bonferroni post-hoc testing where appropriate. The total number of sessions required to reach 2 seconds baseline criteria was analyzed according to genotype. Standard performance in the 5CSRTT was evaluated as average performance during the final three sessions conducted during 2secondsbaseline training in which mice met the baseline performance criteria. RSD performance was determined as the average performance across the two consecutive tests conducted for each schedule. Reinforcer preference and standard performance were analyzed according to genotype. RSD performance was analyzed with genotype and duration as the between and within factors, respectively. For vITI testing, average performance during trials for each delay was calculated, and genotype and delay were used as the between and within subject factors, respectively.

**Results for Cohort 2**

***5CSRTT***

All mice showed similar levels of preference for the liquid reinforcer as measured in a 2 hour two-bottle choice test [F_(2, 36)_=0.73, *p*=4.88E-01; **Figure S35A**]. Time to achieve baseline criteria differed according to genotype [F_(2,36)_=7.42, *p*=2.00E-03], with HOM mice learning the task in fewer days than the WT (*p=*1.41E-02) littermates (**Figure S35B**).

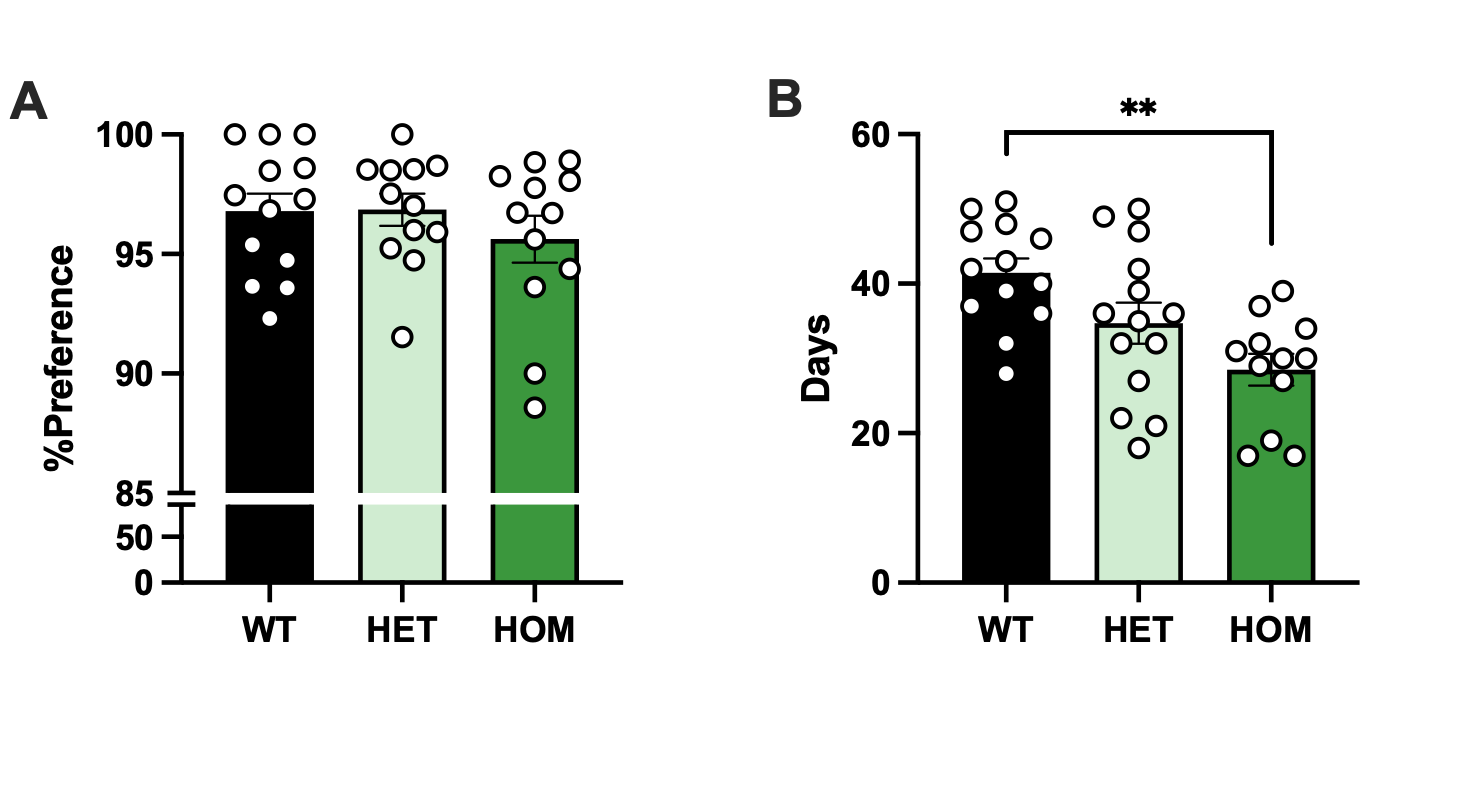

**Figure S35.** Preference for the milkshake reinforcer used in the 5CSRTT (**A**). Average number of training days according to genotype showed that HOM mice required fewer days than WT littermates to achieve baseline performance criteria in the 5CSRTT (**B**). Data are shown as the mean ± S.E.M., ***p*<0.01.

During baseline conditions, premature responses [F_(2,36)_=8.74, p=8.06E-04] and perseverative correct responses [F_(2,35)_=4.59, *p*=1.70E-02] were different across the genotypes (**Figure S36A-B**), with WT mice marginally (*p=*0.02) and HOM mice significantly (*p*=9.41E-04) showing significantly fewer premature responses than HET mice, and HOM mice making fewer perseverative correct responses than WT (*p*=1.38E-02) littermates.

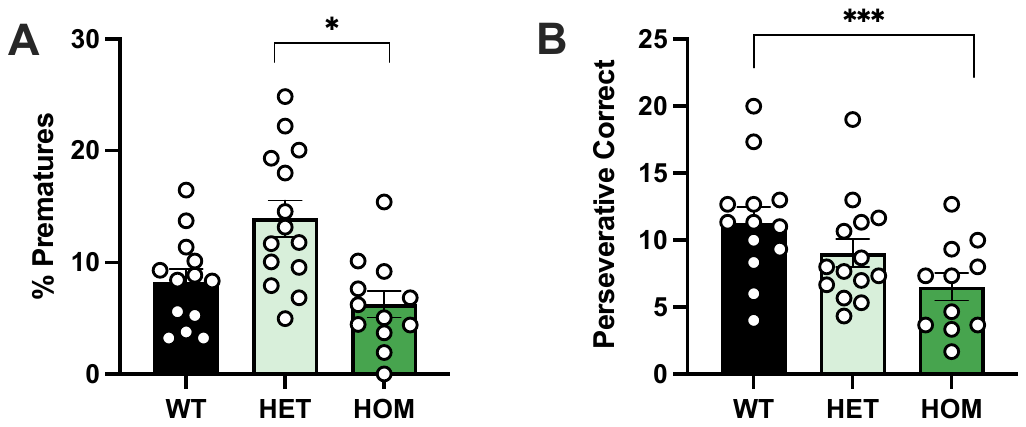

**Figure S36. 5CSRTT baseline performance.** Premature responses were lower in HOM and WT mice in comparison to HET mice (**A**). Perseverative responses were also lower in HOM mice compared to WT mice (**B**). Data are shown as the mean ± S.E.M. ****p*<0.001, **p*<0.05.

During the RSD challenge (**Figure S37**), there was a trend toward a main effect of genotype on premature response rate [F_(2, 11.49)_=3.67, *p*=5.90E-02], with WT and HOM mice committing marginally fewer premature responses than HET mice. A significant effect of genotype was also observed in accuracy rate [F_(2, 32)_=3.68, *p*=3.60E-02], with HOM mice showing poorer accuracy compared to WT littermates (*p*=3.90E-02). Overall, a main effect of stimulus duration was observed such that accuracy rate was lower [F_(3, 96)_=81.04, *p=*3.30E-26] and omission rate was higher [F_(3, 108)_=55.30, *p=*9.79E-22] when stimulus duration was shorter.

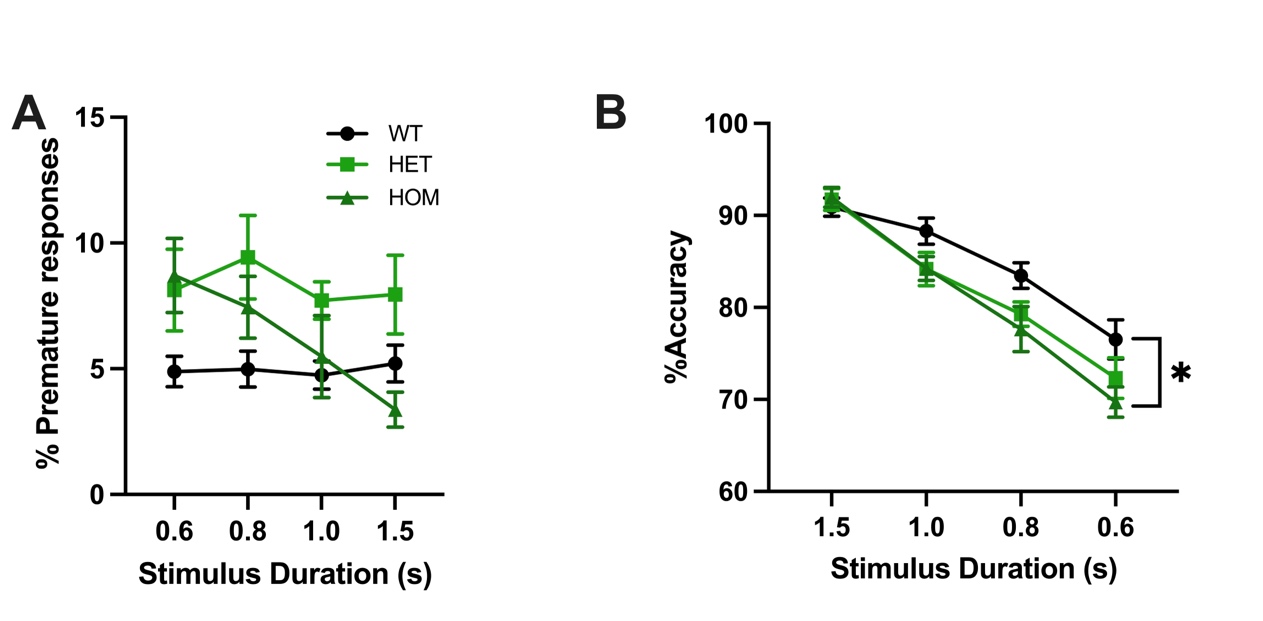

**Figure S37. Reduced Stimulus Duration (RSD) performance.** WT and HOM mice committed marginally fewer premature responses than HET mice during the RSD challenge (**A**). Accuracy was lower in HOM mice compared to WT mice (**B**). Data are shown as the mean ± S.E.M., **p*<0.05

During the long ITI (**Figure S38**), there was a main effect of genotype [H_(2)_=16.10, p=3.20E-04] on premature responding, with HOM mice showing fewer premature responses that HET (*p*=3.99E-04) and WT (*p*=6.64E-03) littermates. In addition, there was a significant main effect of genotype on session completion time [H_(2)_=18.51, *p*=9.58E-05] and perseverative incorrect responses [H_(2)_=11.88, *p*=2.64E-03], with HOM mice completing sessions faster than WT (*p*=9.83E-05) and HET mice (*p*=3.81E-03), and WT mice committing significantly fewer perseverative incorrect responses than HOM littermates (*p*=1.74E-03).

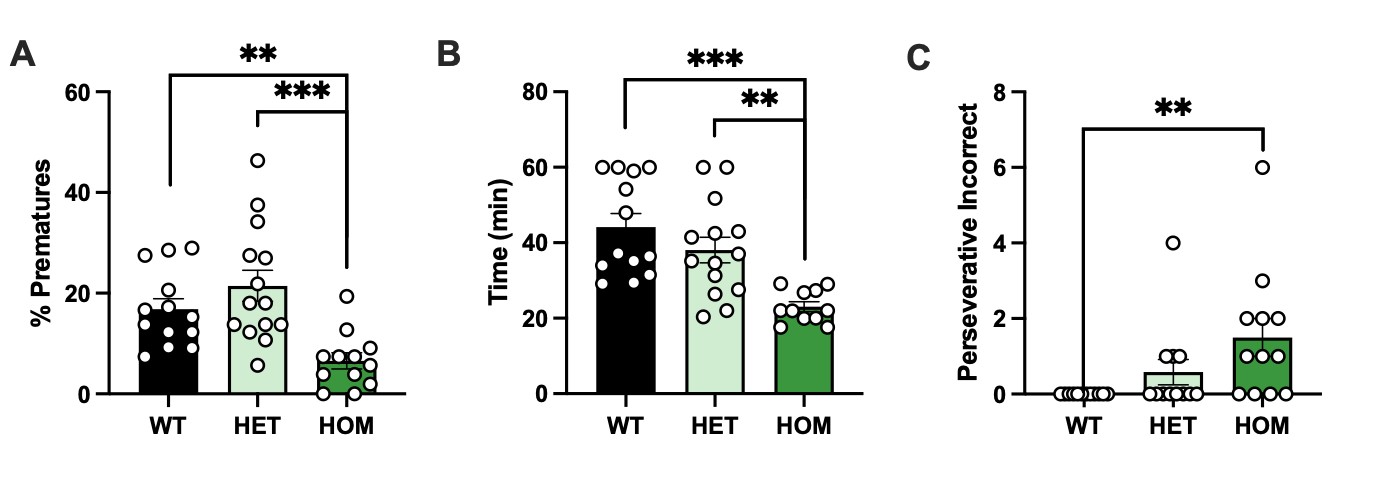

**Figure S38. Long ITI performance.** WT and HET mice committed significantly more premature responses than HOM mice during the long ITI challenge (**A**) and were slower to complete the session (**B**). WT mice committed fewer perseverative incorrect responses compared to HOM mice (**C**). Data are shown as the mean ± S.E.M., ****p*<0.001 ***p*<0.01.

There were few significant effects between groups under the vITI session (**Figure S39**). All mice showed less accurate responding [F_(3,108)_=5.87, *p*=9.42E-04] and more premature responses [F_(3,11.586)_=26.62, *p*=1.73E-05], but no main effect of genotype or interactions between delay and genotype were observed in any of the test measures.

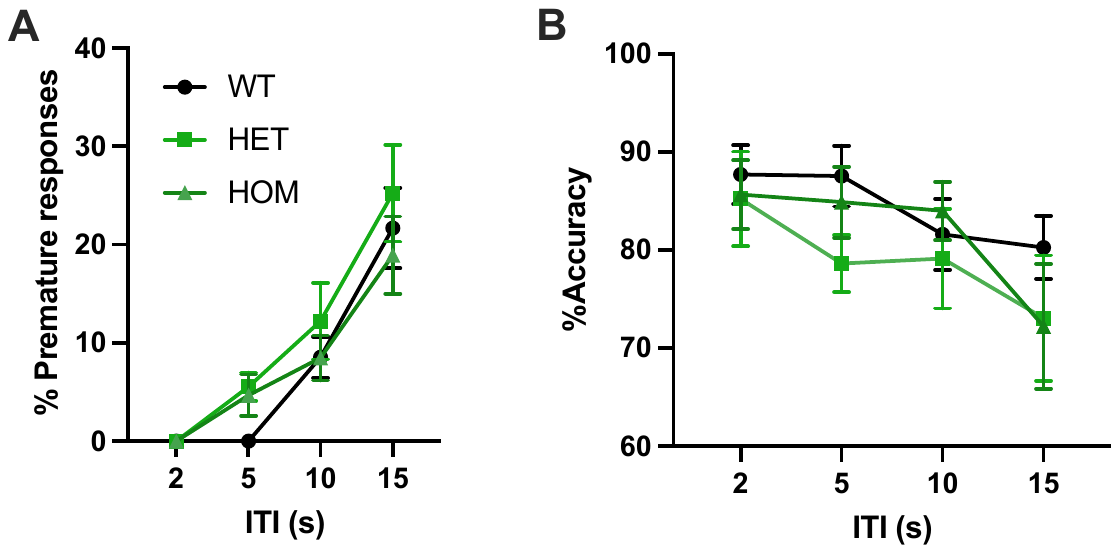

**Figure S39. vITI performance.** Greater premature responding and lower accuracy were observed with increases in the ITI time. No main effects of genotype were detected.

**COHORT 3**

**Subjects and behavioral testing**

To examine general locomotion, emotionality and ethanol drinking, mice (WT: 8 males, 9 females; HET: 20 males, 14 females; HOM: 9 males, 3 females) were administered a battery task in the following order: Open Field test (**OFT**), Elevated Plus Maze (**EPM**), Light-dark box (**LDB**), and drinking-in-the-dark (**DID**). In all experiments, mice were littermate matched. Mice were between 2-4 months of age at the time of testing. The mice were maintained under a 12h light/12h dark cycle with lights on at 0600 h. Unless otherwise specified, mice were housed (2-5 per cage) with access to food (Harlan 8604, Madison, WI, USA) and water *ad libitum*.

**Open field testing and apparatus**

The OFT was administered to measure locomotor activity, as previously described [32–35]. Mice were moved into the testing room and allowed to acclimate for at least 30 minutes prior to testing. Testing took place between 10:00 and 12:00 h. Mice were placed in the center of a square chamber (43 × 43 × 33 cm; the size of the center was 26 x 16 cm) with dim overhead lighting inside of sound and light attenuating boxes, and were allowed to freely explore for 30 minutes. A grid of infrared detection beams in each chamber and Versamax software was used to track animal location and locomotor activity (distance traveled) during the test. We also recorded the time spent in the center zone as a measure of anxiety-like behavior. The chambers were wiped down with a solution containing 30% ethanol between each animal to eliminate odors.

Measures taken were entries into the center, total locomotor activity (number of line crossings) and time spent in the center square (central zone). Number of rearings (data not shown) and defecations were also measured.

**Elevated Plus Maze, Light-dark box and apparatus**

We tested mice in two well-established tests of anxiety-like behavior, the elevated plus maze (**EPM**) test [36] and the light-dark (**LDB**) box test [37]. We have previously described the testing apparatus and procedures for LDB and EPM [33].

Immediately after the OFT testing, each animal was placed in the center of the EPM facing one of the open arms and allowed to explore the apparatus freely for 5 minutes. The EMP (Stoelting) consisted of 2 open arms (35 cm long × 5 cm wide) and 2 closed arms (35 cm long × 5 cm wide × 15 cm high) forming the shape of a cross. The apparatus was made from black Perspex and was elevated 40 cm above the ground. The room was illuminated with dim light and the open arms of the maze were under illumination of 15 lux. Measures taken included time spent in the open and closed arms and entries into the open and closed arms. An entry was defined as placing all four paws within the given arm.

One day after EPM, each animal was placed in the LDB. The LDB consisted of two compartments: a large, illuminated compartment (27 × 27 cm) and a small, dark compartment (18 × 27 cm) connected by a shuttle door (7.5 × 7.5 cm) located in the center of the partition at floor level. The light box was open at the top, painted white and illuminated by a 60 W bulb located 30 cm above the apparatus providing illumination in the range of 380-470 lux at floor level. The dark box was painted matt black and had a removable black lid at the top. Mice were placed at the center of the illuminated compartment, and the animal was allowed to freely explore both compartments for 5 minutes. Total number of crossings between the two compartments (defined the placement of all four paws in a given compartment), latency to enter into the dark compartment, latency to enter the illuminated compartment after the first entry into the dark box and time spent in the light compartment were measured.

Both the EPM and LDB were cleaned with a solution containing 30% ethanol after each 5-minute run and wiped dry before the next test.

In the EPM and LDB, the movement of each animal was recorded using a video camera (Sony SPT- M108CE) connected to a recorder to allow subsequent analysis, and scored by a human observer.

**Drinking-in-the-dark (DID) procedure**

Following 1-week habituation to the shifted light/dark cycle (lights off at 8 am), the animals were single-housed and trained to drink in the dark a 20% ethanol solution from spring loaded sipper tubes over a 1-week period, using the DID procedure [38].

Mice were exposed to the DID procedure for 5 consecutive days. 1 hour prior to the experiment, body weight was recorded, water bottles were removed to motivate drinking behavior, and the cages were placed on the experimental table. The animals were left undisturbed for 1 hour prior to the experiment. Starting 3 hours after lights off, a spring-loaded sipper tube attached to a 10-ml graduated pipette containing the experimental solution was inserted into the nozzle space in the cage lid where the water bottle was usually placed. Tubes were loaded with water (days 1 and 5) or a 20% ethanol solution (days 2-4). Animals were left undisturbed for 2 h. After this period, the volume of solution consumed was measured by reading from the bottom of the liquid meniscus in the pipette. Cages were returned to the holding rack and water bottles were replaced.

**Statistical analyses for cohort 3**

OFT, EPM and LDB data were assessed for normality and genotype effects were analyzed by ANOVA, with between-subject factors of genotype (WT, HET, HOM) and sex. For the DID, ethanol drinking data were averaged and converted from ml to g ethanol/kg body weight for analysis. Drinking data was analyzed using ANOVA, with between-subject factors of genotype (WT, HET, HOM) and sex. Water consumption was averaged (days 1, 5) and analyzed by ANOVA. A *p*<0.05 was required for results to be considered statistically significant.

**Results for cohort 3**

***OFT, EPM, LDB***

For the OFT, the total distance travelled in the boxes was different across the genotypes ([F_(2,70)_=7.53, *p*=1.00E-03], **Figure S40A**), HOM mice showing higher levels of locomotor activity than WT mice (*p*=1.40E-02). The distance travelled in the center, sometimes taken as a measure of anxiety-like behavior, was consistent across the genotypes ([F_(2,70)_=0.29, *p*=4.17E-01], **Figure S40B**). The number of defecations did not differ ([H_(2)_=0.10, *p*=9.52E-01]; **Figure S40C**).

**Figure S40** shows that there were no differences between the WT, HET and HOM mice in the EPM or LDB tests. In the EPM, WT, HET and HOM mice did not differ in the total number of arm entries [F_(2,78)_=0.16, *p*=0.85], again indicating normal locomotor activity (**Figure S40D**). HET and HOM mice made an equal number of entries into the open arms [F_(2,78)_=0.29, *p*=7.50E-01] and spent the same amount in the open arms [F_(2,78)_=0.21, *p*=8.13E-01] compared with WT mice (**Figure S40E-F**).

In the LDB test, HET and HOM mice spent an equal amount of time in the light compartment compared with WT mice [F_(2,76)_=0.66, *p*=5.22E-01]; **Figure S40I**); HET and HOM mice consistently performed the same as WT mice across all the other measures in this task ([F_(2,76)_<1.42, *p*>2.48E-01]; **Figure S40G-J**). Together, these data demonstrate that suppression of *Cadm2* does not modify anxiety-like behavior in these tasks.

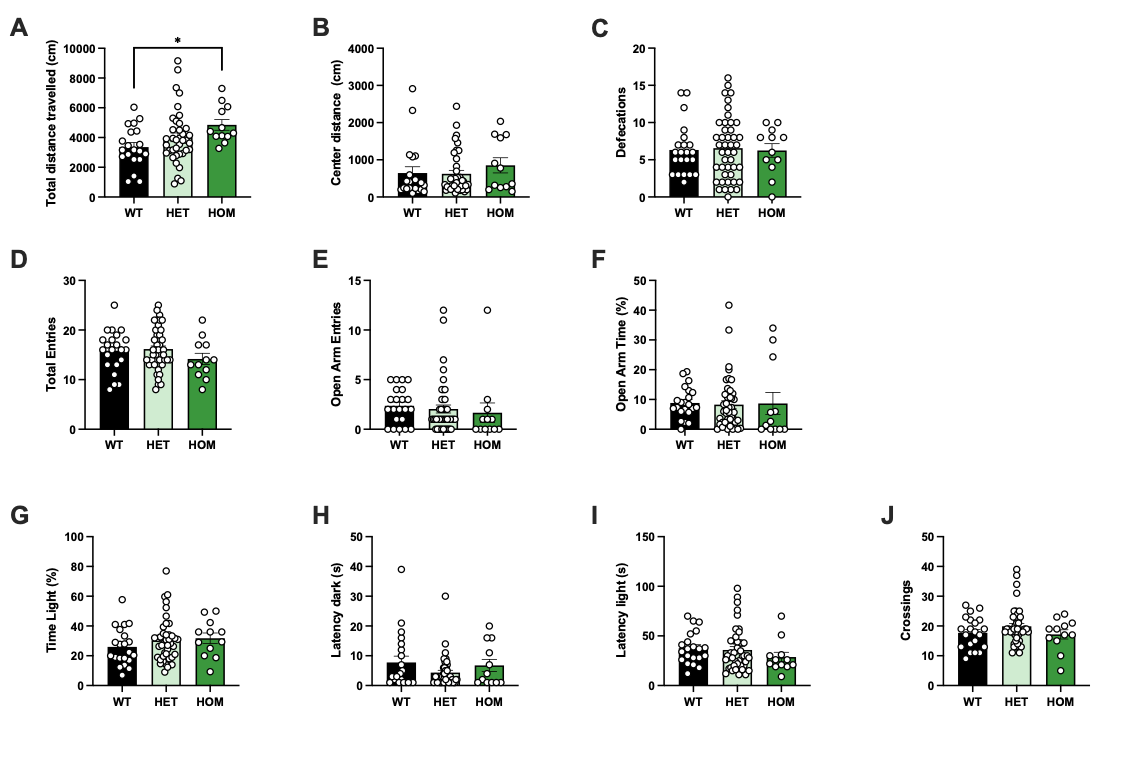
**Figure S40. Loss of *Cadm2* elevated total distance travelled in the Open Field (A-C) but did not modify anxiety-like behavior in the elevated plus maze (D-F), or the light-dark box (G-J).** HOM mice travelled a greater total distance in the apparatus compared to WT mice (**A**), but all genotypes spent equal time in the center (**B**) and displayed a similar number of defecations (**C**) during the OF task. Similarly, WT, HET and HOM mice displayed similar numbers of total (**D**) and open arm entries (**E**), and spent an equal percentage of time in the open arms (**F**) as measured in the EPM. Lastly, latency to enter the dark compartment (**G**) or to re-emerge to the light compartment (**H**) was equal between WT, HET and HOM mice, as was the percentage of time spent in the light (**I**) and the total number of crossings between compartments (**J**) in the LDB. Data expressed as mean ± S.E.M., * *p*<0.05.

***DID procedure***

Unexpectedly, we observed a genotype effect on the amount of water consumed ([F_(2,78)_=4.83, *p*=3.10E-02], **Figure S41A**), HOM mice drinking less water than WT (*p*=1.00E-03) and HET (*p*=1.00E-03) mice. The total amount of ethanol consumed did not differ across the groups ([F_(2,78)_=0.41, *p*=5.26E-01], **Figure S41B**). We observed sex effects in the total amount of water consumed (F_(2,78)_=8.07, *p*=1.00E-03), but none of the genotype by sex interactions were significant (**Figure S41B**).

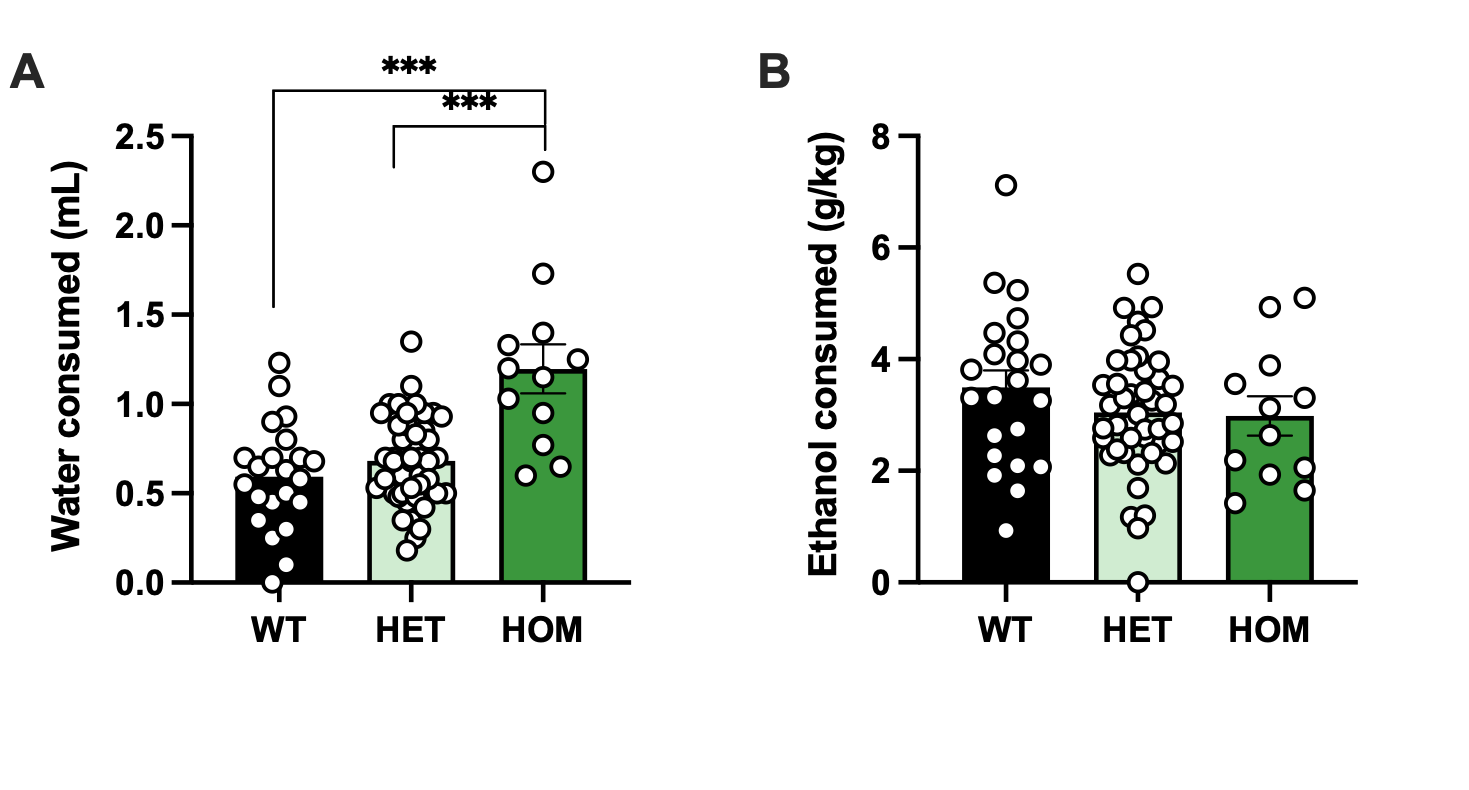

**Figure S41. Drinking-in-the-dark procedure.** The amount of data consumed was different across the genotypes (**A**), but we observed no differences in the total amount of ethanol consumed (**B**). Data expressed as mean ± S.E.M., *** *p*<0.001.

**COHORT 4**

**Subjects and body weight measurements**

Body weight (g) was monitored weekly for 27 weeks from 5-weeks to 35-weeks of age using a digital electronic balance. Mice (WT: 8 males, 14 females; HET: 30 males, 24 females; HOM: 9 males, 5 females) were littermate matched. The mice were maintained under a 12h light/12h dark cycle with lights on at 0600 h. Mice were housed (2-5 per cage) with access to food (Harlan 8604, Madison, WI, USA) and water *ad libitum*. We analyzed body weight from week 21 onwards (when body weights between groups started to diverge) using a linear mixed-effects model, with body weight as the dependent variable and genotype and sex as dependent variables. Subject was included as a random factor.

Relative to WT mice, there was a significant reduction in body weight in HOM mice (β=-3.74±1.27, p=0.004; **Figure S42**). The apparent trend towards a reduction in weight in HET mice was non-significant (β=-0.83±0.7, *p*=2.30E-01). Although there was a significant effect of sex (β=8.94±0.89, *p*<1.00E-03), there was no interaction between sex for HET (β = -1.12 ± 1.08, *p*=3.00E-01) or HOM mice (β = -1.92 ± 1.59, *p*=2.30E-01).

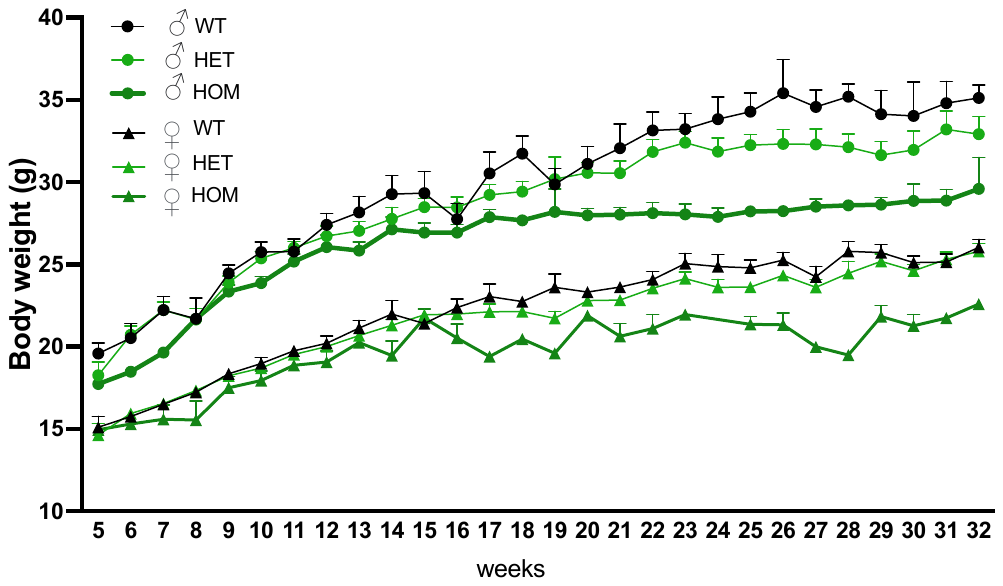

**Figure S42.** Body weight changes across the lifespan of WT, HET and HOM male and female mice. Data expressed as mean ± S.E.M.

**COHORT 5**

**Dendritic spine analysis**

To measure dendritic spines on medium spiny neurons (**MSN**) in the nucleus accumbens (**NAc**), we used particle-based biolistic delivery of the lipophilic dye DiI to dye-fill MSN in male mouse brain sections (WT N=3, HET N=3, HOM N=3) [47,48]. Mice were within 6-10 months of age to match the age from cohorts 1 and 2 at the time of the behavioral testing.

Deeply anesthetized mice were intracardially perfused with chilled saline for 5 minutes followed by another perfusion run with chilled 4% paraformaldehyde in PBS (Electron Microscopy Sciences) for 8 minutes at 1.5ml/min flow rate. Collected brains were postfixed in the same fixative for 1 hour at room temperature and later stored in PBS overnight at 4C. The brain tissues were sectioned using a vibratome into 150-μm thick sections containing the NAc (Bregma 1.70–1.18).

Dendritic spines were imaged as previously described [41]. Briefly, three slices from each brain were used for particle-based biolistic delivery of DiI using a Gene Gun system (Bio-Rad) with helium at 200 psi. Sections were mounted and imaged within 16–24 h of dye delivery to allow for dye diffusion within targeted neurons. MSN in NAc were imaged without differentiating between core and shell owing to the random labeling obtained with this approach that did not yield enough dye-filled neurons for separate analysis. Imaging was performed on a laser scanning confocal microscope (SP8; Leica Microsystems), with excitation at 543 nm. 5–10 dendrites per animal were imaged and Z-stacks were collapsed into projection images for analysis. The five most common dendritic spine categories include: thin, mushroom, stubby, filopodia, and branched/cup spines. Spine types were distinguished using morphometric criteria [49] as originally defined in studies of the rat cortex [50]. Briefly, spines with head bulb diameters much greater than their neck diameters and having thick stalks were classified as mushroom-shaped, spines that are short and thick and have similar head and neck diameters were scored as stubby, and spines with a slender stalk that expands into a small, oval or rounded end-bulb were classified as thin. For each spine type, total density was determined.

**Statistics for cohort 5**

Statistical analysis was performed using the ‘Statistical Package for Social Sciences’ (SPSS, version 28.0). All analyses were performed with the researchers blind to the condition.

ImageJ (Version 2.0.0-rc-69/1.52p) was used to analyze images of secondary and tertiary dendritic spines from neurons in the NAc. The results of dendritic spine density and type for each mouse was averaged per 10μm to standardize measures. Spine density for each dendrite type was averaged across nine images for each mouse. Statistical analyses of spine measurements were performed using ANOVA with genotype as a between-subject factor. A *p*<0.05 was required for results to be considered statistically significant.

**Results for cohort 5**

***Dendritic spine density***

Quantitative analyses of MSN in the NAc of WT, HET or HOM male mice (**Figure S43A**) revealed that there was no difference in dendritic spine density (total number of spines, **Figure S43B**) or type (thin, mushroom, stubby, **Figure 43C-E**; **Table S21**).

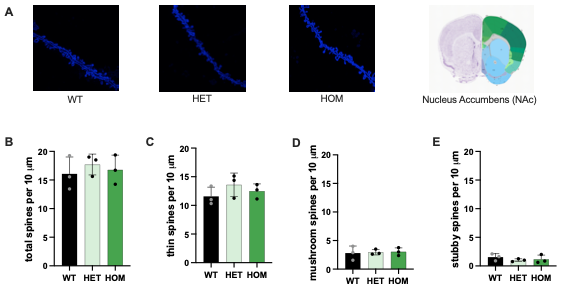

**Figure S43.** Dendritic spine density images (**A**) of MSN in the NAc of WT, HET and HOM male mice. There was no difference in dendritic spine density (**B**) or type (thin, mushroom, stubby, **C-E**). Data expressed as mean ± S.E.M.
